# Bebtelovimab, alone or together with bamlanivimab and etesevimab, as a broadly neutralizing monoclonal antibody treatment for mild to moderate, ambulatory COVID-19

**DOI:** 10.1101/2022.03.10.22272100

**Authors:** Michael Dougan, Masoud Azizad, Peter Chen, Barry Feldman, Matthew Frieman, Awawu Igbinadolor, Princy Kumar, Jason Morris, Jeffrey Potts, Lauren Baracco, Lisa Macpherson, Nicole L. Kallewaard, Dipak R. Patel, Matthew M. Hufford, Linda Wietecha, Emmanuel Chigutsa, Sarah L. Demmon, Bryan E. Jones, Ajay Nirula, Daniel M. Skovronsky, Mark Williams, Robert L. Gottlieb

**Author notes:** **Corresponding author:** Robert L. Gottlieb, MD, PhD, Baylor University Medical Center and Baylor Scott & White Research Institute, Dallas, TX, USA.

## Abstract

**BACKGROUND:** Bebtelovimab is a potent, fully human IgG1 monoclonal antibody (mAb) targeting the S-protein of SARS-CoV-2, with broad neutralizing activity to all currently known SARS-CoV-2 variants of concern, including omicron variant lineages. Specialized developmental approaches accelerated the initiation of a clinical trial designed to evaluate the efficacy and safety of bebtelovimab alone (BEB) or together with bamlanivimab (BAM) and etesevimab (ETE) delivered via slow intravenous push for the treatment of mild-to-moderate COVID-19.

**METHODS:** This portion of the phase 2, BLAZE-4 trial (J2X-MC-PYAH; NCT04634409) enrolled 714 patients (between May and July 2021) with mild-to-moderate COVID-19 within 3 days (≤3 days) of laboratory diagnosis of SARS-CoV-2 infection. Patients at low risk for severe COVID-19 were randomized 1:1:1 (double-blinded) to placebo, BEB 175 mg, or BEB 175 mg+BAM 700 mg+ETE 1400 mg (BEB+BAM+ETE). Patients at high risk for progression to severe COVID-19 were randomized 2:1 (open-label) to BEB or BEB+BAM+ETE, and a subsequent treatment arm enrolled patients to BEB+BAM+ETE using Centers for Disease Control and Prevention (CDC) updated criteria for High-risk. All treatments were administered intravenously over ≥30 seconds (open-label BEB) or ≥6.5 minutes (all other treatment arms). For the placebo-controlled patients (termed Low-risk), the primary endpoint was the proportion of patients with persistently high viral load (PHVL) (log viral load >5.27) on Day 7. For the open-label patients (termed High-risk), the primary endpoint was safety. In nonclinical studies, SARS-CoV-2 isolates were tested using an endpoint neutralization assay to measure BEB’s inhibitory concentration greater than 99% (IC_99_).

**RESULTS:** Baseline viral sequencing data were available from 611 patients; 90.2% (n=551) aligned with a variant of interest or concern (WHO designation), with the majority infected with delta (49.8%) or alpha (28.6%) variants. Among the Low-risk patients, PHVL occurred in 19.8% of patients treated with placebo, as compared to 12.7% (p=0.132) of patients treated with BEB+BAM+ETE and 12.0% (p=0.097) of patients treated with BEB, a 36% and 40% relative risk reduction, respectively. Viral load-area under the curve analysis from baseline to Day 11 showed statistically signficant reductions for patients treated with BEB (p=0.006) and BEB+BAM+ETE (p=0.043) compared to patients who received placebo. Time to sustained symptom resolution was reduced by a median of 2 days for patients treated with BEB (6 days; p=0.003) and 1 day for patients treated with BEB+BAM+ETE (7 days; p=0.289) compared to placebo (8 days). The incidence of COVID-19-related hospitalization or all-cause deaths by day 29 were similar across treatment arms, as expected given the patients’ risk status (the Low risk cohorts had a Low risk of hospitalization, and High risk cohorts received only active therapy without placebo). Overall, safety results were consistent with previous studies investigating mAbs targeting SARS-CoV-2. The proportion of patients with treatment emergent adverse events (AEs) were 9.7% in Low-risk (n=37/380) and 14.7% in High-risk (n=48/326) patients treated with BEB or BEB+BAM+ETE; majority of AEs were considered mild or moderate in severity. Serious AEs were reported in 2.1% of High-risk patients (n=7/326), including one death (a cerebrovascular accident); 1 serious AE was reported among Low-risk patients. In an in vitro neutralization assay, BEB neutralized the omicron isolate (BA.1) with <2.44ng/ml estimated IC_99_.

**CONCLUSIONS:** In patients with mild-to-moderate COVID-19, treatment with BEB or BEB+BAM+ETE was associated with greater viral clearance, a reduction in time to sustained symptom resolution, and safety results consistent with mAbs that target SARS-CoV-2. Integration of clinical findings with in vitro neutralization of emerging viral variants offered a pragmatic framework for investigating the efficacy of a new antiviral mAb agent, as demonstrated by bebtelovimab.

## INTRODUCTION

The distribution of variants has rapidly evolved over the course of the Coronavirus Disease 2019 (COVID-19) pandemic contributing to surges in new cases and has led to escape from some effective treatments, including selected neutralizing monoclonal antibodies (mAbs). As vaccination programs continue globally and vaccine-based immunity develops, there is an ongoing need for safe and efficacious treatment options for unvaccinated individuals and those at increased risk for severe disease, particularly during times of emerging viral variants of concern.

Neutralizing mAbs offer effective, passive immunization to recipients by directly interfering with virus entry and pathogenesis^2-4^. For example, the neutralizing mAbs bamlanivimab (BAM) and etesevimab (ETE) target the receptor binding domain (RBD) of the spike protein of SARS-CoV-2, thereby inhibiting viral attachment and entry into target cells^5-8^. In outpatients at increased risk for severe COVID-19, infusion of BAM or BAM+ETE was associated with improved clinical outcomes, such as reductions in COVID-19-related hospitalizations^9-11^. Positive clinical outcomes with BAM+ETE were associated with rapid viral clearance as well as the absence of persistently high viral load (PHVL) seven days following treatment^10^.

By November 2021, a number of authorized neutralizing mAbs were available in the United States that were effective against the delta variant, the predominant variant strain at the time; however, the emergence of the omicron (B.1.1.529) variant threatened the utility of many existing antibody treatments, including BAM+ETE^4,12^. The rapid spread of the omicron variant reinforced the need to preemptively identify and develop additional antibodies capable of neutralizing new variants, and emphasized the need to develop creative approaches to expedite clinical development and production of mAbs in order to meet evolving public health needs.

Leveraging the extensive clinical evidence that support the efficacy of neutralizing mAbs in reducing SARS-CoV-2 viral load^9,10,13-16^, candidate mAbs that maintain potency against emerging variants can be identified through the use of in vitro assays and pharmacokinetic (PK) modeling approaches^17^. In vitro virus neutralization experiments have shown to be consistent with clinical activity of authorized neutralizing mAbs, including their use for determining when an antibody should and should not be used for a particular variant^4,18,19^. Development of a candidate mAb can also be expedited by proactive communications with regulatory authorities, use of innovative clinical study designs, leveraging platform processes and methods, use of efficient cell culture approaches for large-scale antibody manufacturing, streamlined manufacturing testing, and running parallel processes. Collectively, these efforts are designed to bring new COVID-19 therapeutic medicines more rapidly to the clinic.

Originally derived from a convalesent COVID-19 patient and identified in April 2020, bebtelovimab (BEB; also known as LY-CoV1404, 1404) is a potent neutralizing mAb targeting the RBD of the SARS-CoV-2 S-protein, with an epitope distinct from BAM and ETE^16,20^. In vitro pseudovirus and authentic SARS-CoV-2 virus neutralization assays demonstrated that bebtelovimab’s activity was maintained against all currently known variants of interest and concern (including the alpha, delta, and omicron lineages and sublineages) as of January 2022^16,20^. Due to the potency of BEB, PK models indicated that the drug would be effective in adults at 175 mg, which would permit intravenous (IV) push of the medication as opposed to an IV infusion, reducing the need for specialized resources.

Herein, we investigated the PK, efficacy, and safety results of BEB treatment in the Phase 1/2 BLAZE-4 trial (NCT04634409). In a Phase 1 single ascending-dose addendum of BLAZE-4, increasing doses and infusion rates of BEB were investigated in patients with mild-to-moderate COVID-19. In the subsequent Phase 2 portion of the trial, we evaluated the efficacy and safety results of BEB alone or in combination with BAM+ETE in patients with mild-to-moderate COVID-19. The results from these studies, along with in vitro virus neutralization results, contributed to the emergency use authorization (EUA) of BEB for the treatment of ambulatory COVID-19 patients at increased risk for severe disease.

## METHODS

### STUDY DESIGN

This portion of the BLAZE-4 trial (NCT04634409) is a randomized, single-dose, Phase 1/2 study in ambulatory patients in the United States, presenting with mild-to-moderate COVID-19, within 3 days of first positive test for SARS-CoV-2 infection. The study evaluated early intervention with neutralizing mAbs to SARS-COV-2 in reducing overall viral load. The efficacy and safety of single dose IV infusions of bebtelovimab alone (BEB) or together with BAM+ETE (BEB+BAM+ETE) were examined in patients at lower (term “Low-risk”) or increased (term “High-risk”) risk of severe COVID-19. High-risk patients were ≥ 12 years of age at the time of randomization and required to have at least 1 risk factor for developing severe COVID-19, based on the Centers for Disease Control and Prevention guidance^21^. Low-risk patients were those who were between 18 to 64 years of age at the time of randomization and did not have the risk factors defined as part of the High-risk criteria. Full inclusion and exclusion criteria can be found in the Supplementary Material.

In the Phase 1 portion of the trial, we investigated ascending doses and infusion rates of IV administration in Low-risk patients. Each BEB cohort represented a dose increase or infusion rate change (increase in mg/minute) from the preceding cohort. The first two patients (double-blinded, randomized 1:1 placebo to active treatment) were sentinel; safety and tolerability were reviewed for these patients for up to 24 hours after dosing and subsequent patients were randomized 1:5 placebo to active treatment. Overall study design of the Phase 1 portion of the trial can be found in the Supplementary Marterial.

In the Phase 2 portion of the trial, Low-risk patients were randomized 1:1:1 (double-blinded) to placebo, BEB 175 mg, or BEB 175 mg+BAM 700 mg+ETE 1400 mg (BEB+BAM+ETE) (placebo-controlled). Patients at high risk for severe COVID-19 were randomized 2:1 (open-label) to BEB or BEB+BAM+ETE. A subsequent treatment arm enrolled patients to BEB+BAM+ETE using updated CDC criteria for High-risk^21^. Given the availability of efficacious mAbs, no placebo control was implemented in the High-risk patient cohort and therefore the focus was on safety; the placebo-controlled portion focused on investigating viral surrogates in younger/healthier subjects (Low-risk). Treatments were administered intravenously, in a single dose, ≥30 seconds (open-label BEB) or ≥6.5 minutes (all other treatment arms). In the Phase 1 and 2 portions of the trial, post-treatment follow-up assessments were conducted at Days 60 and 85 to assess clinical status and for adverse events (AEs). A patient was considered lost to follow-up if they repeatedly failed to return for scheduled visits and was not contactable by the study site.

This trial was conducted in accordance with the consensus ethical principles of the Declaration of Helsinki, the International Council for Harmonisation Good Clinical Practice Guidelines, the Council for International Organizations of Medical Sciences International Ethical Guidelines, and applicable laws and regulations. Patients or their legally authorized representatives provided written informed consent and child/adolescent assent, as appropriate, prior to participation.

### BASELINE SARS-CoV-2 VIRAL SEQUENCING

Viral nucleic acid was extracted from the baseline sample and next-generation sequencing was preformed as described in, Gottlieb et al. Viral lineage determined using Pangolin (Version 3.1.11^22^) using the assignment algorithm pangoLEARN (24 August 21 release with pango-designation Version 1.2.66). High level strain definitions were as assigned by the embedded Scorpio software (Version 0.3.12, constellations Version 0.0.15), which uses defined constellations of mutations to attribute a summary viral strain classification to a sequence.

### SYMPTOMS AND OVERALL CLINICAL STATUS

Outpatients rated the severity of their COVID-19-related symptoms and overall clinical status using a daily questionnaire^9^ consisting of three domains describing (i) the severity of symptoms (ii) general physical health, and (iii) change in overall health. Symptom severity was scored daily by the patient as experienced during the past 24 hours; none or absent = 0; mild = 1; moderate = 2; severe = 3. Symptom resolution was defined as the first assessment with a score of 0 for shortness of breath, feeling feverish, body aches and pains, sore throat, chills and headache; and a score of 0 or 1 for cough and fatigue. The time to symptom resolution was defined (in days) as the first study day when symptom resolution status is “Yes” (date of infusion defined as Day 1). Sustained symptom resolution was defined as the first study day of 2 consecutive assessments with symptom resolution. Loss of taste and loss of smell were assessed by yes/no questions. A complete physical examination was performed at screening and symptom-directed physical examinations performed at select visits. Investigators paid special attention to clinical signs related to COVID-19, historical and ongoing medical conditions. Vital signs including body temperature, blood pressure, pulse and respiration rate, saturation of peripheral oxygen and incident requirement for supplemental oxygen were measured. To define baseline serostatus, a qualitiative assay (Roche, Elecsys® Anti-SARS-CoV-2) confirmed the presence or absence of antibodies specific only to the nucleocapsid protein of SARS-CoV-2.

### DOSE JUSTIFICATION

The IV target therapeutic doses of 175 mg BEB, 700 mg BAM, and 1400 mg ETE were selected using PK/PD modeling^17,24^ in order to identify a dose resulting in a drug concentration above IC_90_ in ≥90% of patients for ≥28 days, and in maximum viral load reduction^13,23^. The safety, tolerability, PK and pharmacodynamics (PD) of BAM alone or together with ETE (BAM+ETE) were determined in prior clinical studies (PYAA, PYAB, PYAH)^9,13,23^.

### OUTCOMES

For the Phase 1 portion of the BLAZE-4 trial, the primary objective was to characterize the safety and tolerability of BEB alone and BEB+BAM+ETE after IV infusion. For the Phase 2 portion of the trial, the primary endpoint for the placebo-controlled patients (Low-risk), was the proportion of patients with persistently high viral load (PHVL) (log viral load >5.27) on Day 7. For the open-label patients (High-risk), the primary endpoint was safety outcomes. Secondary outcomes included, but were not limited to, evaluation of SARS-CoV-2 viral load, COVID-19-related hospilitizations (≥24 hours of acute care) and all-cause mortality by Day 29, and time to sustained symptom resolution.

### STATISTICAL ANALYSES

All enrolled patients who received any amount of study intervention were included in the Safety population. All patients in the Safety population who additionally provided at least one post-baseline measurement were also included in the Efficacy population.

For the placebo-controlled cohort (Low-risk), the sample size was determined with 84% power and an assumed PHVL rate of 28% in the placebo arm and 12% in the BEB and BEB+BAM+ETE arms at Day 7. The assumed PHVL rate was determined from previous treatment arms enrolling Low-risk patients. A hierarchical multiple comparisons procedure, which controlled type I error for the primary endpoint analysis, was implemented. First, BEB+BAM+ETE was compared to placebo and then BEB alone was compared to placebo for the proportion of patients with PHVL on Day 7. All hypothesis tests were 2-sided at an alpha level of 0.05.

Treatment comparisons for binary endpoints were made using logistic regression with a Firth penalized likelihood (Firth 1993). Where the number of observed events was fewer than 5 in any treatment arm, an exact test (i.e., Fisher’s exact) was conducted instead of using a logistic regression. Missing data was imputed using last observation carried forward.

Treatment comparisons for change from baseline in viral load were made using a mixed-effects model repeated measures (MMRM) analysis that included: (a) treatment group, (b) baseline value, (c) visit, and (d) the interactions of treatment-by-visit as fixed factors. The Kenward-Roger method was used to estimate the denominator degrees of freedom. Type III sums of squares for the least squares (LS) means was used for statistical comparison. For all change from baseline analyses, patients who did not have a valid baseline measure were excluded.

Treatment comparisons of continuous efficacy, safety, and health outcome variables with a single post-baseline timepoint were made using analysis of covariance (ANCOVA) with: (a) treatment group, and (b) baseline value in the model. Type III sums of squares for LS means was used for statistical comparison between treatment groups.

The Kaplan-Meier (KM) product limit method was used for time-to-event analyses. For High-risk patients, no treatment comparisons were peformed. All outcome variables are reported descriptively.

### SARS-CoV-2 VIRUS MICRONEUTRALIZATION ASSAY

SARS-CoV-2 isolates were tested using an microneutralization assay (Dr. Matt Frieman laboratory, University of Maryland School of Medicine, Baltimore, MD, USA). This quantitative neutralization assessment measures the antibody concentration that is necessary for complete viral inhibition, which is equivalent to an IC_99_ value. Three different SARS-CoV-2 isolates were tested from three different lineages; WA1 (Washington 1 isolate; 2019-nCoV/USA-*WA1*/2020, CDC [USBI, BEI]), delta (B.1.617.2; obtained from Dr. Andy Pekosz at Johns Hopkins Bloomberg School of Public Health; hCoV-19/USA/MD-HP05285/2021), and omicron (B.1.1.529/BA.1); obtained from Dr. Mehul Suthar at Emory; GISAID accession #EPI_ISL_7171744). SARS-CoV-2 neutralization was performed similarly as described in Keech et al. (2020)^25^ with some modifications described herein. Samples were diluted in duplicate to an initial concentration of 20 µg/ml followed by 1:2 serial dilutions, resulting in a 12-dilution series with each well containing 60 μL. 60 μL of diluted SARS-CoV-2 inoculum was added to each well to result in a multiplicity of infection = 0.01 upon transfer to 96-well titer plates and a 1:2 sample dilution, resulting in an initial concentration of 10 µg/ml. Non-treated, virus-only control, and mock infection controls were included on every plate. The antibody and virus mixture was incubated at 37°C (5% CO2) for 1 hour before transferring 100 µl to 96-well titer plates with confluent Vero/TMPRSS2 cells. Titer plates were incubated at 37°C (5% CO2) for 72 hours for most variants, but 96 hours for omicron, followed by cytopathic effect (CPE) determination for each well in the plate. The first sample dilution to show CPE was reported as the minimum sample dilution required to inhibit (neutralize) greater than 99% of the concentration of SARS-CoV-2 tested; this was expressed as an inhibitory concentration of greater than 99% (IC>99).

## RESULTS

### Patients

Between April 19^th^ to July 19^th^, 2021, a total of 40 patients with mild-to-moderate COVID-19 were randomized to receive BEB, BEB+BAM+ETE or placebo in the Phase 1 intravenous portion of the BLAZE-4 trial. The IV target therapeutic doses of 175 mg BEB, 700 mg BAM and 1400 mg ETE were selected using PK/PD modeling, in order to identify a dose resulting in a drug concentration above IC_90_ in ≥90% of patients for ≥28 days, and in maximum viral load reduction. Patient demographic and baseline characteristics, as well as efficacy and safety results are included in the Supplementary Material. PK and safety results obtained from the Phase 1 portion (dose 1, 2a, 2b, and 2c, Figure S1) informed the dosing strategy implemented in the Phase 2 study, following confirmation of no reports of COVID-19-related hospitalizations or deaths and verification that increased doses and infusion rates of BEB were not associated with increased treatment emergent adverse events (TEAEs) through at least 24-48hrs.

The Phase 2 portion of the BLAZE-4 trial commenced once the BEB+BAM+ETE cohort from the Phase 1 portion of the BLAZE-4 trial dosed all patients and safety data was assessed for at least 48 hours after the final IV infusion. Overall, 706 patients with mild-to-moderate COVID-19 were enrolled and dosed. Patients in the double-blinded, Low-risk cohort were enrolled between May 7^th^ 2021 through July 21^st^ 2021 and randomized to receive BEB (N=125), BEB+BAM+ETE (N=127) or placebo (N=128). Patients in the open-label, High-risk cohort were enrolled between May 7^th^ 2021 through June 21^st^ 2021 and randomized to receive BEB (N=100) or BEB+BAM+ETE (N=50). Those enrolled according to the updated CDC criteria (that lowered BMI from ≥ 35 to >25, among other changes) were enrolled between June 21^st^ 2021 through July 12^th^ 2021 and also received the combination therapy of BEB+BAM+ETE (N=176) (Figure 1). All patients were centrally randomized to study intervention using an Interactive Web Response System. Patients were stratified by duration since symptom onset to randomization (≤8 days versus >8 days). High-risk patients were also stratified by whether they received any dose of a SARS-CoV-2 vaccine, or not, prior to screening; Low-risk patients were exclusively unvaccinated against SARS-CoV-2.

**Figure 1:**
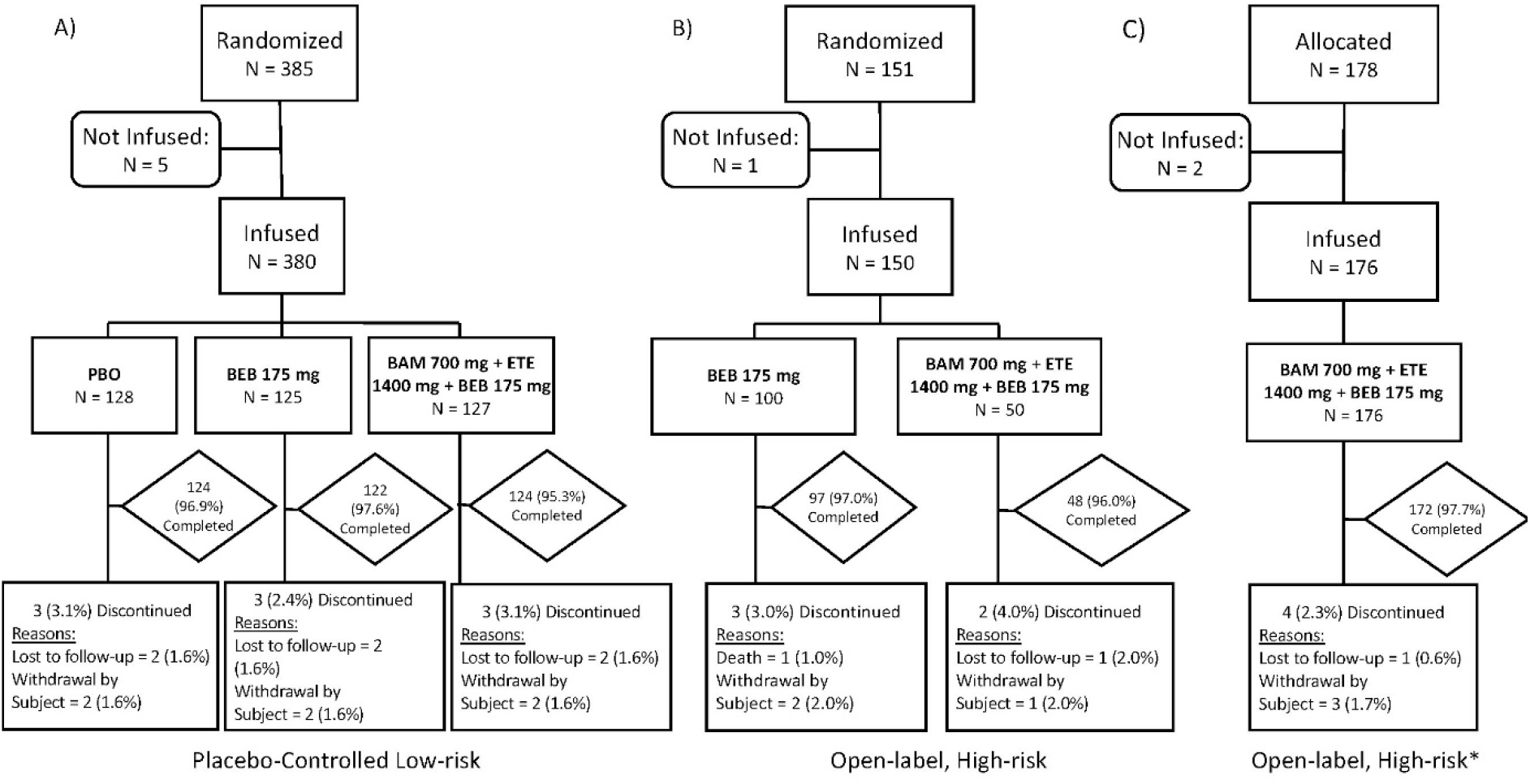
Study design and patient treatment disposition across the treatment period (through Day 29) in (A) placebo-controlled, Low-risk patients (B) open-label, High-risk patients and (C) open-label, High-risk patients (*updated CDC criteria).

Baseline sequencing data available for 611 patients, revealed that 90.2% (n=551) of the sequences obtained aligned with a variant of interest or concern (WHO designation), with the majority of patients infected with delta (49.8%) and alpha (28.6%) variants. Table S2 details the variant lineages at baseline for the Low and High-risk patients who participated in Phase 2 of the trial.

The median age of Low-risk patients was 35 years, 55.5% were female, 79% identified as white, 36% as Hispanic/Latinx and 19% as Black or African American. The mean BMI for these patients was 26.4, and the majority (74%) presented with mild COVID-19. Seropositivity rates at baseline were generally low among Low-risk patients and a numerically higher percentage of patients with seropositive status were placebo-treated (Table 1).

**Table 1:** Baseline characteristics among ambulatory patients at Low and High-risk of progressing to severe COVID-19.

|  | Double-Blinded, Low-risk |  |  | Open-Label, High-risk |  |  |
| --- | --- | --- | --- | --- | --- | --- |
|  | PBO<br>N=128 | BEB<br>N=125 | BEB+BAM<br>+ETE<br>N=127 | BEB<br>N = 100 | BEB+BAM<br>+ETE<br>N=50 | BEB+BAM<br>+ETE†<br>N=176 |
| Female, n (%) | 72 (56) | 63 (50) | 76 (60) | 52 (52) | 26 (52) | 98 (56) |
| Age, (median) | 34.0 | 34.0 | 37.0 | 48.5 | 52.5 | 51.0 |
| Age ≥65 years | 1 (1)<sup>‡</sup> | 0 | 0 | 17 (17) | 11 (22) | 35 (20) |
| Race or ethnic group, n (%) |  |  |  |  |  |  |
| Black or African American | 29 (24) | 19 (16) | 19 (16) | 14 (14) | 13 (28) | 28 (17) |
| Hispanic or Latinx | 45 (35) | 46 (37) | 45 (35) | 19 (19) | 8 (16) | 49 (28) |
| White | 90 (74) | 97 (83) | 99 (81) | 78 (78) | 32 (68) | 136 (80) |
| BMI, mean (SD) | 26.1 (4.3) | 27.5 (4.1) | 25.7 (4.6) | 34.5 (6.6) | 32.8 (7.4) | 32.7 (8.0) |
| SpO<sub>2</sub> Category<sup>1</sup>, n (%) |  |  |  |  |  |  |
| <96% | 14 (11) | 9 (7) | 10 (8) | 18 (18) | 15 (31) | 32 (18) |
| ≥96% | 114 (89) | 115 (93) | 117 (92) | 81 (82) | 34 (69) | 144 (82) |
| COVID-19 Disease Status, n (%) |  |  |  |  |  |  |
| Mild | 101 (79) | 84 (67) | 97 (76) | 74 (74) | 38 (76) | 129 (73) |
| Moderate | 27 (21) | 41 (33) | 30 (24) | 26 (26) | 12 (24) | 47 (27) |
| Duration of symptom onset to randomization, median days (range) | 3.5 (0-11) | 3.0 (1-11) | 3.0 (0-12) | 4.0 (1-13) | 4.0 (1-12) | 4.0 (1-13) |
| Baseline log viral load, mean | 6.0 | 6.4 | 6.0 | 5.7 | 5.2 | 6.5 |
| Baseline viral load Ct, mean | 25.10 | 23.63 | 25.09 | 26.03 | 27.64 | 23.44 |
| Seropositive at baseline, n (%)<sup>2</sup> | 18 (15) | 9 (8) | 12 (10) | 11 (11) | 8 (16) | 10 (6) |
| Vaccinated at baseline, n (%) | - | - | - | 22 (22) | 9 (18) | 55 (31) |

<sup>1</sup>SpO<sub>2</sub> data for three individuals not included. <sup>2</sup>Defined as the presence of anti-SARS-COV-2 nucleocapsid protein antibodies (total Ig) at baseline. Note: Vaccinated at baseline refers to at least partial vaccination. PBO, placebo; BAM, bamlanivimab; ETE, etesevimab; BEB, Bebtelovimab, 175 mg; BEB+BAM+ETE, bebtelovimab (175 mg) together with bamlanivimab (700 mg) and etesevimab (1400 mg). BEB+BAM+ETE †, High-risk patients enrolled according to updated CDC criteria; ‡, this was a protocol deviation since the age range for Low-risk patients was ≥18 to <65 years; N = number of patients in the analysis population; n = number of patients in the specified category; BMI, body mass index (kg/m<sup>2</sup>); SpO<sub>2</sub> = oxygen saturation.

Among patients in the three High-risk cohorts, median age ranged from 48.5-52.5 years, 52-56% were female, 68-80% identified as white, 16-28% as Hispanic/Latinx and 14-28% as Black of African American. The mean BMI was 32.7-34.5, and the majority of these patients (73-76%) presented with mild COVID-19. Vaccination levels and baseline-log viral load in High-risk patients were higher among those enrolled using the CDC updated High-risk criteria, who enrolled after the initial two High-risk treatment groups, reflecting the emergence of the delta variant in the United States. The majority of High-risk patients (74%) presented with mild COVID-19 (Table 1).

### Pharmacokinetic/Pharmacodynamic (PK/PD) modeling

The population PK dataset for BEB included 573 patients from the Phase 2 portion of the BLAZE-4 study who received either BEB or BEB+BAM+ETE, and blood samples were collected over an 85 day period after dosing. BEB serum concentration-time data were analysed using nonlinear mixed-effects modeling. The PK data were adequately described using a 2 compartment structural model. Parameter estimates of the model are reported in supplementary Table S3. Simulations using the PK model confirmed prior model predictions that a dose of 175 mg would result in adequate drug exposure above the serum IC_90_ (95% CI=1.43 to 1.92 µg/mL) through a 28-day period after a single dose (Figure 2).

**Figure 2:**
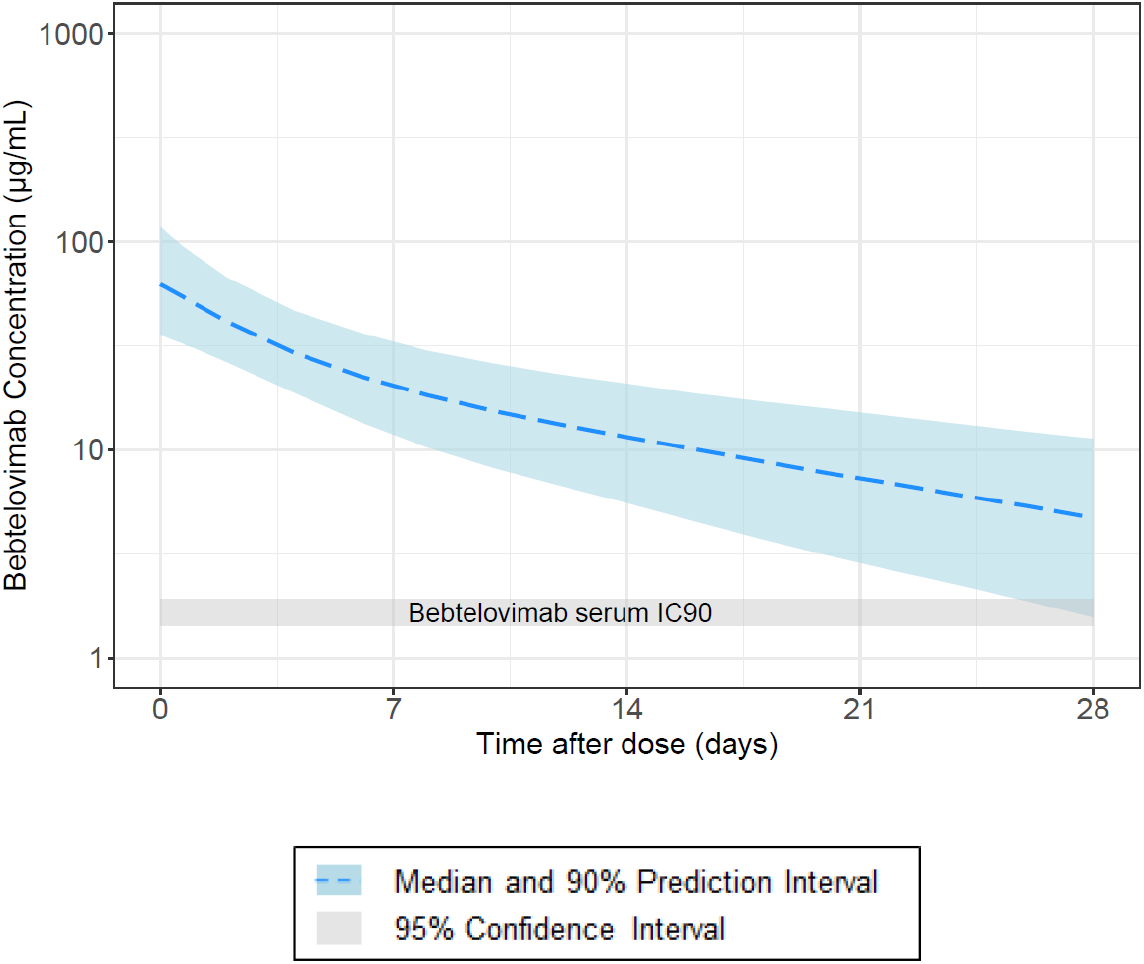
Bebtelovimab PK model-predicted concentration-time profile and derived in vivo IC_90_ following administration of 175 mg bebtelovimab. Abbreviation: IC_90_ = serum concentration that results in 90% of maximum inhibition.

**Figure 3:**
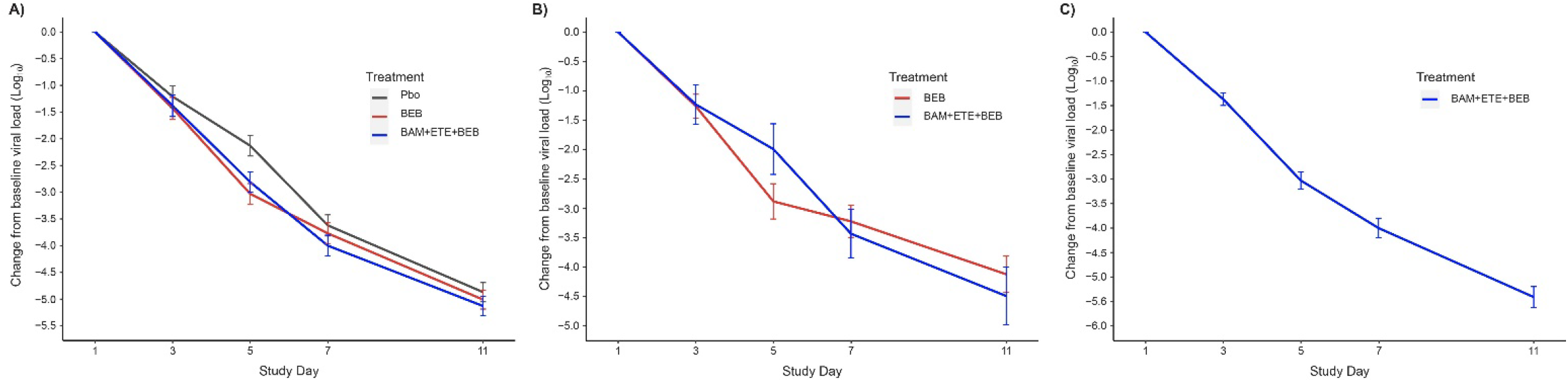
Viral Load Change from Baseline among patients receiving treatment with (A) BEB, BEB+BAM+ETE, or placebo in the placebo-controlled, Low-risk patients (B) BEB or BEB+BAM+ETE in open-label, High-risk patients, or (C) BEB+BAM+ETE in High-risk patients enrolled according to CDC updated criteria.

#### Viral dynamic modeling

Viral load measurements from 699 patients in the Phase 2 portion of the BLAZE-4 study were taken over a 29 day period, pooled together and analyzed using a previously reported viral dynamic model^23^. Based on this timecourse modeling approach, administration of BEB or BEB+BAM+ETE had a significant effect in increasing the elimination rate of the virus (i.e. nasopharyngeal viral RNA clearance) relative to placebo (p<0.0001). The increase in the elimination rate of the virus was estimated to result in a reduction in viral load area-under-the-curve of up to 88.3% over a 7 day period relative to placebo (supplementary Table S4), depending on how soon the drug is administered relative to the onset of symptoms.

There was no significant difference in viral load reduction between BEB administered alone or in combination with BAM+ETE. Covariate analysis showed that the delta variant was associated with higher viral load compared to those without the delta variant. However, this did not impact the drug effect (Figure S2). Pooling of all the viral load data from the Phase 2 portion of the BLAZE-4 study allowed evaluation of the impact of risk status on viral dynamics, in addition to other demographic factors such as age, bodyweight, and BMI. Risk status (Low vs. High) was not a significant covariate. Rather, age was a significant covariate, with older patients having slower viral clearance than younger patients. Supplementary Figure S3 shows a simulation done for a typical 21 year old and a 71 year old, representing the 5th and 95th percentiles of the age distribution in the current study.

The simulation in Figure S3 suggests that there may be a slightly greater reduction in viral load in the older patients because they stand to benefit more from drug administration instead of relying on their intrinsically slower rate of viral clearance. The pooled viral dynamic modeling approach that included testing of Low vs High risk status enabled bridging of the difference of drug treatment from placebo in Low-risk to the High-risk population, based on differences in age. Although the High-risk treatment arms did not include placebo, this analysis suggests that the difference in viral load between High-risk patients receiving BEB and those who would receive placebo would be at least similar to the difference seen in the Low-risk population.

### Efficacy

The results for the primary endpoint for the placebo-controlled, Low-risk patients revealed that active treatment arms had a numerically lower proportion of patients with PHVL at Day 7 (15/125 (12%) who received BEB; 16/126 (12.7%) who received BEB+BAM+ETE) compared with 25/126 (19.8%) who received placebo, but not at a level of statistical significance (p=0.097 BEB vs placebo; p=0.132 BEB+BAM+ETE vs placebo).

Pre-specified viral load endpoints were also examined. Viral load-area under the curve analysis from baseline to Day 11 showed statistically significant reductions for patients treated with BEB (mean (SD); 34.39 (17.12), p=0.006) and BEB+BAM+ETE (mean (SD); 32.41 (18.38), p=0.043) compared to placebo. A significant change in viral load from baseline was observed at Day 5 in Low-risk patients who received BEB compared to placebo (LSM (SE); -3.03 (0.19), p=<0.001) or BEB+BAM+ETE compared to placebo (LSM (SE); -2.81 (0.19), p=0.012) (Table 2). Statistical differences between BEB or BEB+BAM+ETE relative to placebo were not observed at other time points collected.

**Table 2:**
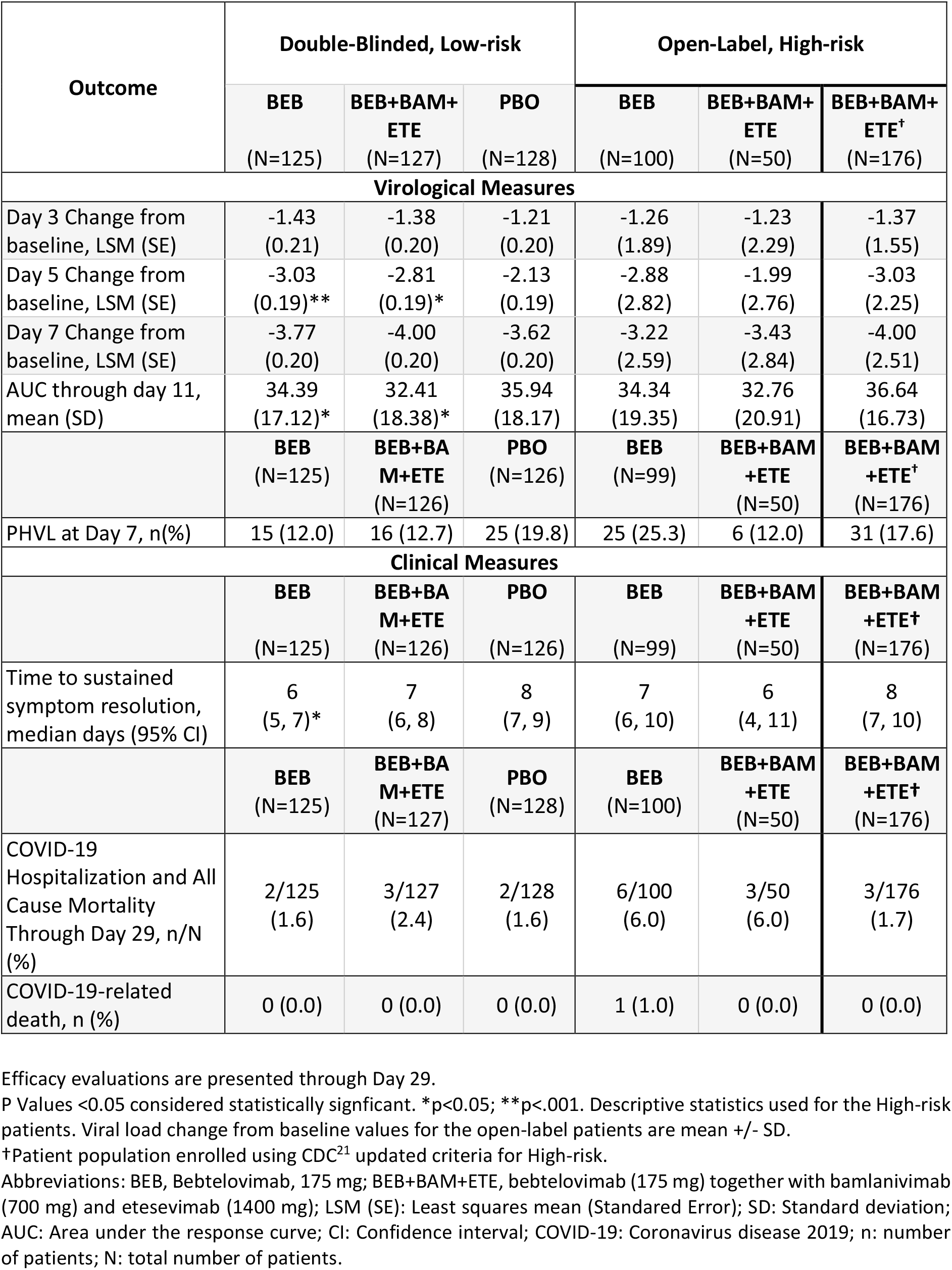
Viral Load Endpoints and Clinical Outcomes among Low and High-risk patients treated with bebtelovimab (BEB) alone or together with bamlanivimab and etesevimab (BEB+BAM+ETE)

The time to sustained symptom resolution was significantly decreased (p=0.003) among patients who received BEB alone (median days (95% CI); 6 (5,7)) relative to placebo (median days (95% CI); 8 (7,9)) (Figure 4). The time to sustained symptom resolution was also decreased among patients who received BEB+BAM+ETE (median days (95% CI); 7(6,8)) relative to placebo, albeit not at a level of statistical significance (p=0.289).

**Figure 4:**
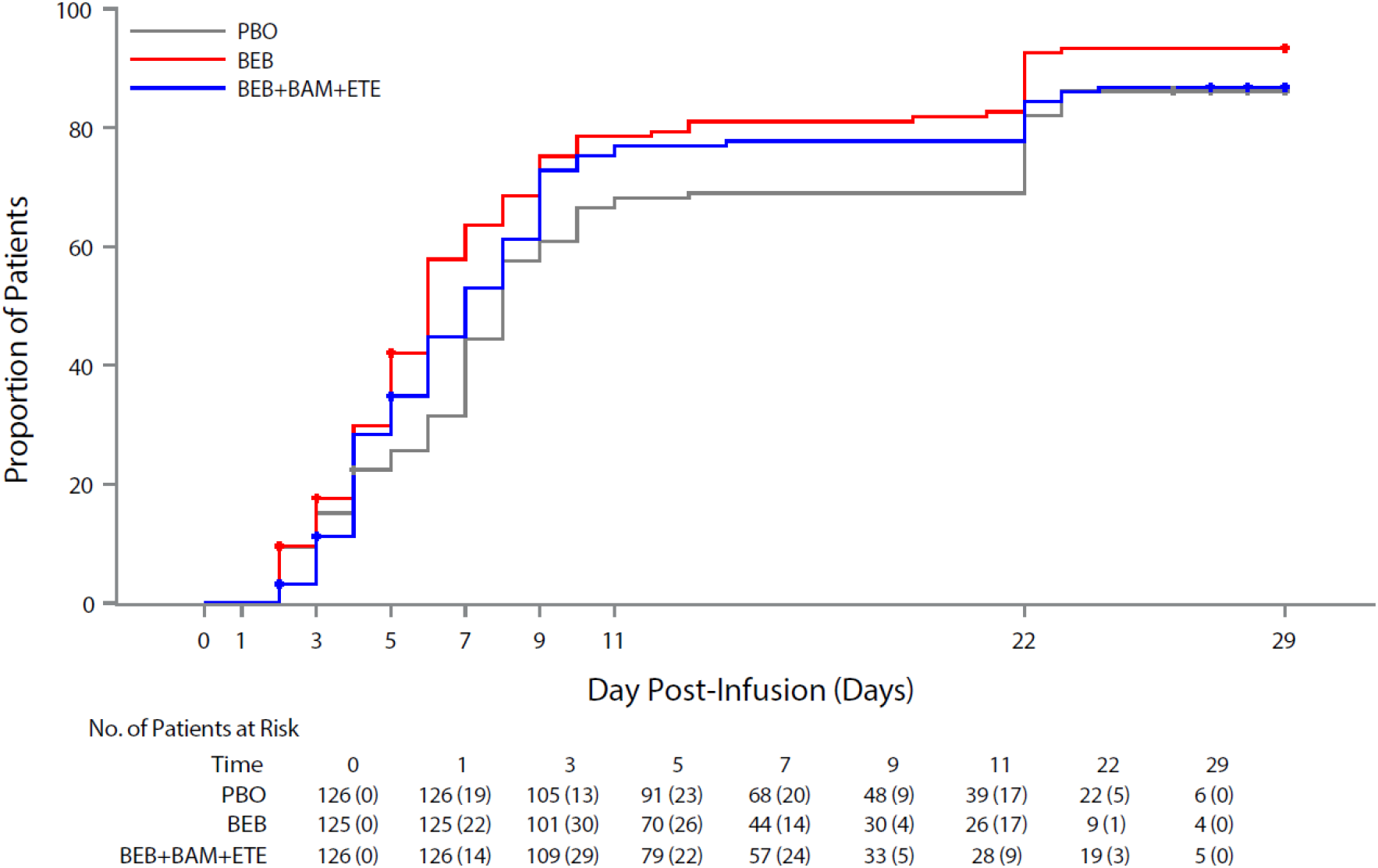
Kaplan–Meier Estimate of the Time to Sustained Symptom Resolution among Low-risk patients receiving bebtelovimab alone (BEB), bebtelovimab together with bamlanivimab and etesevimab (BEB+BAM+ETE) or placebo through Day 29. Patients at risk displayed under Day x are calculate based on patients whose time to event or censoring > Day x date. Number of events displayed under Day x are calculated based on events occurred during the time interval from Day x (excluding Week x date) to the day of next reported Day y (including Day y date). Abbreviations: BEB, Bebtelovimab, 175 mg; BEB+BAM+ETE, bebtelovimab (175 mg) together with bamlanivimab (700 mg) and etesevimab (1400 mg); PBO placebo.

The incidence of COVID-19-related hospitalization or all-cause deaths by day 29 were similar within the Low-risk population (Table 2). One death due to COVID-19 (study disease) on Day 5 was reported in a patient who received BEB+BAM+ETE.

### Safety

The safety profiles observed with BEB alone and together with BAM+ETE (BEB+BAM+ETE) were similar. In the Phase 1 portion there were no deaths, SAEs, or discontinuations due to AEs (supplementary Table S5). The safety profiles observed with increased dose levels and increased infusion rates were consistent with those reported with BAM and BAM+ETE in prior studies^9,10,13,14,26^.

In the Phase 2 portion there were no discontinuations due to TEAEs among the Low-risk patients (Table 3). There was one death at Study Day 5 due to COVID-19 in a High-risk patient who received BEB+BAM+ETE. This was not reported as an AE per protocol defined criteria (Section 10.3.2 in [protocol/supplementary appendix]). There was one SAE of abortion spontaneous at Study Day 59 in the BEB arm. All other TEAEs were mild or moderate in severity, and the overall frequencies of TEAEs were comparable between the BEB (8.8%) and BEB+BAM+ETE groups (12.6%), and placebo (7.8%).

**Table 3:** Adverse Events.

| Safety | Double-Blinded, Low-risk |  |  | Open-Label, High-risk |  |  |
| --- | --- | --- | --- | --- | --- | --- |
|  | BEB<br>(N=125) | BEB+BAM+ETE<br>(N=127) | PBO<br>(N=128) | BEB<br>(N=100) | BEB+BAM+ETE<br>(N=50) | BEB+BAM+ETE†<br>(N=176) |
| **TEAEs, n (%)** |  |  |  |  |  |  |
| Mild | 4 (3.2) | 10 (7.9) | 6 (4.7) | 10 (10.0) | 3 (6.0) | 9 (5.1) |
| Moderate | 7 (5.6) | 5 (3.9) | 4 (3.1) | 8 (8.0) | 4 (8.0) | 7 (4.0) |
| Severe | 0 | 1 (0.8) | 0 | 2 (2.0) | 1 (2.0) | 4 (2.3) |
| **SAEs, n/N (%)** | 0 | 1/127 (0.8) | 0 | 3/100 (3.0) | 2/50 (4.0) | 2/176 (1.1) |
| **Discontinuation from treatment due to AE, n/N (%)** | 0 | 0 | 0 | 1/100 (1.0)‡ | 0 | 0 |
| **Infusion Related Reactions, n** | 0 | 0 | 0 | 0 | 1/50 (2.0) | 1/176 (0.6) |
| **Anaphylaxis, n** | 0 | 0 | 0 | 0 | 0 | 0 |

Adverse Events are presented through Day 85. Descriptive statistics used for all safety outcomes
†Patient patients enrolled using CDC expanded criteria for High-risk.‡Discontinued from treatment due to death. Abbreviations: Adverse Event: AE; n: number of patients; N: total number of patients; TEAEs: Treatment emergent AEs; SAEs: Serious AEs.

**Table 4:** Authentic SARS-CoV-2 Cytopathic Effect Inhibition of Spike Variants by Bebtelovimab (BEB), Bamlanivimab with Etesevimab (BAM+ETE), and Bebtelovimab together with Bamlanivimab with Etesevimab (BEB+BAM+ETE).

| Antibody | WA1 Isolate | Delta (B.1.617.2) |  | Omicron (B.1.1.529/BA.1) |  |
| --- | --- | --- | --- | --- | --- |
|  | IC<sub>99</sub> ng/mL<sup>a</sup> | IC<sub>99</sub> ng/mL<sup>a</sup> | Fold Change<sup>b</sup> | IC<sub>99</sub> ng/mL<sup>a</sup> | Fold Change<sup>b</sup> |
| BEB | 3.66 | 4.88 | 1.3 | <2.44<sup>c</sup> | -1.5<sup>c</sup> |
| BAM+ETE (1:2 molar ratio) | 78.13 | 19.53 | -4 | >10,000 | >128 |
| BEB+BAM+ETE (1:4:8) | 7.32 | 4.88 | -1.5 | 9.77 | 1.3 |

Abbreviations: BAM+ETE = bamlanivimab + etesevimab (1:2 molar ratio); BEB = bebtelovimab; IC<sub>99</sub> = concentration inhibiting maximal activity by 99%; SARS-CoV-2 = severe acute respiratory syndrome coronavirus 2; WA = Washington.
- <sup>a</sup> IC<sub>99</sub> values presented are a result of a single experiment with technical replicates. - <sup>b</sup> Fold shifts are calculated by comparing IC<sub>99</sub> of variant to the WA1 strain control. - <sup>c</sup> Values of <2.44 were given when an antibody showed full neutralization even at the lowest concentration tested, fold shifts for these values were calculated using 2.44 as the IC<sub>99</sub>.

In the High-risk patients, there was one SAE of death due to cerebrovascular accident in the BEB arm. This was the only TEAE that prevented study completion in the High-risk patients. Additional SAEs of meniscus injury and pulmonary embolism were reported in the BEB arm, and femur fracture, lower limb fracture, psychotic disorder, and osteomyelitis in the BEB+BAM+ETE arms.

The majority of TEAEs were mild or moderate in severity among Low-risk (n=37/380, 9.7%) and High-risk patients (n=48/326, 14.7%). Two infusion related-reactions (IRRs) were observed in patients receving BEB+BAM+ETE, both within one minute of infusion. Both resolved upon interruption or withdrawal of infusion. No IRRs were observed in patients receiving BEB and there were no reports of anaphylaxis. Overall, the safety profiles observed in the Phase 1 and Phase 2 portions of the study with BEB alone or BEB+BAM+ETE were consistent with those reported in studies of other anti-SARS-CoV-2 neutralizing monoclonal antibodies^15,16^.

### SARS-CoV-2 Virus Neutralization Breadth of Coverage

To evaluate the breadth of neutralization coverage, we used an endpoint neutralization assay that measures multiple viral lifecycle stages (viral entry and spread) and provides the antibody concentration that is necessary for complete inhibition, which is equivalent to an IC_99_ value. BEB, BAM+ETE, and BEB+BAM+ETE all neutralized the ancestral WA1 SARS-CoV-2 isolate and the delta SARS-CoV-2 isolate (B.1.617.2) with similar potencies. When evaluating the omicron SARS-CoV-2 isolate (B.1.1.529/BA.1), BAM+ETE showed limited to no neutralization of the omicron viral isolate, whereas BEB potently neutralized the omicron isolate with a <2.44 ng/ml estimated IC_99_, showing similar or greater potency against omicron as that of delta and WA1 isolates. These results align with the activity patterns observed with the omicron pseudovirus assessment^20,27^.

## Discussion and Conclusions

To counteract future variants of SARS-CoV-2 that could render clinically available neutralizing mAbs ineffective, the discovery and characterization of increasingly more potent mAbs continue. Bebtelovimab, derived from an antibody from a convalscent patient, was initially identified in April 2020. Importantly, BEB binds a highly conserved region of the RBD of the S protein with high potency^20^. In vitro neutralization assays using dozens of pseudoviruses encoding potential and emerging viral variants of interest confirmed its broad neutralization capability^20^. Further, whole virus neutralization assays, including the results provided herein of BEB neutralization of the omicron variant, confirm earlier pseudovirus findings^20^.

With the initial identification of emerging viral variants that were resistant to BAM+ETE, clinical development for BEB was initiated in January 2021. Non-clinical data indicated BEB should be effective against all currently known viral variants of interest or concern, including the delta and omicron lineages. Proactive communications with regulatory authorities and streamlined clinical design allowed for sequential Phase 1 and Phase 2 trials. In parallel, manufacturing and product delivery functions streamlined efforts and conducted stability studies on representative development batches to enable dating of FHD material in coordination with regulatory authorities. Batch release testing was also rapidly transferred to an external testing partner which was enabled by earlier experience with BAM+ETE, and the formulation for BEB was designed simliar to BAM+ETE to ensure compatibility with combination therapies. Phase 1 studies also utilized sterile mobile units (fully integrated mobile central processing facility for sterile manufacturing) to enable rapid small drug product batch production and expedite study initiation. Collectively, these efforts enabled FHD in April 2021.

A key element ensuring rapid clinical trial results was studying a single dose level of BEB. This ultimately allowed the trial to be completed as soon as possible in order to meet the urgent need of a new effective drug during the pandemic. The phase 2 single-dose level of BEB (175 mg) was selected using PK/PD modeling which leveraged results from multiple BAM+ETE studies. The PK/PD modeling approach was deemed reliable because the approach had been previously implemented to predict the efficacious doses of BAM+ETE, before any clinical trial data was available^17^. The clinical trial data later confirmed the model predictions of 700 mg BAM and 1400 mg ETE resulting in maximum drug effect; the PK/PD modeling approach described in Chigutsa et al., 2022^17^ was further strengthened and refined through incorporating the BAM+ETE clinical trial data to update the model^24^. With the modeling platform now established and validated, the objective was to replace the potency (IC_90_) of the drug in question in the model with that of the new drug in development (BEB) and determine the dose that would result in concentrations above IC_90_ up to a 28 day period and a maximum reduction in viral load.

Because there were a number of effective and authorized mAbs to SARS-CoV-2 for patients at increased risk for severe disease at the time of study design for BLAZE-4 (BAM, BAM+ETE, and casirivimab together with imdevimab (REGN-COV2, Regeneron had been authorized in the US by mid-February 2021^29^), a placebo-controlled study composed of High-risk patients would not have been appropriate for ethical reasons. Therefore, PHVL at Day 7 was identified as a feature predictive of progression to severe COVID-19^10^ and was designated the primary outcome of the placebo-controlled Low-risk population. Amongst patients who were hospitalized with COVID-19 or had died from any cause by Day 29 in the Phase 2/3 BLAZE-1 randomized controlled trial, 69% (9 of 13 patients) of the Phase 2 population (composed of Low- and High-risk patients) and 69% (45 of 65 patients) of the Phase 3 population (composed solely of High-risk patients) had PHVL at Day 7. In patients who were never hospitalized, only 13% (74 of 564 patients) and 23% (403 of 1733 patients) had PHVL at Day 7, respectively. The initial identification of PHVL as a potential surrogate for clinical outcomes predated the emergence of delta and omicron vairiants which have different viral dynamics compared to earlier variants of SARS-CoV-2, which adds uncertainty around the threshold of >5.27 at day 7 defining PHVL.

Both active treatment arms in the Low-risk patients did not achieve statistical separation in the proportion of patients with PHVL at Day 7 (15/125 who received BEB (12%) and 16/126 who received BEB+BAM+ETE (12.7%) compared with 25/126 (19.8%) who received placebo). Based on previous data collected in Low-risk patients, the sample size was determined with an assumed PHVL rate of 28% in the placebo arm; however, the observed PHVL rate with Low-risk patients on placebo was noticably less than predicted. This ultimately reduced the power of the study to test statistical separation. Actual reasons for the lower than expected placebo response are unclear but could have been influenced by the previous PHVL data being based on Low-risk patients enrolled in the winter of 2020/2021 when ancestrial strains of SARS-CoV-2 were predominant in the United States. This data set was appropriate when the Phase 2 portion of the BLAZE-4 study was designed in early 2021, however, during the conduct of this trial, several emerging viral variants were observed in our patients (including the alpha, gamma, delta, and mu lineages), which could have complicated our estimation of PHVL rates during study design. Additionally, there were more seropositive patients at baseline in the placebo arm than the active treatment arms, and seropositivity was associated with lower levels of patients with PHVL at Day 7 (data not shown). Finally, enrollment in this study was generally in a younger population with lower BMI and lower mean viral load than previous trials, which may have resulted in faster, unassisted viral clearance for the overall population due to more robust endogenous antibody responses.

Nonetheless, pre-specified secondary outcomes measuring changes in viral load indicate BEB and BEB+BAM+ETE treatments lead to rapid viral clearance relative to placebo. Both active treatment arms were statistically separated from placebo at Day 5 in change from viral baseline as well as AUC analysis of viral load out to Day 11 of treatment. Generally, the kinetics of viral clearance between BEB and BEB+BAB+ETE were similar amongst Low- and High-risk patients. In line with non-clinical data, BEB treatment was also associated with numerically greater viral load reduction compared to placebo in patients infected with the alpha and delta variants; other variants of concern and interest were noted in the efficacy population, but not sufficiently well-represented to make conclusions on the potential benefit of active treatment. There is a known relationship between SARS-CoV-2 RNA copy number and severity of symptoms in outpatients; this was exemplified in that we observed a reduction in the median time to sustained symptom resolution in patients treated with BEB (2 day reduction) and BEB+BAM+ETE (1 day reduction).

In all patients receiving active treatment, TEAEs were mild to moderate in severity and similar across treatment groups. Two mild, nonserious IRRs were reported, and patients recovered quickly with either interruption or withdrawal of study drug. No anaphylaxis was reported. Overall, the safety data was consistent with other clinical trials investigating neutralizing mAbs to SARS-CoV-2.

In the Phase 1 portion of the study, faster infusion rates of BEB or BEB+BAM+ETE were not associated with an increasing number and/or severity of TEAEs. Informed by this data, BEB was administered to patients at High-risk for severe disease with an IV injection of at least 30 seconds while BEB+BAM+ETE was administered over at least 6.5 minutes. Both administration rates are markedly faster than previously authorized mAbs for COVID-19 patients in the clinic (21-60 minute infusions). Considering BEB can be administered over at least 30 seconds, IV push of the medication saves considerable time relative to the preparation and administration of currently authorized mAbs and may facilitate greater treatment capacity throughout infusion centers.

There are several limitations to this study. The study limited recruitment exclusively to the United States. As noted previously, placebo-controlled data was limited to patients at Low risk for severe disease. Thus, we could not demonstrate statistically significant differences in viral clearance or rapid resolution of symptoms in patients at increased risk for severe disease between two active therapies at a time when circulating variants largely remained susceptible to BAM+ETE. This proof-of-concept study was also not powered to examine improvements in clinical outcomes in those with active treatment; however, the proportion of patients with COVID-19 related hospitalization and all cause mortality are in line with similar observations with other mAbs to SARS-CoV-2. Finally, this study was conducted over the spring and summer of 2021, prior to the emergence of omicron in November 2021. There are no clinical data evaluating the clinical efficacy of BEB in patients infected with omicron; however, non-clinical pseudovirus data with all sublineages of omicron (including BA.2) and confirmatory authentic omicron SARS-CoV-2 virus neutralization assays presented here indicate that BEB is effective in neutralizing omicron^20^. Moreover, this same limitation would be applicable to any EUA product studied at an earlier phase of the pandemic, and for which surrogates of antiviral efficacy would also be required.

The BLAZE-4 study enrolled its last patient in July 2021. At that time, the delta variant, which was identified in November 2020 and emerged in the United States in March 2021, was the predominant circulating variant world-wide. In vitro neutralization assays confirmed that BAM+ETE and other authorized neutralizing mAbs were effective against this viral lineage, negating the need to seek authorization for BEB or BEB+BAM+ETE at that time. However, the rapid emergence of the omicron variant starting in late November 2021 and in vitro neutralization assays indicating that sotrovimab was the only authorized mAb to retain activity against omicron, led to the initiation of regulatory activities seeking authorization of BEB in the United States. By early January 2022, an intial request for EUA of BEB was submitted. In parallel and to meet regulatory expectations and expected clinical demand, key information for the EUA was submitted on a rolling basis. On February 12th, 2022, BEB received EUA for the treatment of patients at increased risk for progression to severe COVID-19 based on the data provided from in vitro neutralization assays and clinical results from the BLAZE-4 study. During the rolling EUA submission, stability studies were rapidly restarted and concluded, remaining manufacturing testing was completed to provide additional drug product manufacturing capacity, and efforts were made to streamline labels and package vials. Collectively these efforts enabled an additional 100,000 doses of drug product to be available in early 2022 than was intially projected. The EUA and acceleration of a new available therapeutic was made possible by continuous drug discovery, streamlined and innovative approaches in clinical trial design, manufacturing and drug product delivery, and close engagement with regulatory authorities with the impetus of a clear unmet medical need during an evolving pandemic.

## Supporting information

Redacted PYAH Protocol

Redacted PYAH Protocol Addenda (4)

## Data Availability

Lilly provides access to all individual participant data collected during the trial, after anonymization, with the exception of pharmacokinetic or genetic data. Data are available to request 6 months after the indication studied has been approved in the US and EU and after primary publication acceptance, whichever is later. No expiration date of data requests is currently set once data are made available. Access is provided after a proposal has been approved by an independent review committee identified for this purpose and after receipt of a signed data sharing agreement. Data and documents, including the study protocol, statistical analysis plan, clinical study report, blank or annotated case report forms, will be provided in a secure data sharing environment. For details on submitting a request, see the instructions provided at www.vivli.org.

## Acknowledgements

We thank the BLAZE-4 investigators and their support staff for their dedication and hard work throughout this program. Authentic virus assays were performed independently by investigators and support staff at the University of Maryland as part of the US government COVID-19 therapeutics response efforts. We thank Timothy R. Holzer, Ph.D, Philip J. Ebert, Ph.D, Richard E. Higgs, Ph.D, John Calley, Ph.D, Gary Heady, Ph.D, Ling Zhang, Ph.D, Angie Fulford, PhD., Drew Nedderman, Ph.D, Erin Wray, Ph.D, and Leslie O’Neil Reising, MS, of Eli Lilly and Company for assistance with viral sequencing for the Blaze-4 study. We thank Honglu Liu, MSc and the PYAH statistical analyst team of Eli Lilly and Company for their support with statistical analyses. We thank Lynn Naughton, Ph.D of Eli Lilly and Company for providing medical writing and editorial support throughout the manuscript. We thank Carmen Deveau, Ph.D, Adam Clooney, Ph.D, and Natalie Haustrup, Ph.D of Eli Lilly and Company for editorial and process support throughout preparation of the manuscript.

