## Supplementary for "Bebtelovimab, alone or together with bamlanivimab and etesevimab, as a broadly neutralizing monoclonal antibody treatment for mild to moderate, ambulatory COVID-19"

**Supplementary Material**

Inclusion and Exclusion Criteria pg 2

**Figure S1:** Phase 1 Study Design pg 5

**Table S1:** Phase 1 Patient Demographics and Baseline Characteristics pg 6

**Table S2:** Phase 2 Variant Lineages at Baseline pg 7

**Table S3:** Bebtelovimab Population Pharmacokinetic Model Parameters pg 8

**Table S4:** Model-Estimated Reduction in Viral Load Following a Single IV Dose of 175 mg Bebtelovimab Relative to Placebo Depending on the Time of Treatment pg 9

**Figure S2:** Model-Estimated Typical Viral Load Profiles Showing Impact of Infection with the Delta Variant. pg 10

**Figure S3:** Model-Estimated Typical Viral Load Profiles Showing Impact of Age on Viral Dynamics. pg 11

**Table S5:** Phase 1: Overview of Adverse Events following IV Infusion pg 12

**Inclusion and Exclusion Criteria**

Phase 1 substudy and placebo-controlled Low-risk population

| **Inclusion Criteria** | **Exclusion Criteria** |
|---|---|
| <u>Age</u><br>• ≥18 and <65 years of age at the time of randomization | <u>Medical Conditions</u><br>• BMI ≥35<br>• SpO2 ≤93% on room air at sea level or PaO2/FiO2 <300, respiratory rate ≥30 per minute, heart rate ≥125 per minute<br>• Require mechanical ventilation or anticipated impending need for mechanical ventilation |
| <u>Disease Characteristics</u><br>• Do not have a risk factor defined for High-risk population (for placebo-controlled Low-risk population only)<br>• Currently not hospitalized<br>• Have one or more mild or moderate COVID-19 symptoms<br> ○ Fever<br> ○ Cough<br> ○ Sore throat<br> ○ Malaise<br> ○ Headache<br> ○ Muscle pain<br> ○ Gastrointestinal symptoms, or<br> ○ Shortness of breath with exertion<br> ○ Nasal congestion or runny nose<br> ○ New loss of smell<br> ○ Chills<br>• Must have sample collection for first positive SARS-CoV-2 viral infection determination ≤3 days prior to start of the infusion | <u>Other Exclusions</u><br>• History of a positive SARS-CoV-2 serology test<br>• History of a positive SARS-CoV-2 test prior to the 1 serving as eligibility for this study<br>• Received any treatment, vaccine, or intervention for SARS-CoV-2 |

Abbreviations: BMI = body mass index; COVID-19 = coronavirus disease 2019; FiO2 = fraction of inspired oxygen in the air; PaO2 = partial pressure of oxygen; SARS-CoV-2 = severe acute respiratory syndrome coronavirus 2; SpO2 = saturation of peripheral oxygen.

Open-label, High-Risk population

| **Inclusion Criteria** | **Exclusion Criteria** |
|---|---|
| Age and Risk Factors<br>• ≥18 years of age and have at least 1 of the following risk factors<br> ○ Are ≥65 years of age<br> ○ Have a BMI ≥35<br> ○ Have chronic kidney disease<br> ○ Have type 1 or type 2 diabetes<br> ○ Have immunosuppressive disease<br> ○ Are currently receiving immunosuppressive treatment, or<br> ○ Are ≥55 years of age AND have<br> ○ cardiovascular disease, OR<br> ○ hypertension, OR<br> ○ chronic obstructive pulmonary disease or other chronic respiratory disease<br>• 12 to 17 years of age and have at least 1 of the following risk factors<br> ○ Have a BMI ≥85th percentile for their age and gender based on CDC growth charts<br> ○ Have sickle cell disease<br> ○ Have congenital or acquired heart disease<br> ○ Have neurodevelopmental disorders,<br> ○ Have a medical-related technological dependence<br> ○ Have asthma or reactive airway or other chronic respiratory disease that requires daily medication for control<br> ○ Have type 1 or type 2 diabetes<br> ○ Have chronic kidney disease<br> ○ Have immunosuppressive disease, or<br> ○ Are currently receiving immunosuppressive treatment. | Medical Conditions<br>• SpO2 ≤93% on room air at sea level or PaO2/FiO2 <300, respiratory rate ≥30 per minute, heart rate ≥125 per minute<br>• Require mechanical ventilation or anticipated impending need for mechanical ventilation |

<table>
<tr>
<td>
<u>Age and Risk Factors (enrolled according to updated CDC criteria)</u>
<ul>
<li>≥12 years of age and have at least 1 of the following risk factors
<ul>
<li>Are ≥65 years of age</li>
<li>Are adults (≥18 years of age) with BMI >25 kg/m2, or if age 12-17, have BMI ≥85th percentile for their age and gender based on CDC growth charts</li>
<li>Have chronic kidney disease</li>
<li>Have type 1 or type 2 diabetes</li>
<li>Have immunosuppressive disease</li>
<li>Are currently receiving immunosuppressive treatment</li>
<li>Have cardiovascular disease (including congenital heart disease) or hypertension</li>
<li>Have chronic lung diseases</li>
<li>Have sickle cell disease</li>
<li>Have a neurodevelopmental disorder or other conditions that confer medical complexity, or</li>
<li>Have a medical-related technological dependence</li>
</ul>
</li>
</ul>
</td>
<td></td>
</tr>
<tr>
<td>
<u>Disease Characteristics</u>
<ul>
<li>Currently not hospitalized</li>
<li>Have one or more mild or moderate COVID-19 symptoms
<ul>
<li>Fever</li>
<li>Cough</li>
<li>Sore throat</li>
<li>Malaise</li>
<li>Headache</li>
<li>Muscle pain</li>
<li>Gastrointestinal symptoms, or</li>
<li>Shortness of breath with exertion</li>
<li>Nasal congestion or runny nose</li>
<li>New loss of smell</li>
<li>Chills</li>
</ul>
</li>
<li>Must have sample collection for first positive SARS-CoV-2 viral infection</li>
</ul>
</td>
<td>
<u>Other Exclusions</u>
<ul>
<li>History of a positive SARS-CoV-2 serology test</li>
<li>History of a positive SARS-CoV-2 test prior to the 1 serving as eligibility for this study</li>
<li>Received any treatment or investigational intervention for SARS-CoV-2 (not including SARS-CoV-2 vaccine)</li>
</ul>
</td>
</tr>
</table>

| |
|---|
| determination ≤3 days prior to start of the infusion |

Abbreviations: BMI = body mass index; COVID-19 = coronavirus disease 2019; FiO2 = fraction of inspired oxygen in the air; PaO2 = partial pressure of oxygen; SARS-CoV-2 = severe acute respiratory syndrome coronavirus 2; SpO2 = saturation of peripheral oxygen.

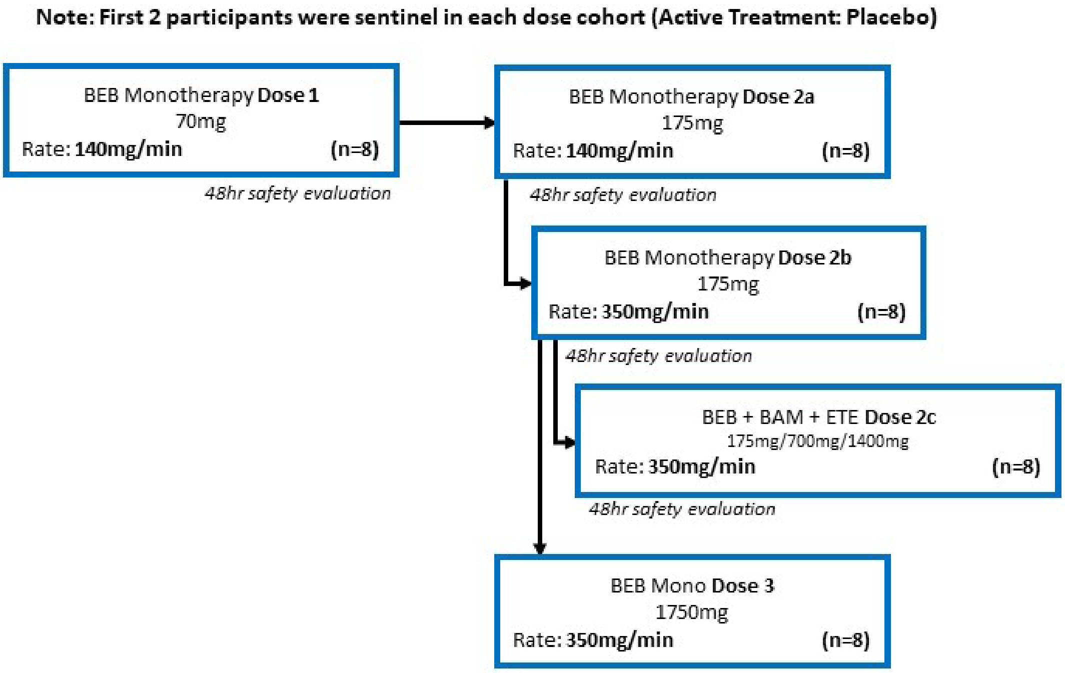

**Figure S1:** Phase 1 Study Design. Abbreviations: BAM = bamlanivimab; BEB = bebtelovimab; ETE = etesevimab; hr = hour; mg = milligrams; min = minutes; n = number of participants**. Note:** The first 2 participants in each cohort were sentinel (BEB:placebo). Subsequent participants were randomized to the remaining treatment allocations, 5 to BEB or BEB+BAM+ETE, and 1 to placebo.

**Table S1:** BLAZE-4 Phase I: Demographics and Baseline Characteristics of Ambulatory Patients with COVID-19 Administered mAb Treatment via Intravenous Infusion

| Dose | PBO | BEB<br>70 mg | BEB<br>175 mg | BEB<br>175 mg | BEB+BAM +ETE<br>175+700+ 1400 mg | BEB<br>1750 mg | Total |
|---|---|---|---|---|---|---|---|
| **Rate** | | 140 mg / min | 140 mg / min | 350 mg / min | 350 mg / min | 350 mg / min | |
| **Total (N)** | N=10 | N=6 | N=6 | N=6 | N=6 | N=6 | N=40 |
| **Female,** n (%) | 3 (30) | 3 (50) | 6 (100) | 1 (17) | 3 (50) | 5 (83) | 21 (53) |
| **Age,** (median) | 51.0 | 33.5 | 50.0 | 29.5 | 41.5 | 37.5 | 41.5 |
| **Race or ethnic group,** n (%) | | | | | | | |
| Black or African American | 1 (10) | 0 | 0 | 0 | 0 | 0 | 1 (3) |
| Hispanic or Latino | 3 (30) | 0 | 0 | 1 (17) | 1 (17) | 2 (33) | 7 (18) |
| White | 9 (90) | 6 (100) | 6 (100) | 6 (100) | 6 (100) | 6 (100) | 39 (98) |
| **BMI,** (mean (SD)) | 30.7 (7.0) | 28.1 (2.3) | 25.8 (4.6) | 26.7 (5.3) | 26.4 (5.0) | 25.5 (5.3) | 27.6 (5.4) |
| **$SpO_2$ Category,** n (%) | | | | | | | |
| <96% | 2 (20) | 0 | 0 | 0 | 1 (17) | 1 (17) | 4 (10) |
| ≥96% | 8 (80) | 6 (100) | 6 (100) | 6 (100) | 5 (83) | 5 (83) | 36 (90) |
| **COVID-19 Disease Status**, n (%) | | | | | | | |
| Mild | 10 (100) | 5 (83) | 5 (83) | 6 (100) | 4 (67) | 5 (83) | 35 (88) |
| Moderate | 0 | 1 (17) | 1 (17) | 0 | 2 (33) | 1 (17) | 5 (13) |
| **Duration of symptom onset to randomization**, Median (range) | 3.0 (0-5) | 2.0 (1–4) | 3.5 (0-7) | 3.5 (3-7) | 3.5 (2-7) | 4.0 (2-5) | 3.0 (0-7) |
| **Baseline viral load,** mean | 6.9 | 7.3 | 5.9 | 6.9 | 6.7 | 6.3 | 6.7 |
| **Seropositive at baseline,** n (%) | 2 (22) | 1 (17) | 0 | 0 | 1 (17) | 1 (17) | 5 (14) |

**Abbreviations:** BAM, bamlanivimab; BEB, bebtelovimab; BMI, body mass index (kg $m^2$); COVID-19 = coronavirus disease 2019; ETE, etesevimab; kg, kilograms; m, meters; mAb, monoclonal antibody;

mg, milligrams; min, minutes; N, number of patients in the analysis population; n, number of patients in the specified category; PBO, pooled placebo; SD, standard deviation; $SpO_2$, oxygen saturation

**Table S2.** Variant Lineages at Baseline_BLAZE-4 Phase 2 Low and High-Risk Patient Cohorts.

| | **Placebo** | **BEB** | **BEB+BAM+ETE** | **Total** |
|---|---|---|---|---|
| **Alpha** | 23.1% (25/108) | 41.8% (84/201) | 21.9% (66/302) | 28.6% (175/611) |
| **Beta** | 0 (0/108) | 0.5% (1/201) | 0.7% (2/302) | 0.5% (3/611) |
| **Gamma** | 6.5% (7/108) | 5.0% (10/201) | 5.6% (17/302) | 5.6% (34/611) |
| **Delta** | 60.2% (65/108) | 31.3% (63/201) | 58.3% (176/302) | 49.8% (304/611) |
| **Delta + K417N** | 0 (0/108) | 0.5% (1/201) | 1.3% (4/302) | 0.8% (5/611) |
| **Iota** | 0.9% (1/108) | 1.5% (3/201) | 0 (0/302) | 0.7% (4/611) |
| **Lambda** | 0 (0/108) | 0.5% (1/201) | 0.7% (2/302) | 0.5% (3/611) |
| **Mu** | 2.8% (3/108) | 5.0% (10/201) | 3.3% (10/302) | 3.8% (23/611) |
| **Non-WHO classified** | 1.9% (2/108) | 4.5% (9/201) | 2.3% (7/302) | 2.9% (18/611) |
| **Not determined** | 4.6% (5/108) | 9.5% (19/201) | 6.0% (18/302) | 6.9% (42/611) |

Abbreviations: BAM = bamlanivimab; BEB = bebtelovimab; ETE = etesevimab; SARS-CoV-2 = severe acute respiratory syndrome coronavirus 2; WHO = World Health Organization.

**Table S3:** Bebtelovimab Population Pharmacokinetic Model Parameters

| Parameter Description | Population Estimate | %SEE | Interindividual Variability | %SEE |
|---|---|---|---|---|
| CL (L/d) | 0.29 | 1.452 | 33.4 | 5.92 |
| Q (L/d) | 0.248 | 1.847 | -- | -- |
| Effect[a] of body weight on CL and Q | 0.81 | (Fixed) | -- | -- |
| V1 (L) | 2.45 | 1.424 | 30.5 | 7.25 |
| V2 (L) | 1.37 | 1.453 | -- | -- |
| Effect[b] of body weight on V1 and | 1 | (Fixed) | -- | -- |
| Residual Error (proportional) | 0.188 | 2.037 | 87.1 | 7.22 |

Abbreviations: CL = systemic clearance; Q = intercompartmental clearance; SEE = standard error of the estimate; V1 = central compartment volume of distribution; V2 = peripheral volume of distribution.

[a]Parameter = Population Estimate × (body weight/70)$^{0.81}$. (Exponent from [Betts et al. 2018[1]])

[b]Parameter = Population Estimate × (body weight/70).

**Table S4:** Model-Estimated Reduction in Viral Load Following a Single IV Dose of 175 mg Bebtelovimab Relative to Placebo Depending on the Time of Treatment

| Bebtelovimab administration relative to symptoms onset (Days) | Reduction in time-weighted average $\log_{10}$ viral load from Days 0 to 7 relative to placebo (% absolute decrease)[a] |
|---|---|
| 1 (Delta positive) | 0.93 (88.3%) |
| 1 (Delta negative) | 0.87 (86.5%) |
| 4 (Delta positive) | 0.80 (84.2%) |
| 4 (Delta negative) | 0.55 (71.8%) |
| 10 (Delta positive) | 0.31 (51.0%) |
| 10 (Delta negative) | 0.09 (18.7%) |

Abbreviations: IV = intravenous; mg = milligrams.

[a] % absolute reduction is calculated from the $\log_{10}$ values according to the equation below: $P = (1 - 10^{-L}) \times 100$, where P is the percent absolute decrease, and L is the $\log_{10}$ reduction.

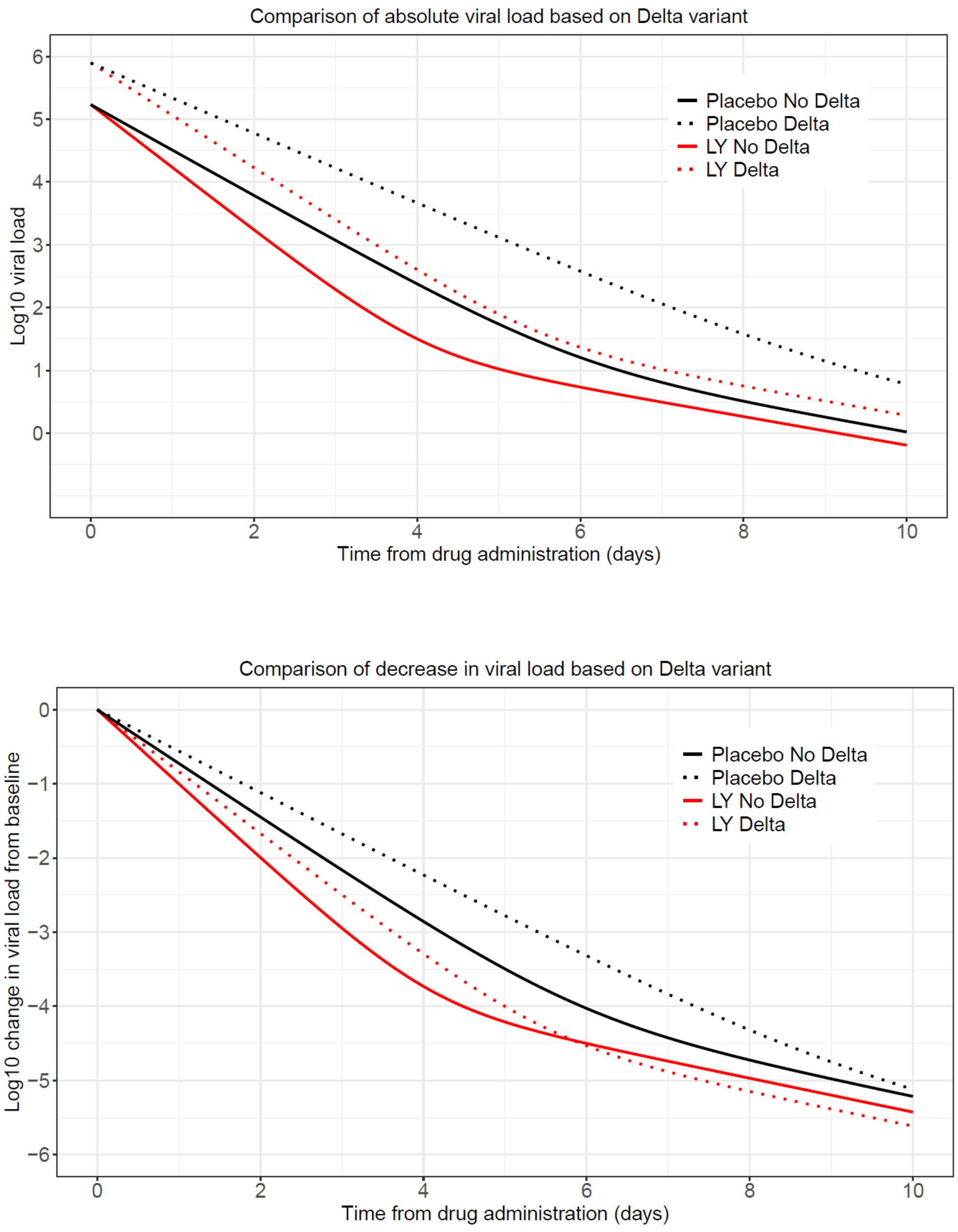

**Figure S2: Model-estimated typical viral load profiles showing impact of infection with the Delta variant.** Change in viral load from baseline. Abbreviation: LY = 175 mg bebtelovimab.

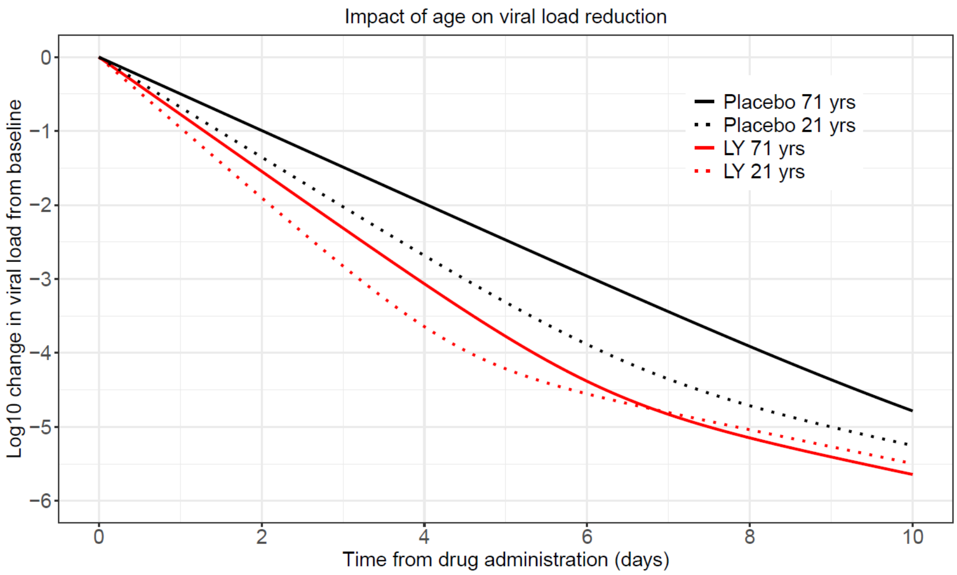

**Figure S3:** Model-estimated typical viral load profiles showing impact of age on viral dynamics.

**Table S5**: BLAZE-4 Phase 1: Overview of Adverse Events following IV infusion

| | **Pooled Placebo**<br>**N = 10** | **70 mg (BEB)**<br>**N = 6** | **175 mg (BEB)**<br>**N = 6** | **175 mg (BEB)**<br>**N = 6** | **700 mg (BAM) + 1400 mg (ETE) + 175 mg (BEB)**<br>**N = 6** | **1750 mg (BEB)**<br>**N = 6** | **Total**<br>**N = 40** |
|---|---|---|---|---|---|---|---|
| | | **Dose 1 140 mg/min** | **Dose 2a 140 mg/min** | **Dose 2b 350 mg/min** | **Dose 2c 350 mg/min** | **Dose 3 350 mg/min** | |
| Participants with $^{3}$1 TEAE n (%) | 1 (10.0) | 0 | 2 (33.3) | 0 | 1 (16.7) | 1 (16.7) | 5 (12.5) |
| TEAE by severity[a] n (%) | | | | | | | |
| Mild | 1 (10.0) | 0 | 2 (33.3) | 0 | 1 (16.7) | 1 (16.7) | 5 (12.5) |
| Moderate | 0 | 0 | 0 | 0 | 0 | 0 | 0 |
| Severe | 0 | 0 | 0 | 0 | 0 | 0 | 0 |
| Deaths n (%) | 0 | 0 | 0 | 0 | 0 | 0 | 0 |
| SAEs n (%) | 0 | 0 | 0 | 0 | 0 | 0 | 0 |
| Discontinuation from study treatment due to AEs n (%) | 0 | 0 | 0 | 0 | 0 | 0 | 0 |

Abbreviations: BAM = bamlanivimab; ETE = etesevimab; BEB = bebtelovimab; mg = miligrams; min = minutes; AE = adverse event.; N = number of subjects in the analysis population; n = number of subjects in the specified category; SAE = serious adverse event; TEAE = treatment-emergent adverse event.

[a]Patients with multiple occurrences of the same event are counted under the highest severity.

Note: Patients with multiple occurrences of these categories are counted once for each category. Patients may be counted in more than one category. Deaths are also included as serious adverse events and discontinuations due to adverse events.
