## Supplementary material for "Bebtelovimab, alone or together with bamlanivimab and etesevimab, as a broadly neutralizing monoclonal antibody treatment for mild to moderate, ambulatory COVID-19": Redacted PYAH Protocol

### Title Page

**Confidential Information**

The information contained in this document is confidential and is intended for the use of clinical investigators. It is the property of Eli Lilly and Company or its subsidiaries and should not be copied by or distributed to persons not involved in the clinical investigation of LY3819253, LY3832479, LY3853113, and CCI [redacted] unless such persons are bound by a confidentiality agreement with Eli Lilly and Company or its subsidiaries.

**Note to Regulatory Authorities:** This document may contain protected personal data and/or commercially confidential information exempt from public disclosure. Eli Lilly and Company requests consultation regarding release/redaction prior to any public release. In the United States, this document is subject to Freedom of Information Act (FOIA) Exemption 4 and may not be reproduced or otherwise disseminated without the written approval of Eli Lilly and Company or its subsidiaries.

**Protocol Title:**

A Randomized, Double-blind, Placebo-Controlled, Phase 2 Study to Evaluate the Efficacy and Safety of Mono and Combination Therapy with Monoclonal Antibodies in Participants with Mild to Moderate COVID-19 Illness (BLAZE-4)

**Protocol Number:** J2X-MC-PYAH

**Amendment Number:** g

**Compound(s):** LY3819253, LY3832479, LY3853113, CCI [redacted]

**Study Phase:** 2

**Short Title:** A randomized, double-blind, placebo-controlled, Phase 2 study to evaluate combinations of monoclonal antibodies in participants with mild to moderate COVID-19 illness

**Sponsor Name:** Eli Lilly and Company

**Legal Registered Address:** Indianapolis, Indiana, USA 46285

**Regulatory Agency Identifier Number(s)**

IND: 150440

**Approval Date:** Protocol Amendment (g) Electronically Signed and Approved by Lilly on date provided below.

Approval Date: 18-May-2021 GMT

**Medical Monitor Name and Contact Information will be provided separately**

### Protocol Amendment Summary of Changes Table

| DOCUMENT HISTORY | |
|---|---|
| **Document** | **Date** |
| Amendment f | 16 Mar 2021 |
| Amendment e | 25 Jan 2021 |
| Amendment d | 15 Jan 2021 |
| Amendment c | 06 Jan 2021 |
| Amendment b | 03 Dec 2020 |
| Amendment a | 16 Oct 2020 |
| Original Protocol | 5 Oct 2020 |

#### Amendment g

#### Overall Rationale for the Amendment:

The purpose of this amendment is to incorporate a new treatment arm into the study design. Additionally, minor clarifications and edits were made for operational efficiency and consistency.

| Section # and Name | Description of Change | Brief Rationale |
|---|---|---|
| 1.1 Synopsis; 1.2 Schema; 1.3 Schedule of Activities; 3.4 Treatment Arms 12-14; 4.1 Overall Design; 4.2 Scientific Rationale for Study Design; 4.3 Justification for Dose; 6.1 Study Intervention(s) Administered; 6.3 Measure to Minimize Bias: Randomization and Blinding; 9.2 Sample Size Determination; 9.4 Statistical Analyses; 9.4.2 Primary Endpoints | Added treatment arm 14 to study design | Allowance of additional high-risk participants in triplet combination cohort |
| 2.2 Background | Updated LY3853113 pseudovirus assay information | Alignment with Investigator's Brochure |
| 5.1 Inclusion Criteria | Specified that criteria 27 and 28 are applicable to treatment arms 12 and 13 only | Differentiation of high-risk participants applicable for treatment arms 12 and 13 and treatment arm 14 |
| 5.1 Inclusion Criteria | Added new inclusion criterion 30 for treatment arm 14 | Differentiation of high-risk participants applicable for treatment arms 12 and 13 and treatment arm 14 |
| 5.2 Exclusion Criteria | Specified in exclusion criterion 22 that participation in previous SARS-CoV-2 vaccine study is excluded if participant has received a SARS-CoV-2 vaccine or is currently blinded to treatment allotment | Clarification |

| Section # and Name | Description of Change | Brief Rationale |
|---|---|---|
| 6.1 Study Intervention(s) Administered; 8.2.2 Vital Signs | Removed instructions for multiple administrations in treatment arms 9-14 | Arms 9-14 will not utilize multiple administrations |
| Throughout | Minor editorial and formatting changes | Minor, therefore not detailed |

### 1. Protocol Summary

#### 1.1. Synopsis

**Protocol Title:**

A Randomized, Double-blind, Placebo-Controlled, Phase 2 Study to Evaluate the Efficacy and Safety of Mono and Combination Therapy with Monoclonal Antibodies in Participants with Mild to Moderate COVID-19 Illness (BLAZE-4)

**Rationale:**

The severe acute respiratory syndrome coronavirus 2 (SARS-CoV-2) has resulted in the pandemic spread of coronavirus disease 2019 (COVID-19), which in critical cases results in progressive pulmonary failure, complications with acute respiratory distress syndrome (ARDS), and death. There is an urgent need for effective therapeutics to modify disease outcomes.

LY3819253, LY3832479, and LY3853113 are neutralizing IgG1 monoclonal antibodies (mAb) to the spike protein of SARS-CoV-2. They are designed to block viral attachment and entry into human cells, and to neutralize the virus, potentially treating even established COVID-19 by providing a combination of specific, potent, neutralizing antibodies to SARS-CoV-2.

[redacted]

The aim of the study is to evaluate the impact of [redacted] LY3853113 alone, [redacted], LY3853113+ LY3819253 + LY3832479, and [redacted] on viral clearance and clinical outcomes in participants with COVID-19 illness. Additionally, the study aims to characterize the safety profile of LY3853113 alone and in combination with LY3819253 and LY3832479 in participants at high risk of developing severe COVID-19 illness. The data from this study will inform decisions for the clinical development of LY3819253, LY3832479, LY3853113, and [redacted]

**Objectives and Endpoints:**

[redacted]

CCI

[redacted]

CCI

[redacted]

CCI [redacted]

**Treatment Arms 9-11:**

| | |
|---|---|
| **Primary** | |
| Characterize the effect of LY3853113 alone and in combination with LY3819253 and LY3832479 after intravenous infusion compared to placebo on SARS-CoV-2 viral load | • Proportion of participants with SARS-CoV-2 viral load greater than 5.27 on Day 7 |
| **Secondary**<br>The secondary objectives are to characterize the effect of LY3853113 alone and in combination with LY3819253 and LY3832479 after intravenous infusion compared to placebo on… | |
| • overall participant clinical status | • Proportion (percentage) of participants who experience these events by Day 29<br> o COVID-19 related hospitalization (defined as ≥24 hours of acute care)<br> o Death |
| • SARS-CoV-2 viral load and viral clearance | • 75th Percentile of SARS-CoV-2 viral load at Day 7<br>• Change from baseline to<br> o Day 3<br> o Day 5<br> o Day 7<br> o Day 11<br>• Proportion of participants that achieve SARS-CoV-2 clearance (Days 3, 5, 7, 11, and 29)<br>• Time to SARS-CoV-2 clearance<br>• SARS-CoV-2 viral load area under the response-time curve (AUC) assessed through Day 11 |
| • symptom resolution | • Time to symptom resolution<br>• Time to sustained symptom resolution<br>• Proportion of participants demonstrating symptom resolution via the symptom questionnaire on Days 2-11, 22, and 29 |

| | |
|---|---|
| • symptom improvement | • Time to symptom improvement<br>• Proportion of participants demonstrating symptom improvement via the symptom questionnaire on Days 2-11, 22, and 29 |
| • safety | • Safety assessments such as AEs and SAEs |
| • overall participant clinical status | • Proportion (percentage) of participants who experience these events by Day 22<br>o COVID-19 related hospitalization (defined as ≥24 hours of acute care)<br>o Death |
| • Characterize clinical status for participants | • Proportion (percentage) of participants who experience these events through Day 29:<br>o COVID-19 related hospitalization (defined as ≥24 hours of acute care),<br>o COVID-19 related emergency room visit, or<br>o death |
| • Characterize the pharmacokinetics of LY3853113, LY3819253, and LY3832479 | • Mean concentration of LY3853113, LY3819253, and LY3832479 on Day 29 |

Abbreviations: AE = adverse event; SAE = serious adverse event; SARS-CoV-2 = severe acute respiratory syndrome coronavirus 2.

#### Treatment Arms 12-14:

| **Primary** | |
|---|---|
| Characterize the safety profile of LY3853113 alone and in combination with LY3819253 and LY3832479 after intravenous infusion on SARS-CoV-2 viral load | • Safety assessments such as AEs and SAEs |
| **Secondary**<br>The secondary objectives are to characterize the effect of LY3853113 alone and in combination with LY3819253 and LY3832479 after intravenous infusion on… | |
| • overall participant clinical status | • Proportion (percentage) of participants who experience these events by Day 29<br>o COVID-19 related hospitalization (defined as ≥24 hours of acute care)<br>o Death |
| • SARS-CoV-2 viral load and viral clearance | • 75th Percentile of SARS-CoV-2 viral load at Day 7<br>• Change from baseline to<br>o Day 3<br>o Day 5<br>o Day 7<br>o Day 11<br>• Proportion of participants that achieve SARS-CoV-2 clearance (Days 3, 5, 7, 11, and 29) |

| | |
|---|---|
| | • Time to SARS-CoV-2 clearance<br>• SARS-CoV-2 viral load area under the response-time curve (AUC) assessed through Day 11<br>• Proportion of participants with SARS-CoV-2 viral load greater than 5.27 on Day 7 |
| • symptom resolution | • Time to symptom resolution<br>• Time to sustained symptom resolution<br>• Proportion of participants demonstrating symptom resolution via the symptom questionnaire on Days 2-11, 22, and 29 |
| • symptom improvement | • Time to symptom improvement<br>• Proportion of participants demonstrating symptom improvement via the symptom questionnaire on Days 2-11, 22, and 29 |
| • overall participant clinical status | • Proportion (percentage) of participants who experience these events by Day 22<br>o COVID-19 related hospitalization (defined as ≥24 hours of acute care)<br>o Death |
| • Characterize clinical status for participants | • Proportion (percentage) of participants who experience these events through Day 29:<br>o COVID-19 related hospitalization (defined as ≥24 hours of acute care),<br>o COVID-19 related emergency room visit, or<br>o death |
| • Characterize the pharmacokinetics of LY3853113, LY3819253, and LY3832479 | • Mean concentration of LY3853113, LY3819253, and LY3832479 on Day 29 |

Abbreviations: AE = adverse event; SAE = serious adverse event; SARS-CoV-2 = severe acute respiratory syndrome coronavirus 2.

**Overall Design:**

This is a Phase 2, randomized, single-dose study in participants with mild to moderate COVID-19 illness. Treatment arms 1-11 are double-blind and placebo controlled, and treatment arms 12-14 are open label.

**Design Outline**

***Screening***

Interested participants will sign the appropriate informed consent and child/adolescent assent document(s), as appropriate, prior to completion of any procedures.

The investigator will review symptoms, risk factors and other non-invasive inclusion and exclusion criteria prior to any invasive procedures. If the participant is eligible after this review, then the site will perform the invasive procedures to confirm eligibility.

***Double-blind Treatment and Assessment Period***

This is the general sequence of events during the treatment and assessment period:

- Complete baseline procedures and sample collection

- Participants are randomized to an intervention group
- Participants receive study intervention, and
- Complete all safety monitoring and post-treatment sample collection.

This table describes the visit and activity types for this study.

| Study Day | Activity | Visit Type |
|---|---|---|
| 1 | Follow SoA | Site |
| 2, 4, 6, and 22 | Follow SoA | Telephone visit |
| 3, 5, 7, 11, 29, ED, 60, and 85 | Follow SoA | May be conducted as outpatient clinic or home visit. Symptom assessments can be collected via telephone visit. |
| 8, 9, 10 | Collection of symptom assessments | Telephone visit |
| Early discontinuation and follow-up | Follow SoA | May be conducted as outpatient clinic or home visit |
| CCI [redacted] | [redacted] | [redacted] |

Abbreviations: SoA = schedule of activities

If a participant is hospitalized, procedures and assessments will continue per the SoA (Section 1.3).

This table describes discharge from hospital scenarios if an outpatient is subsequently hospitalized.

***Discharge from hospital (Outpatients Subsequently Hospitalized)***

| If hospital discharge… | Then… |
|---|---|
| Occurs prior to Day 29 | participants will be asked to complete the remaining study assessments at the timepoints indicated in the SoA (Section 1.3).<br>NOTE: Strategies to manage infection risks and reduce the burden of return visits should be used by sites, such as home visits. |
| Occurs on the same day as a study assessment visit | assessments do not need to be repeated if the study assessments occurred within 8 hours of discharge and there has been no change in clinical status and the information is available to the site. |
| Does not occur at or prior to Day 29 | assessments will continue to occur weekly up to Day 60 or until hospital discharge, whichever is sooner. |

Abbreviations: SoA = schedule of activities

***Post-treatment follow-up***

Post-treatment follow-up assessments will be conducted at Day 60 and Day 85 to assess clinical status and for adverse events. Strategies to manage infection risks and reduce the burden of return visits may be used by sites, such as home visits. CCI [redacted]

**Disclosure Statement**: This is a treatment study that is participant and investigator blinded, unless noted otherwise.

**Number of Participants:**

[redacted]

*Treatment arms 9-11*

The planned sample size is approximately 122 participants per treatment arm.

*Treatment arms 12-14*

Approximately 100 participants will be enrolled into treatment arm 12 and approximately 50 participants will be enrolled into treatment arm 13. Approximately 140 participants will be enrolled into treatment arm 14 once treatment arms 12 and 13 have completed enrollment.

**Intervention Groups and Duration:**

This table describes the planned treatment arms.

[redacted]

| | | | |
|---|---|---|---|
| 9 | 175 mg | LY3853113 | Low risk |
| 10 | 175 mg + 700 mg + 1400 mg | LY3853113 + LY3819253 + LY3832479 | Low risk |
| 11 | --- | Placebo | Low risk |
| 12 | 175 mg | LY3853113 | High risk[a] |
| 13 | 175 mg + 700 mg + 1400 mg | LY3853113 + LY3819253 + LY3832479 | High risk[a] |
| 14 | 175 mg + 700 mg + 1400 mg | LY3853113 + LY3819253 + LY3832479 | High risk[b] |

a See Section 5.1, Inclusion Criteria 27 and 28 for definition of high-risk participants applicable for treatment arms 12 and 13

b See Section 5.1, Inclusion Criterion 30 for definition of high-risk participants applicable for treatment arm 14

[redacted]

Treatment arm 11 is the corresponding placebo control for treatment arms 9 and 10.

Optional treatment arms may be added based on interim analysis results and/or other emerging data.

Participants will receive a single treatment on Day 1, assessments occur to Day 29 and follow-up to Day 85 for [redacted] 9-14, [redacted].

**Data Monitoring Committee:** Yes, there will be an assessment committee.

#### 1.2. Schema

CCI

[redacted]

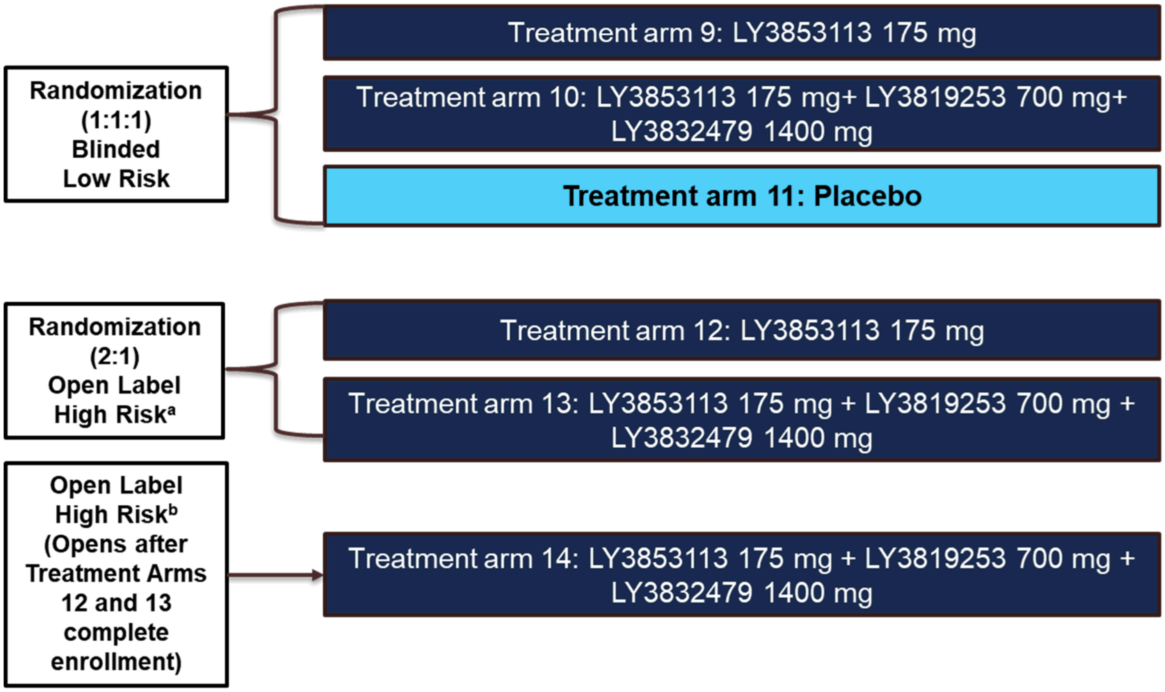

a See Section 5.1, Inclusion Criteria 27 and 28 for definition of high-risk participants applicable for treatment arms 12 and 13

b See Section 5.1, Inclusion Criterion 30 for definition of high-risk participants applicable for treatment arm 14

**Note:** Additional arms may be added based on interim analysis results and other emerging data.

Abbreviations: LY = Lilly study intervention.

**Figure 1. Study J2X-MC-PYAH schema**

#### 1.3. Schedule of Activities (SoA)

Assessments obtained previously as part of routine clinical care may be used as the baseline assessment if they were done no more than 48 hours before randomization. Visits may be conducted as a telephone call, outpatient clinic or home visit, as long as the protocol SoA is followed. Refer to the study day and visit type table in Section 4.1.1 for additional clarification.

CCI

[redacted]

CCI

[redacted]

CCI

[redacted]

CCI

[redacted]

CCI

[redacted]

CCI

[redacted]

CCI

[redacted]

CCI

[redacted]

Abbreviations: ACVPU = alert, consciousness, verbal, pain, unresponsive scale; ADA = anti-drug antibody; AEs = adverse events; BP = blood pressure; CRF = case report form; ED = early discontinuation visit; FiO2 = fraction of inspired oxygen in the air; ICU = intensive care unit; NP = nasopharyngeal; NSAIDs = non-steroidal anti-inflammatory drugs; SAEs = serious adverse events; SARS-CoV-2 = severe acute respiratory syndrome coronavirus 2; SpO2 = saturation of peripheral oxygen; WOCBP = women of child-bearing potential.

CCI

[redacted]

CCI

[redacted]

CCI

[redacted]

CCI

[redacted]

CCI

[redacted]

CCI

[redacted]

CCI

[redacted]

CCI

[redacted]

CCI

[redacted]

CCI

[redacted]

Abbreviations: ACVPU = alert, consciousness, verbal, pain, unresponsive scale; ADA = anti-drug antibody; AEs = adverse events; BP = blood pressure; CRF = case report form; ED = early discontinuation visit; FiO2 = fraction of inspired oxygen in the air; ICU = intensive care unit; NP = nasopharyngeal; NSAIDs = non-steroidal anti-inflammatory drugs; SAEs = serious adverse events; SARS-CoV-2 = severe acute respiratory syndrome coronavirus 2; SpO2 = saturation of peripheral oxygen; WOCBP = women of child-bearing potential.

##### 1.3.3. Treatment Arms 9-14

| Schedule of Activities for Treatment Arms 9-14 | | | | | | | | | | | | | | | | |
|---|---|---|---|---|---|---|---|---|---|---|---|---|---|---|---|---|
| | **Screen** | **Double-blind treatment and assessments** | | | | | | | | | | ED | Follow-up if hospital inpatient on Day 29 | **Post-treatment follow-up** | | **Comments** |
| **Study Day** | | **1** | **2*** | **3** | **4*** | **5** | **6*** | **7** | **11** | **22*** | **29** | ED | Every 7 days until discharge or Day 60 | **60** | **85** | Screening and Day 1 procedures may occur on the same day.<br>* = telephone visit(s) for all treatment arms |
| **Visit window (number of days)** | | -- | -- | +1 | -- | +1 | -- | +1 | -2/+3 | ±3 | ±3 | ±2 | ±2 | ±4 | ±4 | Visits may not be combined. |
| **Procedures** | | | | | | | | | | | | | | | | |
| Informed Consent | X | | | | | | | | | | | | | | | |
| Informed assent for adolescent participants | X | | | | | | | | | | | | | | | Parent or legal guardian signs informed consent form and participant signs assent form, as appropriate per local requirements. |
| Inclusion and exclusion criteria review | X | | | | | | | | | | | | | | | |
| Demographics | X | | | | | | | | | | | | | | | Including age, gender, race, ethnicity |
| Preexisting conditions and medical history | X | | | | | | | | | | | | | | | Obtained from interview or available information. Includes: vaccination history (including SARS-CoV-2 vaccinations), risk factors and comorbidities associated with severe COVID-19 illness, such as living in a nursing home or long-term care facility. |
| Prespecified medical history: COVID-19 | X | | | | | | | | | | | | | | | Includes COVID-19 diagnosis date and onset of COVID-19 symptoms. |
| Height | | X | | | | | | | | | | | | | | |
| Weight | | X | | | | | | | | | | | | | | |

| Schedule of Activities for Treatment Arms 9-14 | | | | | | | | | | | | | | | | |
|---|---|---|---|---|---|---|---|---|---|---|---|---|---|---|---|---|
| | **Screen** | **Double-blind treatment and assessments** | | | | | | | | | | ED | Follow-up if hospital inpatient on Day 29 | **Post-treatment follow-up** | | **Comments** |
| **Study Day** | | **1** | **2*** | **3** | **4*** | **5** | **6*** | **7** | **11** | **22*** | **29** | ED | Every 7 days until discharge or Day 60 | **60** | **85** | Screening and Day 1 procedures may occur on the same day.<br>* = telephone visit(s) for all treatment arms |
| **Visit window (number of days)** | | -- | -- | +1 | -- | +1 | -- | +1 | -2/+3 | ±3 | ±3 | ±2 | ±2 | ±4 | ±4 | Visits may not be combined. |
| Prior treatments of special interest within the last 30 days | X | | | | | | | | | | | | | | | NSAIDs, antivirals, antibiotics, anti-malarials, corticosteroids, immunomodulators or other investigational treatments. |
| Substance use (Tobacco) | X | | | | | | | | | | | | | | | Includes use of e-cigarettes, such as vaping |
| Concomitant medications | X | X | X | X | X | X | X | X | X | X | X | X | X | X | X | |
| Adverse events (AEs) | X | X | X | X | X | X | X | X | X | X | X | X | X | X | X | Any events that occur after signing the informed consent are considered AEs as defined in Section 10.3.<br>Additional details regarding reporting frequency and method of detecting AEs and SAEs can be found in Section 8.3. |
| **Physical Evaluation or Clinical Assessments** | | | | | | | | | | | | | | | | |
| Physical examination | X | | | | | | | | | | | | | | | |
| Symptom-directed physical exam | | | | X | | | | | | | X | X | | | | As indicated based on participant status and standard of care. May be performed by qualified personnel per local regulations. |

| Schedule of Activities for Treatment Arms 9-14 | | | | | | | | | | | | | | | | |
|---|---|---|---|---|---|---|---|---|---|---|---|---|---|---|---|---|
| | **Screen** | **Double-blind treatment and assessments** | | | | | | | | | | ED | Follow-up if hospital inpatient on Day 29 | **Post-treatment follow-up** | | **Comments** |
| **Study Day** | | **1** | **2*** | **3** | **4*** | **5** | **6*** | **7** | **11** | **22*** | **29** | ED | Every 7 days until discharge or Day 60 | **60** | **85** | Screening and Day 1 procedures may occur on the same day.<br>* = telephone visit(s) for all treatment arms |
| **Visit window (number of days)** | | -- | -- | +1 | -- | +1 | -- | +1 | -2/+3 | ±3 | ±3 | ±2 | ±2 | ±4 | ±4 | Visits may not be combined. |
| Vital signs and Oxygen Support | X | X | | X | | X | | X | X | | X | X | X | X | X | Documentation of hospital-based exam is acceptable.<br>Includes: body temperature, pulse rate, BP, respiratory rate, SpO2, and supplemental oxygen flow rate, FiO2 if known, method of delivery, if applicable, and oxygen support procedures.<br>Record SpO2 while participant is at rest.<br>**Screening visit only**: SpO2 while breathing room air. Data not collected on CRF.<br>**Day 1 timing:**<br>• immediately before administration<br>• immediately following completion of infusion if infusion is <15 minutes<br>• every 15 minutes during the infusion, as possible and applicable<br>• every 30 minutes for 1 hour after the end of infusion<br>See Section 8.2.2 for data collected on CRF.<br>**All other study days:** once daily. |

**Schedule of Activities for Treatment Arms 9-14**

| | Screen | Double-blind treatment and assessments | | | | | | | | | | ED | Follow-up if hospital inpatient on Day 29 | Post-treatment follow-up | | Comments |
|---|---|---|---|---|---|---|---|---|---|---|---|---|---|---|---|---|
| **Study Day** | | **1** | **2*** | **3** | **4*** | **5** | **6*** | **7** | **11** | **22*** | **29** | ED | Every 7 days until discharge or Day 60 | **60** | **85** | Screening and Day 1 procedures may occur on the same day.<br>* = telephone visit(s) for all treatment arms |
| **Visit window (number of days)** | | -- | -- | +1 | -- | +1 | -- | +1 | -2/+3 | ±3 | ±3 | ±2 | ±2 | ±4 | ±4 | Visits may not be combined. |
| Hospitalization events | | | Daily | | | | | | | X | X | X | X | X | X | Record if the following events occur or occurred since prior visit:<br>• Emergency room visits<br>• hospitalized<br>• ICU admittance,<br>• Extended care facility admittance, and<br>• Discharge |
| **Laboratory Tests and Sample Collection** | | | | | | | | | | | | | | | | |
| Hematology | | X | | X | | | | | X | | X | X | | | | Day 1: before treatment administration<br>All other days: sample may occur at any time during the day<br>No samples needed if participant is hospitalized<br>Lilly-designated central laboratory |
| Clinical Chemistry | | X | | X | | | | | X | | X | X | | | | Day 1: before treatment administration<br>All other days: sample may occur at any time during the day<br>No samples needed if participant is hospitalized<br>Lilly-designated central laboratory |
| C-reactive protein (CRP); high - sensitivity | | X | | X | | | | | X | | X | X | | | | For adults only<br>Day 1: before treatment administration<br>All other days: sample may occur at any time during the day<br>No sample needed if participant is hospitalized<br>Lilly-designated central laboratory |

| Schedule of Activities for Treatment Arms 9-14 | | | | | | | | | | | | | | | | |
|---|---|---|---|---|---|---|---|---|---|---|---|---|---|---|---|---|
| | **Screen** | **Double-blind treatment and assessments** | | | | | | | | | | ED | Follow-up if hospital inpatient on Day 29 | **Post-treatment follow-up** | | **Comments** |
| **Study Day** | | **1** | **2*** | **3** | **4*** | **5** | **6*** | **7** | **11** | **22*** | **29** | ED | Every 7 days until discharge or Day 60 | **60** | **85** | Screening and Day 1 procedures may occur on the same day.<br>* = telephone visit(s) for all treatment arms |
| **Visit window (number of days)** | | -- | -- | +1 | -- | +1 | -- | +1 | -2/+3 | ±3 | ±3 | ±2 | ±2 | ±4 | ±4 | Visits may not be combined. |
| Ferritin | | X | | X | | | | | X | | X | X | | | | For adults only<br>Day 1: before treatment administration<br>All other days: sample may occur at any time during the day<br>No sample needed if participant is hospitalized<br>Lilly-designated central laboratory |
| D-dimer | | X | | X | | | | | X | | X | X | | | | For adults only<br>Day 1: before treatment administration<br>All other days: sample may occur at any time during the day<br>No sample needed if participant is hospitalized<br>Lilly-designated central laboratory |
| Procalcitonin | | X | | X | | | | | X | | X | X | | | | For adults only<br>Day 1: before treatment administration<br>All other days: sample may occur at any time during the day<br>No sample needed if participant is hospitalized<br>Lilly-designated central laboratory |
| Troponin I and Troponin T | | X | | X | | | | | X | | X | X | | | | For adults only<br>Day 1: before treatment administration<br>All other days: sample may occur at any time during the day<br>No sample needed if participant is hospitalized<br>Lilly-designated central laboratory |

| Schedule of Activities for Treatment Arms 9-14 | | | | | | | | | | | | | | | | |
|---|---|---|---|---|---|---|---|---|---|---|---|---|---|---|---|---|
| | **Screen** | **Double-blind treatment and assessments** | | | | | | | | | | ED | Follow-up if hospital inpatient on Day 29 | **Post-treatment follow-up** | | **Comments** |
| **Study Day** | | **1** | **2*** | **3** | **4*** | **5** | **6*** | **7** | **11** | **22*** | **29** | ED | Every 7 days until discharge or Day 60 | **60** | **85** | Screening and Day 1 procedures may occur on the same day.<br>* = telephone visit(s) for all treatment arms |
| **Visit window (number of days)** | | -- | -- | +1 | -- | +1 | -- | +1 | -2/+3 | ±3 | ±3 | ±2 | ±2 | ±4 | ±4 | Visits may not be combined. |
| Documentation of positive SARS-CoV-2 viral infection | X | | | | | | | | | | | | | | | Sample for first positive test must be collected within 3 days prior to start of infusion.<br>Local laboratory and/or Point-of-Care testing. |
| Urine or serum pregnancy | X | | | | | | | | | | | | | X | X | Only for WOCBP (Section 10.4 Appendix 4)<br>Local laboratory |
| Pharmacokinetic (PK) sample for LY3853113 LY3819253 LY3832479 | | X | | X | | | | X | X | | X | X | | X | X | Day 1: no pre-dose sample needed; one post-dose sample within 30 minutes after the end of IV infusion (may include the flush).<br>All other scheduled samples may occur at any time during the day.<br>No samples needed if participant is hospitalized.<br>Lilly-designated central laboratory |
| Immunogenicity (ADA) sample for LY3853113 LY3819253 LY3832479 | | X | | | | | | | X | | X | X | | X | X | Day 1: collect before treatment administration.<br>All other scheduled and unscheduled immunogenicity samples should be collected with a time-matched PK sample.<br>No samples needed if participant is hospitalized<br>Lilly-designated central laboratory |
| Pharmacodynamic (PD) NP swab | | X | | X | | X | | X | X | | X | X | | | | Swab is taken from both nostrils.<br>Day 1: swab before treatment administration.<br>No samples needed if participant is hospitalized<br>Lilly-designated central laboratory |

| Schedule of Activities for Treatment Arms 9-14 | | | | | | | | | | | | | | | | |
|---|---|---|---|---|---|---|---|---|---|---|---|---|---|---|---|---|
| | **Screen** | **Double-blind treatment and assessments** | | | | | | | | | | ED | Follow-up if hospital inpatient on Day 29 | **Post-treatment follow-up** | | **Comments** |
| **Study Day** | | **1** | **2*** | **3** | **4*** | **5** | **6*** | **7** | **11** | **22*** | **29** | ED | Every 7 days until discharge or Day 60 | **60** | **85** | Screening and Day 1 procedures may occur on the same day.<br>* = telephone visit(s) for all treatment arms |
| **Visit window (number of days)** | | -- | -- | +1 | -- | +1 | -- | +1 | -2/+3 | ±3 | ±3 | ±2 | ±2 | ±4 | ±4 | Visits may not be combined. |
| Exploratory biomarker samples | | X | | X | | X | | | X | | X | X | | | | Day 1: before treatment administration. |
| Exploratory serum sample | | X | | | | | | | | | | | | X | X | |
| Pharmacogenetics sample | | X | | | | | | | | | | | | | | For adults only<br>Lilly-designated central laboratory |
| **Randomization and Dosing** | | | | | | | | | | | | | | | | |
| Randomization | | X | | | | | | | | | | | | | | |
| Administer study intervention | | X | | | | | | | | | | | | | | Study intervention must be administered within 3 days from the time of first positive SARS-CoV-2 test sample collection.<br>For participants that receive dialysis on the same day as administration of study intervention, complete dialysis first followed by the study treatment.<br>Participants will be monitored for at least 1 hour after completion of treatment administration. |
| **Participant Questionnaire** | | | | | | | | | | | | | | | | |
| Symptoms (Patient Symptom Assessment) and overall clinical status | | Daily on Days 1-11 for outpatients only | | | | | | | | X | X | X | | X | X | Day 1: assess prior to dosing |

Abbreviations: ADA = anti-drug antibody; AEs = adverse events; BP = blood pressure; CRF = case report form; ED = early discontinuation visit; FiO2 = fraction of inspired oxygen in the air; ICU = intensive care unit; NP = nasopharyngeal; NSAIDS = non-steroidal anti-inflammatory drugs; PK = pharmacokinetics; SAEs = serious adverse events; SARS-CoV-2 = severe acute respiratory syndrome coronavirus 2; SpO2 = saturation of peripheral oxygen; WOCBP = women of child-bearing potential.

### 2. Introduction

The efficient community spread of severe acute respiratory syndrome coronavirus 2 (SARS-CoV-2) has resulted in the current pandemic of coronavirus disease 2019 (COVID-19), which in severe and critical cases results in progressive pulmonary infection, complicated by respiratory failure, with a high prevalence of acute respiratory distress syndrome. Although several therapies have been explored in severe COVID-19, none have improved survival, including antivirals, glucocorticoids, and immunoglobulins (Liu et al 2020).

LY3819253, LY3832479, and LY3853113 are neutralizing IgG1 monoclonal antibodies (mAb) to the spike protein of SARS-CoV-2. They are designed to block viral attachment and entry into human cells, and to neutralize the virus, potentially treating even established COVID-19 by providing a combination of specific, potent, neutralizing antibodies to SARS-CoV-2. The blocking of viral entry into respiratory cells and viral replication, and viral neutralization is expected to mitigate the severity of disease in patients in whom ongoing viral spread and replication is the primary driver of COVID-19 pathophysiology. The decrease in viral replication may also shorten a patient's extent and duration of viral shedding and transmission, positively impacting public health.

[redacted]

#### 2.1. Study Rationale

This study aims to evaluate the impact of [redacted] [redacted] LY3853113 alone, LY3853113 + LY3819253 + LY3832479, [redacted] on viral clearance and clinical outcomes in participants with mild to moderate COVID-19 illness. Additionally, this study aims to characterize the safety profile of LY3853113 alone and in combination with LY3819253 and LY3832479 in participants at high risk of developing severe COVID-19 illness. The data from this study will inform decisions for the clinical development of these neutralizing IgG1 mAbs.

#### 2.2. Background

**LY3819253 and LY3832479**

Nonclinical information for LY3819253 and LY3832479 is described in each respective Investigator's Brochure (IB).

Lilly is evaluating the safety, tolerability, pharmacokinetics (PK), and pharmacodynamics (PD) of LY3819253 in participants hospitalized for COVID-19, in a randomized, placebo-controlled, double-blind, single ascending dose, Phase 1, first in human study (Study J2W-MC-PYAA [PYAA]). This study has completed dosing.

Lilly is also evaluating the safety, tolerability, PK, and immunogenicity of LY3832479 in healthy participants, in a randomized, placebo-controlled, single dose, Phase 1 study (Study J2Z-MC-PGAA [PGAA]) under IND 150707. Concurrent with Study PGAA, LY3832479 is also under development in China in an ongoing Phase 1 clinical study in healthy participants, Study JS016-001-I.

Lilly is evaluating the safety, tolerability, PK, immunogenicity and efficacy of LY3819253 alone, or in combination with LY3832479, in a randomized, placebo-controlled, double-blind Phase 2 study in participants with mild to moderate COVID-19 (Study J2W-MC-PYAB [PYAB]). Dose levels administered in Study PYAB were informed by PYAA and PGAA. Specific treatment arms in PYAB are evaluating the safety and efficacy of LY3819253 in combination with LY3832479 in participants with risk factors for severe COVID-19. As of 04 November 2020, 542 participants received blinded treatment with either placebo or 2800 mg LY3819253 + 2800 mg LY3832479 in combination. As of that date, there have been no discontinuations due to AEs. Serious adverse events were reported for 4 participants across monotherapy and combination arms.

The SAEs were as follows:

- a severe SAE of upper abdominal pain in a participant who received placebo
- a mild SAE of urinary tract infection in a participant who received LY3819253 and LY3832479 combination therapy
- acute myocardial infarction in a participant who received blinded study treatment with either placebo or LY3819253 2800 mg in combination with LY3832479 2800 mg, and
- a severe SAE of diabetic ketoacidosis in a participant who received LY3819253 700 mg monotherapy

No infusion-related reactions were reported. One event of facial swelling and 1 event of hypersensitivity was noted on the day of infusion, with pruritis as the most common immediate hypersensitivity event. Other non-immediate hypersensitivity events reported included rash, stomatitis, and urticaria. A total of 20 patients receiving LY3819253 in combination with LY3832479 developed treatment-emergent adverse events (TEAEs). Most TEAES were mild to moderate in severity, similar across all dose groups, and comparable to placebo. The most frequently reported TEAEs were nausea and pruritus.

As of 24 September 2020, one serious infusion-related reaction was reported in a blinded ongoing study (J2W-NS-I001; ACTIV-2/A5401) of LY3819253 administered at 7000 mg versus placebo.

In addition, there is an ongoing Lilly and NIAID collaborative Phase 3 randomized, double-blind, placebo-controlled trial (Study J2X-MC-PYAD [PYAD]) evaluating the efficacy and safety of LY3819253 in preventing SARS-CoV-2 infection and COVID-19 in skilled nursing and assisted living facility residents and staff.

**LY3853113**

LY3853113 is a novel, highly potent IgG1 neutralizing mAb targeting the spike protein of SARS-CoV-2 that was created in partnership with AbCellera Biologics Inc. (AbCellera; Vancouver, Canada). It binds an epitope within the receptor binding domain (RBD) that is distinct from those bound by LY3819253 and LY3832479. LY3853113 can neutralize the Wuhan reference strain as well as individual residues present in recent variants of concern (i.e. L452R, D614G, N501Y, N439K, K417N, and E484K). Critically, pseudovirus assays demonstrate that LY3853113 can neutralize variants with the specific combination of RBD residues of the B.1.1.351 (South African origin), B.1.1.28 (P.1/Brazil origin**),** B.1.427/B.1.429 (California origin), and B.1.526 (New York origin) strains. These variants were recently reported in the United States (CDC 2021), thus it is imperative that new treatments for these emerging variants are developed and deployed quickly.

Refer to the LY3853113 IB for additional information.

CCI

[redacted]

#### 2.3. Benefit/Risk Assessment

**LY3819253, LY3832479, and LY3853113**

Information on the safety and tolerability of LY3819253 in humans will come from Studies PYAA and PYAB. Information on the safety and tolerability of LY3832479 will come from Studies PGAA and PYAB. Information on the safety and tolerability of LY3853113 will come from Study PYAH and PYAH Addendum (4).

Anticipated risk is considered low and is based on the known mechanism of action for human derived neutralizing antibodies in acute viral disease states. LY3819253, LY3832479, and LY3853113 consist of highly specific mAbs directed at foreign (non-human) epitope(s). The complementarity determining regions (CDRs) of the mAbs were derived from B lymphocytes of 2 individually convalescent naturally SARS-CoV-2-infected patients and, thus, have undergone natural positive and negative selection pressures in vivo, unlike humanized antibodies generated in mice. No clinically relevant off-target binding has been observed in tissue cross reactivity studies of membrane targets in human tissues. Therefore, off-target binding and tissue cross-reactivity are considered unlikely.

A theoretical risk is that these interventions may cause antibody-dependent enhancement (ADE) of viral replication. This is based on responses observed to some monoclonal antibody therapies used in other unrelated viral diseases. To address this risk, LY3819253 and LY3832479 have been assessed with in vitro cell culture models and, for LY3819253, an in vivo nonhuman primate model.

The risk of clinical ADE is considered low due to

- the structural features of LY3832479, which is engineered to suppress its binding to Fc receptors and C1qm
- the absence of ADE from in vitro studies, and
- the absence of ADE from in vivo nonhuman primate studies for LY3819253.

To date, there is no evidence of productive enhancement of ADE with SARS-CoV-2.

Additional manageable risks associated with most therapeutic monoclonal antibodies are the potential for infusion-related hypersensitivity and cytokine release reactions. Infusion-related hypersensitivity reactions have been observed in clinical studies and under the EUA.

The infusions in this study will be administered at a controlled rate, the study participants will be monitored closely, and adjustments in the infusion rate will be made and/or the infusion stopped, if indicated. Additional information regarding infusion reaction management options is in Section 6.1.1.

Clinical worsening of COVID-19 after administration of LY3819253 has been reported from post-authorization treatment within 24 hours of infusion. Signs and symptoms may include fever, hypoxia or increased respiratory difficulty, arrythmia (e.g., atrial fibrillation, sinus tachycardia, and bradycardia), fatigue, and altered mental status. Some of these events required hospitalization. It is not known if these events were related to LY3819253 use or were due to progression of COVID-19. Refer to the IB, and EUA Fact Sheet for additional information.

Given the data on LY3819253, LY3832479, and LY3853113, the well described safety profile of other therapeutic mAbs, and the lack of disease directed therapeutic options for patients with COVID-19 illness, the overall benefit-risk assessment for this study is considered favorable.

[redacted]

**Investigator's Brochures**

More detailed information about the known and expected benefits and risks and reasonably expected adverse events of LY3819253, LY3832479, LY3853113, and [redacted] may be found in each respective IB.

### 3. Objectives and Endpoints

CCI

[redacted]

[redacted]

syndrome coronavirus 2.

[redacted]

CCI

[redacted]

Abbreviations: AE = adverse event; SAE = serious adverse event; SARS-CoV-2 = severe acute respiratory syndrome coronavirus 2.

#### 3.3. Treatment Arms 9-11

| **Primary** | |
|---|---|
| Characterize the effect of LY3853113 alone and in combination with LY3819253 and LY3832479 after intravenous infusion compared to placebo on SARS-CoV-2 viral load | • Proportion of participants with SARS-CoV-2 viral load greater than 5.27 on Day 7 |
| **Secondary**<br>The secondary objectives are to characterize the effect of LY3853113 alone and in combination with LY3819253 and LY3832479 after intravenous infusion compared to placebo on… | |
| • overall participant clinical status | • Proportion (percentage) of participants who experience these events by Day 29<br>o COVID-19 related hospitalization (defined as ≥24 hours of acute care)<br>o Death |
| • SARS-CoV-2 viral load and viral clearance | • 75th Percentile of SARS-CoV-2 viral load at Day 7<br>• Change from baseline to<br>o Day 3<br>o Day 5<br>o Day 7<br>o Day 11<br>• Proportion of participants that achieve SARS-CoV-2 clearance (Days 3, 5, 7, 11, and 29)<br>• Time to SARS-CoV-2 clearance<br>• SARS-CoV-2 viral load area under the response-time curve (AUC) assessed through Day 11 |
| • symptom resolution | • Time to symptom resolution<br>• Time to sustained symptom resolution<br>• Proportion of participants demonstrating symptom resolution via the symptom questionnaire on Days 2-11, 22, and 29 |
| • symptom improvement | • Time to symptom improvement<br>• Proportion of participants demonstrating symptom improvement via the symptom questionnaire on Days 2-11, 22, and 29 |
| • safety | • Safety assessments such as AEs and SAEs |
| • overall participant clinical status | • Proportion (percentage) of participants who experience these events by Day 22<br>o COVID-19 related hospitalization (defined as ≥24 hours of acute care)<br>o Death |

| | |
|---|---|
| • Characterize clinical status for participants | • Proportion (percentage) of participants who experience these events through Day 29:<br>○ COVID-19 related hospitalization (defined as ≥24 hours of acute care),<br>○ COVID-19 related emergency room visit, or<br>○ death |
| • Characterize the pharmacokinetics of LY3853113, LY3819253, and LY3832479 | • Mean concentration of LY3853113, LY3819253, and LY3832479 on Day 29 |
| **Exploratory** | |
| • Characterize emergence of viral resistance to LY3853113 | • Comparison from baseline to the last evaluable time point up to Day 29 |

Abbreviations: AE = adverse event; SAE = serious adverse event; SARS-CoV-2 = severe acute respiratory syndrome coronavirus 2.

#### 3.4. Treatment Arms 12-14

| **Primary** | |
|---|---|
| Characterize the safety profile of LY3853113 alone and in combination with LY3819253 and LY3832479 after intravenous infusion on SARS-CoV-2 viral load | • Safety assessments such as AEs and SAEs |
| **Secondary**<br>The secondary objectives are to characterize the effect of LY3853113 alone and in combination with LY3819253 and LY3832479 after intravenous infusion on… | |
| • overall participant clinical status | • Proportion (percentage) of participants who experience these events by Day 29<br>○ COVID-19 related hospitalization (defined as ≥24 hours of acute care)<br>○ Death |
| • SARS-CoV-2 viral load and viral clearance | • 75th Percentile of SARS-CoV-2 viral load at Day 7<br>• Change from baseline to<br>○ Day 3<br>○ Day 5<br>○ Day 7<br>○ Day 11<br>• Proportion of participants that achieve SARS-CoV-2 clearance (Days 3, 5, 7, 11, and 29)<br>• Time to SARS-CoV-2 clearance<br>• SARS-CoV-2 viral load area under the response-time curve (AUC) assessed through Day 11<br>• Proportion of participants with SARS-CoV-2 viral load greater than 5.27 on Day 7 |

| | |
|---|---|
| • symptom resolution | • Time to symptom resolution<br>• Time to sustained symptom resolution<br>• Proportion of participants demonstrating symptom resolution via the symptom questionnaire on Days 2-11, 22, and 29 |
| • symptom improvement | • Time to symptom improvement<br>• Proportion of participants demonstrating symptom improvement via the symptom questionnaire on Days 2-11, 22, and 29 |
| • overall participant clinical status | • Proportion (percentage) of participants who experience these events by Day 22<br>o COVID-19 related hospitalization (defined as ≥24 hours of acute care)<br>o Death |
| • Characterize clinical status for participants | • Proportion (percentage) of participants who experience these events through Day 29:<br>o COVID-19 related hospitalization (defined as ≥24 hours of acute care),<br>o COVID-19 related emergency room visit, or<br>o death |
| • Characterize the pharmacokinetics of LY3853113, LY3819253, and LY3832479 | • Mean concentration of LY3853113, LY3819253, and LY3832479 on Day 29 |
| **Exploratory** | |
| • Characterize emergence of viral resistance to LY3853113 | • Comparison from baseline to the last evaluable time point up to Day 29 |

Abbreviations: AE = adverse event; SAE = serious adverse event; SARS-CoV-2 = severe acute respiratory syndrome coronavirus 2.

### 4. Study Design

#### 4.1. Overall Design

This is a Phase 2, randomized, single-dose study in participants with mild to moderate COVID-19 illness. Treatment arms 1-11 are double-blind and placebo controlled, and treatment arms 12-14 are open label.

##### 4.1.1. Design Outline

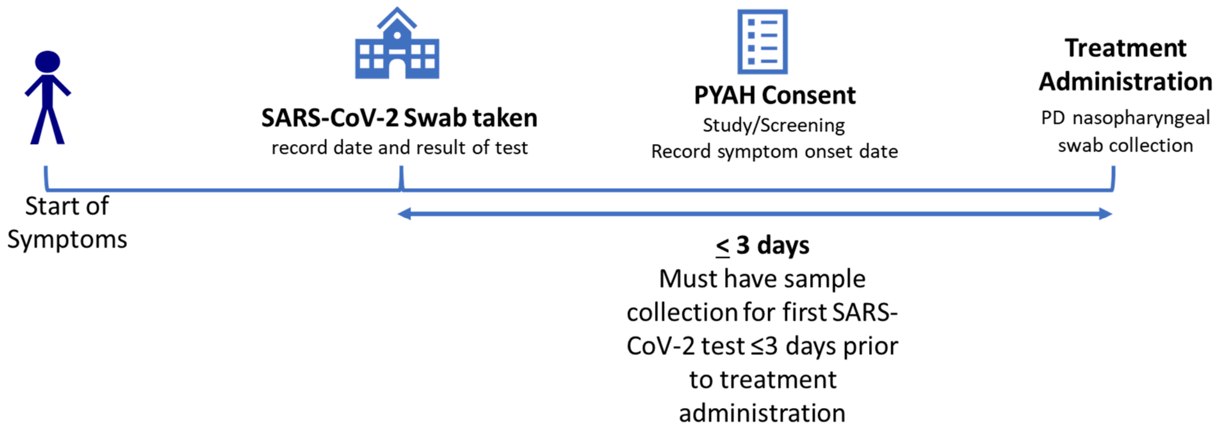

Abbreviations: PD = pharmacodynamic; PYAH = Study J2X-MC-PYAH.

**Figure 2. Overview of participant flow from time of SARS-CoV-2 symptoms to treatment administration.**

**Screening**

Interested participants will sign the appropriate informed consent and child/adolescent assent document(s), as appropriate, prior to completion of any procedures. The participant may enter the study with a previous positive SARS-CoV-2 viral test result from an external testing facility. This test result must be the first time the participant has tested positive for SARS-CoV-2.

The investigator will review symptoms, risk factors and other non-invasive inclusion and exclusion criteria prior to any invasive procedures. If the participant is eligible after this review, then the site will perform the invasive procedures to confirm eligibility.

**Double-blind Treatment and Assessment Period**

This is the general sequence of events during the treatment and assessment period:

- Complete baseline procedures and sample collection
- Participants are randomized to an intervention group
- Participants receive study intervention, and
- Complete all safety monitoring and post-treatment sample collection.

This table describes the planned treatment arms.

| | | | |
|---|---|---|---|
| CCI [redacted] | | | |
| 9 | 175 mg | LY3853113 | Low risk |
| 10 | 175 mg + 700 mg + 1400 mg | LY3853113 + LY3819253 + LY3832479 | Low risk |
| 11 | --- | Placebo | Low risk |
| 12 | 175 mg | LY3853113 | High risk[a] |
| 13 | 175 mg + 700 mg + 1400 mg | LY3853113 + LY3819253 + LY3832479 | High risk[a] |
| 14 | 175 mg + 700 mg + 1400 mg | LY3853113 + LY3819253 + LY3832479 | High risk[b] |

a See Section 5.1, Inclusion Criteria 27 and 28 for definition of high-risk participants applicable for treatment arms 12 and 13

b See Section 5.1, Inclusion Criterion 30 for definition of high-risk participants applicable for treatment arm 14

CCI [redacted]

Treatment arm 11 is the corresponding placebo control for treatment arms 9 and 10.

This table describes the visit and activity types for this study.

| **Study Day** | **Activity** | **Visit Type** |
|---|---|---|
| 1 | Follow SoA | Site |
| 2, 4, 6, and 22 | Follow SoA | Telephone visit |
| 3, 5, 7, 11, 29, ED, 60, and 85 | Follow SoA | May be conducted as outpatient clinic or home visit. Symptom assessments can be collected via telephone visit. |
| 8, 9, 10 | Collection of symptom assessments | Telephone visit |
| Early discontinuation and follow-up | Follow SoA | May be conducted as outpatient clinic or home visit |
| CCI [redacted] | [redacted] | [redacted] |

Abbreviations: SoA = schedule of activities

If a participant is hospitalized, procedures and assessments will continue per the SoA (Section 1.3).

This table describes discharge from hospital scenarios if an outpatient is subsequently hospitalized.

| If hospital discharge… | Then… |
|---|---|
| Occurs prior to Day 29 | participants will be asked to complete the remaining study assessments at the timepoints indicated in the SoA (Section 1.3).<br>NOTE: Strategies to manage infection risks and reduce the burden of return visits should be used by sites, such as home visits. |
| Occurs on the same day as a study assessment visit | assessments do not need to be repeated if the study assessments occurred within 8 hours of discharge and there has been no change in clinical status and the information is available to the site. |
| Does not occur at or prior to Day 29 | assessments will continue to occur weekly up to Day 60 or until hospital discharge, whichever is sooner. |

**Post-treatment follow-up**

Post-treatment follow-up assessments will be conducted at Day 60 and Day 85 to assess clinical status and for adverse events. Strategies to manage infection risks and reduce the burden of return visits may be used by sites, such as home visits. CCI [redacted]

#### 4.2. Scientific Rationale for Study Design

**Overall Design**

This study is designed to evaluate the efficacy of CCI [redacted], LY3853113 alone, CCI [redacted] LY3853113 in combination with LY3819253 and LY3832479, CCI [redacted] at a single dose in order to inform the clinical drug development plan for these interventions. Alternative doses, routes of administration CCI [redacted] and/or treatment combinations may also be assessed via the addition of treatment arms. Up to 100 additional participants may be introduced either for each optional treatment arm or in addition to an existing treatment arm (including placebo).

The follow-up at Day 85 adequately covers the duration for immune response for CCI [redacted] and Arms 9-14. CCI [redacted]

**Participant Characteristics**

The participant population are those infected with SARS-CoV-2 that have developed symptoms consistent with COVID-19. There is historical evidence that patients infected with upper respiratory viruses who are treated early in their disease course have better responses to anti-viral therapies (Aoki et al. 2003). This hypothesis will be tested with a focused subgroup analysis on participants who received intervention within 8 days of symptom onset and using a virology endpoint (see Section 3).

*Low Risk Participants*

The population of participants with mild to moderate COVID-19 illness was chosen to evaluate if effective antiviral antibody therapy may prevent progression to the severe form of COVID-19 illness by treating this population early in their disease course and prior to respiratory compromise and failure.

*High Risk Participants*

The population of participants in treatment arms 12-14 are required to have at least 1 risk factor for developing severe COVID-19 illness (see Section 5.1, Inclusion Criteria 27 and 28 for treatment arms 12 and 13, and Inclusion Criterion 30 for treatment arm 14). The risk factors were based on the Centers for Disease Control guidance (CDC 2021). Participants with these risk factors are at higher risk for more severe disease and hospitalization.

*Adolescent Participants*

There are no approved treatments for adolescents infected with SARS-CoV-2 or to prevent infection in adolescents with comorbidities that place them at increased risk should they become exposed to SARS-CoV-2. Adolescents with mild or moderate COVID-19 at higher risk for severe disease and hospitalization are included in this study.

To minimize invasive procedures and blood volume collection concerns in adolescents, certain laboratory tests and sample collections are excluded for this population.

**Interim Reviews**

The interim safety and efficacy reviews will inform the clinical drug development plans for [redacted] LY3853113 alone, [redacted] LY3853113 in combination with LY3819253 and LY3832479, [redacted]

#### 4.3. Justification for Dose

**LY3819253**

LY3819253 700 mg was estimated as the maximum therapeutic dose based on PK/PD viral dynamics modeling and has a sustained concentration above the in vitro IC90 of viral cell-entry neutralization in the lung tissue for at least 28 days in 90% of the participant population. [redacted]

**LY3832479**

LY3832479 1400 mg dose was selected as the maximum therapeutic dose based on PK/PD viral dynamics modeling and has a sustained concentration above the in vitro IC90 of viral cell-entry neutralization in the lung tissue for at least 28 days in 90% of the participant population. [redacted]

[redacted]

**LY3853113 alone or in combination with LY3819253 and LY3832479**

The intravenous target therapeutic doses of LY3853113 (175 mg), LY3819253 (700 mg) and LY3832479 (1400 mg) were selected individually using PK/PD modeling. The PK/PD modeling approach includes in vitro potency data (i.e. IC90), predicted human PK, and the expected response in terms of maximal reduction in viral load. For each drug separately, the intention was to identify a dose that results in drug concentration above IC90 in at least 90% of patients for at least 28 days, and that also results in maximum viral load reduction based on a PK-viral dynamic model. Also, 700 mg of LY3819253 and 1400 mg of LY3832479 are the currently authorized doses.

**[redacted], LY3853113 + LY3819253 + LY3832479 Combination, [redacted]**

Arms [redacted] 9-10, 12-14: To provide coverage of the different epitopes on SARS-CoV-2 receptor binding domain sites, the dose selection rationale for each single mAb intervention in the combination is the same as for the individual dose rationale for a single mAb intervention.

Optional Arm(s): The dose levels for optional arms and any additional arms will be determined based on interim analysis results in the study and/or other emerging data.

#### 4.4. End of Study Definition

A participant is considered to have completed the study if they have completed all required phases of the study including the last scheduled procedure shown in the SoA (Section 1.3).

The end of the study is defined as the date of last scheduled procedure shown in the SoA (Section 1.3) for the last participant in the trial globally.

### 5. Study Population

Prospective approval of protocol deviations to recruitment and enrollment criteria, also known as protocol waivers or exemptions, is not permitted.

Due to the criticality of participant health, verbal interview of the potential participant, or their legal representative or family member, may be the source for disease characteristics and medical history, unless otherwise specified within the eligibility criteria.

#### 5.1. Inclusion Criteria

Participants are eligible to be included in the study only if all of the following criteria apply:

**Age**

1. For low-risk participant arms only: Are ≥18 and <65 years of age at the time of randomization and do not have the risk factors defined in Criterion 27

27. For high-risk participant arms 12 and 13 only:
    a. Are ≥18 years of age and satisfy at least one of the following risk factors at the time of screening
        i. Are ≥ 65 years of age
        ii. Have a BMI ≥ 35
        iii. Have chronic kidney disease
        iv. Have type 1 or type 2 diabetes
        v. Have immunosuppressive disease
        vi. Are currently receiving immunosuppressive treatment, or
        vii. Are ≥ 55 years of age AND have
            1. cardiovascular disease, OR
            2. hypertension, OR
            3. chronic obstructive pulmonary disease or other chronic respiratory disease

28. For high-risk participant arms 12 and 13 only:
    a. Are 12-17 years of age (inclusive) AND satisfy at least one of the following risk factors at the time of screening
        i. Have a BMI ≥85$^{th}$ percentile for their age and gender based on CDC growth charts (CDC 2017)
        ii. Have sickle cell disease
        iii. Have congenital or acquired heart disease
        iv. Have neurodevelopmental disorders, for example, cerebral palsy
        v. Have a medical-related technological dependence, for example, tracheostomy, gastrostomy, or positive pressure ventilation (not related to COVID-19)
        vi. Have asthma or reactive airway or other chronic respiratory disease that requires daily medication for control
        vii. Have type 1 or type 2 diabetes
        viii. Have chronic kidney disease

   ix. Have immunosuppressive disease, or
   x. Are currently receiving immunosuppressive treatment.

30. For high-risk participant arm 14 only:
   a. Are ≥12 years of age and satisfy at least one of the following risk factors at the time of screening
      i. Are ≥65 years of age
      ii. Are adults (≥18 years of age) with BMI >25 kg/m$^2$, or if age 12-17, have BMI ≥85th percentile for their age and gender based on CDC growth charts (CDC 2017)
      iii. Have chronic kidney disease
      iv. Have type 1 or type 2 diabetes
      v. Have immunosuppressive disease
      vi. Are currently receiving immunosuppressive treatment
      vii. Have cardiovascular disease (including congenital heart disease) or hypertension
      viii. Have chronic lung diseases (for example, chronic obstructive pulmonary disease, asthma [moderate-to-severe], interstitial lung disease, cystic fibrosis and pulmonary hypertension)
      ix. Have sickle cell disease
      x. Have a neurodevelopmental disorder (for example, cerebral palsy) or other conditions that confer medical complexity (for example, genetic or metabolic syndromes and severe congenital anomalies), or
      xi. Have a medical-related technological dependence (for example, tracheostomy, gastrostomy, or positive pressure ventilation [not related to COVID-19])

**Disease Characteristics**

2. Are currently not hospitalized
3. Have one or more mild or moderate COVID-19 symptoms (CDC 2020; FDA 2021 )
      i. Fever
      ii. Cough
      iii. Sore throat
      iv. Malaise
      v. Headache
      vi. Muscle pain
      vii. Gastrointestinal symptoms, or
      viii. Shortness of breath with exertion
      ix. Nasal congestion or runny nose
      x. New loss of smell
      xi. Chills
4. Must have sample collection for first positive SARS-CoV-2 viral infection determination ≤3 days prior to start of the infusion

**Sex**

5. Are men or non-pregnant women
   Reproductive and Contraceptive agreements and requirements are provided in Section 10.4, Appendix 4. Contraceptive use by men or women should be consistent with local regulations for those participating in clinical studies.

**Study Procedures**

6. Understand and agree to comply with planned study procedures
7. Agree to the collection of nasopharyngeal swabs and venous blood

**Informed Consent**

8. The participant or legally authorized representative gives signed informed consent and/or assent as described in Section 10.1.3 which includes compliance with the requirements and restrictions listed in the informed consent form (ICF) and in this protocol

#### 5.2. Exclusion Criteria

Participants are excluded from the study if any of the following criteria apply:

**Medical Conditions**

9. For low-risk participants only: Have a BMI ≥35
   Note: BMI is rounded to the nearest whole number, for example, 34.5 is rounded to 35.
10. Have SpO2 ≤ 93% on room air at sea level or PaO2/FiO2 <300, respiratory rate ≥30 per minute, heart rate ≥125 per minute (FDA resource page, WWW)
11. Require mechanical ventilation or anticipated impending need for mechanical ventilation
12. Have known allergies to any of the components used in the formulation of the interventions
13. Have hemodynamic instability requiring use of pressors within 24 hours of randomization
14. Suspected or proven serious, active bacterial, fungal, viral, or other infection (besides COVID-19) that in the opinion of the investigator could constitute a risk when taking intervention
15. Have any co-morbidity requiring surgery within <7 days, or that is considered life-threatening within 29 days
16. Have any serious concomitant systemic disease, condition or disorder that, in the opinion of the investigator, should preclude participation in this study

**Other Exclusions**

17. Have a history of a positive SARS-CoV-2 serology test
18. Have a history of a positive SARS-CoV-2 test prior to the one serving as eligibility for this study
19. Have received an investigational intervention for SARS-CoV-2 prophylaxis within 30 days before dosing
20. Have received treatment with a SARS-CoV-2 specific monoclonal antibody

21. Have received convalescent COVID-19 plasma treatment
22. For low-risk arms only: Have received a SARS-CoV-2 vaccine OR have participated in a previous SARS-CoV-2 vaccine study and are currently blinded to treatment allotment
23. Have participated, within the last 30 days, in a clinical study involving an investigational intervention. If the previous investigational intervention has a long half-life, 5 half-lives or 30 days, whichever is longer, should have passed.
24. Are concurrently enrolled in any other type of medical research judged not to be scientifically or medically compatible with this study
25. Are pregnant or breast feeding
26. Are investigator site personnel directly affiliated with this study
29. Have body weight <40 kg.

#### 5.3. Lifestyle Considerations

Reproductive and Contraceptive guidance is provided in Section 10.4, Appendix 4.

#### 5.4. Screen Failures

Screen failures are defined as participants who consent to participate in the clinical study but are not subsequently enrolled in the study. A minimal set of screen failure information is required to ensure transparent reporting of screen failure participants to meet the Consolidated Standards of Reporting Trials (CONSORT) publishing requirements and to respond to queries from regulatory authorities. Minimal information includes demography, screen failure details, eligibility criteria, and any serious adverse event (SAE).

Individuals who do not meet the criteria for participation in this study (screen failure) may not be rescreened.

### 6. Study Intervention

Study intervention is defined as any investigational intervention(s), marketed product(s), placebo, or medical device(s) intended to be administered to/used by a study participant according to the study protocol.

#### 6.1. Study Intervention(s) Administered

Each participant will receive a single administration of either placebo, [redacted] LY3853113, [redacted] LY3853113 in combination with LY3819253 and LY3832479, [redacted] Study intervention must be administered within 3 days of the first positive SARS-CoV-2 test sample collection.

[redacted]

Abbreviations: IMP = investigational medicinal product.

CCI

[redacted]

Abbreviations: IMP = investigational medicinal product.

**Treatment Arms 9-14**

| **Intervention Name** | Placebo | LY3853113 | LY3853113 + LY3819253 + LY3832479 |
|---|---|---|---|
| **Dose Formulation** | 0.9% sodium chloride solution | Solution | Solution |
| **Dosage Level(s) (mg)** | Not applicable | 175 mg | 175 mg + 700 mg + 1400 mg |
| **Use** | Placebo | Experimental | Experimental |
| **IMP and NIMP** | IMP | IMP | IMP |
| **Sourcing** | Commercially available 0.9% sodium chloride solution | From Lilly | From Lilly |
| **Packaging and Labeling** | Commercially available 0.9% sodium chloride solution | Study Intervention will be provided in glass vials and will be labeled appropriately. | Study Intervention will be provided in glass vials and will be labeled appropriately. |

Abbreviations: IMP = investigational medicinal product.

Infusion information may be found in the pharmacy preparation instructions.

CCI

[redacted]

CCI [redacted]

**Arms 9-14**

Participants should be monitored for at least 1 hour after completion of infusion. The infusion rate may be reduced as deemed necessary if an infusion reaction is observed (Section 6.1.1.2).

Further details will be included in the pharmacy preparation instructions (PPI). The site must have resuscitation equipment, emergency drugs and appropriately training staff available during the infusion and for at least 1 hour after the completion of the infusion.

##### 6.1.1. Special Treatment Considerations

###### 6.1.1.1. Premedication for Infusions

Premedication for infusions is not planned. However, if an infusion reaction occurs during administration or if the participant has a medical history suggesting a potential benefit from premedication, the study investigator(s) should determine the appropriate premedication.

The investigators and sponsor may decide to use premedication if the frequency of infusion reactions among participants warrants it.

If minor infusion reactions are observed, administration of acetaminophen, 500 mg to 1000 mg, antihistamines and/or other appropriately indicated medications may be given prior to the start of infusions for subsequent participants.

The decision to implement premedication for infusions in subsequent participants will be made by the investigator and sponsor and recorded in the study documentation.

Any premedications given will be documented as a concomitant therapy.

###### 6.1.1.2. Management of Infusion Reactions

All participants should be monitored closely, as there is a risk of infusion reaction and hypersensitivity (including anaphylaxis) with any biological agent.

**Symptoms and Signs**

Symptoms and signs that may occur as part of an infusion reaction include, but are not limited to fever, chills, nausea, headache, bronchospasm, hypotension, angioedema, throat irritation, rash including urticaria, pruritus, myalgia, hypoxia, rapid heart rate, fatigue, altered mental status, and dizziness.

Infusion-related reactions severity will be assessed and reported using the Division of AIDS (DAIDS) Table for Grading the Severity of Adult and Pediatric Adverse Events, version 2.1 (July 2017).

This table describes the severity of reactions according to DAIDS.

| Parameter | Mild | Moderate | Severe | Severe and Potentially Life-threatening |
|---|---|---|---|---|
| Acute Allergic Reaction | Localized urticaria (wheals) with no medical intervention indicated | Localized urticaria with intervention indicated<br>OR<br>Mild angioedema with no intervention indicated | Generalized Urticaria<br>OR<br>Angioedema with intervention indicated<br>OR<br>Symptoms of mild bronchospasm | Acute anaphylaxis<br>OR<br>Life-threatening bronchospasm<br>OR<br>Laryngeal edema |
| Cytokine Release Syndrome[a] | Mild signs and symptoms<br>AND<br>Therapy, that is, antibody infusion interruption not indicated | Therapy (that is, antibody infusion) interruption indicated<br>AND<br>Responds promptly to symptomatic treatment<br>OR<br>Prophylactic medications indicated for ≤24 hours | Prolonged severe signs and symptoms<br>OR<br>Recurrence of symptoms following initial improvement | Life-threatening consequences (for example, requiring pressor or ventilator support) |

[a] A disorder characterized by nausea, headache, tachycardia, hypotension, rash, and/or shortness of breath.

Source: Division of AIDS (DAIDS) Table for Grading the Severity of Adult and Pediatric Adverse Events, Version 2.1 (July 2017).

**Site Needs**

The clinical site should have necessary equipment and medications for the management of any infusion reaction, which may include but is not limited to oxygen, IV fluid, epinephrine, acetaminophen and antihistamine.

**Management of Infusion Reactions**

Investigators should determine the severity of the infusion reaction and manage infusion reactions based on standard of care and their clinical judgment. If an infusion reaction occurs, then supportive care should be used in accordance with the signs and symptoms.

If a participant permanently discontinues from study intervention, they should complete AE monitoring and other procedures as stated in the SoA (Section 1.3).

##### 6.1.2. Temporary Stopping Criteria

The Assessment Committee (AC) members individually will review unblinded safety data and meet as described in the AC Charter. The AC will conduct a full safety review before determining if enrollment should be stopped and/or other study parameters should be modified. (see Section 9.6).

This table describes the location of AE-related information in this protocol.

| Topic | Location |
|---|---|
| DAIDS table describing severity of reactions | Section 6.1.1.2 |
| Definition of AEs | Section 10.3.1 |
| Assessment of Intensity/Severity | Section 10.3.3 |

Abbreviations: DAIDS = Division of AIDS.

Changes to the planned dosing schedule must be appropriately documented and communicated with the study personnel and the IRB/IEC before dosing continues.

#### 6.2. Preparation/Handling/Storage/Accountability

The investigator or designee must confirm appropriate storage conditions have been maintained during transit for all study intervention received and any discrepancies are reported and resolved before use of the study intervention.

To protect blinding, the interventions must be prepared by an unblinded site personnel qualified to prepare study intervention who are not involved in any other study-related procedures. Instructions for preparation will be provided by the sponsor.

Only participants enrolled in the study may receive study intervention and only authorized site staff may supply or administer study intervention.

All study intervention must be stored in a secure, environmentally controlled, and monitored (manual or automated) area in accordance with the labeled storage conditions with access limited to the investigator and authorized site staff.

The investigator or designee is responsible for study intervention accountability, reconciliation, and record maintenance (i.e., receipt, reconciliation, and final disposition records).

Further guidance and information for the final disposition of unused study interventions are provided in the pharmacy manual.

#### 6.3. Measures to Minimize Bias: Randomization and Blinding

##### Randomization

All participants will be centrally randomized to study intervention using an Interactive Web Response System (IWRS). Before the study is initiated, the log in information and directions for the IWRS will be provided to each site.

Participants will be stratified by duration since symptom onset to randomization (≤8 days versus >8 days). For treatment arms 12-13, participants will also be stratified by whether they received a SARS-CoV-2 vaccine or not prior to screening.

For arms 1-11, all eligible participants will be randomized, initially following an equal allocation to treatment arms. For arms 12-13, eligible participants will be randomized following a 2:1 allocation to treatment arms. Periodic adjustments to the allocation ratio, informed by planned interim analyses, may be made in an effort to achieve an equal allocation across the treatment arms at the end of enrollment. Given the staggered start of treatment arms, additional participants may be enrolled in existing treatment arms and the allocation ratio may change accordingly. See Section 9.5 for details.

Participants will be assigned to treatment arm 14 after treatment arms 12 and 13 have completed enrollment.

**Blinding**

Treatment arms 1-11 will be blinded. Neither participants, nor investigators, nor the sponsor study team will be aware of treatment assignments prior to the final data base locks at the conclusion of the study. Treatment arms 12-14 will be open label.

This table describes general procedures for unblinding.

| | |
|---|---|
| Unblinding (IWRS) | • Emergency unblinding for adverse events may be performed through the IWRS. All actions resulting in an unblinding event are recorded and reported by the IWRS<br>• In case of an emergency, the investigator has the sole responsibility for determining if unblinding of a participants' intervention assignment is warranted<br>• Participant safety must always be the first consideration in making such a determination. However, the investigator should make all attempts to contact the Medical Monitor in advance of unblinding<br>• If a participant's intervention assignment is unblinded, the sponsor must be notified immediately after breaking the blind even if consultation occurred in advance<br>• The date and reason that the blind was broken must be recorded in the source documentation and case report form. |

Abbreviations: IWRS = interactive web-response system.

If an investigator, site personnel performing assessments, or participant is unblinded while the infusion is ongoing, the participant must be discontinued from the study intervention and the infusion stopped. If any amount of study intervention was administered, follow procedures according to the SoA (Section 1.3).

#### 6.4. Study Intervention Compliance

Participants will receive study intervention directly from the investigator or designee, under medical supervision. The date and time of dosing will be recorded in the source documents and recorded in the CRF. The dose of study intervention and study participant identification will be confirmed at the time of dosing by a member of the study site staff other than the person administering the study intervention.

#### 6.5. Concomitant Therapy

**Prior Treatment**

Any prior therapy, such as antivirals, antibiotics, or anti-malarials used in the prior 30 days to signing informed consent should be recorded.

Any SARS-CoV-2 vaccine should be recorded. Any non-SARS-CoV-2 vaccine received in the prior 90 days to signing informed consent should be recorded.

Therapy prior to enrollment with antivirals including lopinavir/ritonavir, remdesivir, or other therapeutic agents (e.g., corticosteroids) are permitted.

Convalescent COVID-19 plasma treatment is not allowed prior to enrollment.

**Concomitant Therapy**

Participants should be treated according to standard of care, even as it evolves, and medications may be part of the treatment paradigm. Therefore, remdesivir may be initiated as standard of care for hospitalized participants (if available through the FDA Emergency Use Authorizations) outside of local standard of care per written policies or guidelines.

If the local standard of care per written policies or guidelines (that is, not just an individual clinician decision) includes lopinavir/ritonavir, chloroquine, hydroxychloroquine or other investigational agents, then initiating these during the study is permitted, but may require additional safety monitoring by the site.

Convalescent COVID-19 plasma treatment is not allowed.

Any medication, investigational agent, or vaccine, including over the counter or prescription medicines, vitamins, and/or herbal supplements, or other specific categories of interest that the participant is receiving at the time of enrollment or receives during the study must be recorded along with

- Reason for use
- Dates of administration including start and end dates, and
- Dosage information including dose and frequency for concomitant therapy of special interest.

Acetaminophen and corticosteroid use are permitted at any time during the study.

Other concomitant medication may be considered on a case-by-case basis by the investigator in consultation with the sponsor if required.

The sponsor should be contacted if there are any questions regarding concomitant or prior therapy.

#### 6.6. Dose Modification

No dose modifications are planned for this study.

#### 6.7. Intervention after the End of the Study

No continued access is planned after completion of this study, as additional efficacy would be needed to demonstrate continued access criteria.

### 7. Discontinuation of Study Intervention and Participant Discontinuation/Withdrawal

Discontinuation of specific sites or of the trial as a whole are handled as part of regulatory, ethical, and trial oversight considerations in Section 10.1.9, Appendix 1.

#### 7.1. Discontinuation of Study Intervention

In rare instances, it may be necessary for a participant to permanently discontinue (definitive discontinuation) study intervention. If the study treatment is definitively discontinued, the participant will remain in the study for the remainder of the assessment visits through Day 29 and also for the post-treatment follow-up visits on Days 60 and 85 for all treatment arms CCI [redacted]

#### 7.2. Participant Discontinuation/Withdrawal from the Study

A participant may withdraw from the study

- at any time at his/her own request
- at the request of his/her designee (for example, parents or legal guardian)
- at the discretion of the investigator for safety, behavioral, compliance, or administrative reasons
- if the participant becomes pregnant during the study, or
- if enrollment in any other clinical study involving an investigational product or enrollment in any other type of medical research judged not to be scientifically or medically compatible with this study.

If the participant, for any reason, requires treatment with another therapeutic agent that has been demonstrated to be effective for treatment of the study indication, discontinuation from the study occurs prior to introduction of the new agent.

Discontinuation is expected to be uncommon.

At the time of discontinuation, if possible, an early discontinuation visit should be conducted as described in the SoA (Section 1.3). The participant should also return for the post-treatment follow-up visits.

If the participant discontinues on the same day as a normally scheduled visit, only one set of laboratory tests, sample collection and assessments are collected.

If the participant withdraws consent for disclosure of future information, the sponsor may retain and continue to use any data collected before such a withdrawal of consent. If a participant withdraws from the study, they may request destruction of any samples taken and not tested, and the investigator must document this in the site study records.

##### 7.2.1. Discontinuation of Inadvertently Enrolled Participants

If the sponsor or investigator identify a participant who did not meet enrollment criteria and was inadvertently enrolled, then the participant should be discontinued from study intervention unless there are extenuating circumstances that make it medically necessary for the participant to continue study intervention.

If the investigator and the sponsor CRP agree it is medically appropriate to continue, the investigator must obtain documented approval from the sponsor CRP to allow the inadvertently enrolled participant to continue in the study with or without treatment with investigational product.

Safety follow-up is as outlined in

- Section 1.3 (Schedule of Activities)
- Section 8.2 (Safety Assessments), and
- Section 8.3 (Adverse Events and Serious Adverse Events).

#### 7.3. Lost to Follow up

A participant will be considered lost to follow-up if they repeatedly fail to return for scheduled visits and is unable to be contacted by the study site. Site personnel are expected to make diligent attempts to contact participants who fail to return for a scheduled visit or were otherwise unable to be followed up by the site.

Site personnel, or an independent third party, will attempt to collect the vital status of the participant within legal and ethical boundaries for all participants that received study intervention. Public sources may be searched for vital status information. If vital status is determined to be deceased, this will be documented and the participant will not be considered lost to follow-up. Sponsor personnel will not be involved in any attempts to collect vital status information.

### 8. Study Assessments and Procedures

Study procedures and their timing are summarized in the SoA (Section 1.3).

Protocol waivers or exemptions are not allowed.

Monitoring of blinded safety data will continue throughout the study and will be conducted by blinded study team members. Details of the blinded safety reviews, including the frequency and approximate timing, are specified in the trial level safety review plan.

Immediate safety concerns should be discussed with the sponsor immediately upon occurrence or awareness to determine if the participant should continue or discontinue study intervention.

Adherence to the study design requirements, including those specified in the SoA, is essential and required for study conduct.

All screening evaluations must be completed and reviewed to confirm that potential participants meet all eligibility criteria. The investigator will maintain a screening log to record details of all participants screened and to confirm eligibility or record reasons for screening failure, as applicable.

#### 8.1. Efficacy Assessments

Hospitalization events (Section 8.2.4), procedures of special interest (Section 8.2.5), vital signs (Section 8.2.2) and symptomology (Section 8.1.1) will be used to characterize the effect of CCI [redacted], LY3853113 alone, CCI [redacted], LY8353113 in combination with LY3819253 and LY3832479, CCI [redacted] compared to placebo on clinical status from baseline to Days 3, 5, 7, 11, 22 and 29.

##### 8.1.1. Symptoms and Overall Clinical Status Participant Questionnaire

Participants will rate their overall clinical status and severity of symptoms associated with COVID-19 by a daily questionnaire. This questionnaire is for outpatients only.

Participants will complete three questions about their overall clinical status daily, including

- severity of symptoms
- general physical health, and
- change in overall health.

The questionnaire contains these symptoms

- cough
- shortness of breath
- feeling feverish
- fatigue
- body aches and pain
- sore throat
- chills
- headache

- loss of appetite, and
- changes in taste and smell.

Each symptom will be scored daily by the participant as experienced during the past 24 hours.

| Rating | Score |
|---|---|
| None or absent | 0 |
| Mild | 1 |
| Moderate | 2 |
| Severe | 3 |

Participants will rate changes in taste and smell with a yes/no response.

#### 8.2. Safety Assessments

Planned time points for all safety assessments are provided in the SoA (Section 1.3).

##### 8.2.1. Physical Examinations

A complete physical examination will be performed at the screening visit. This examination excludes pelvic, rectal, and breast examinations unless clinically indicated.

A symptom-directed physical examination will be performed at other visits, as specified in the SoA and as clinically indicated.

Investigators should pay special attention to clinical signs related to COVID-19, to ongoing medical conditions and to previous serious illnesses.

##### 8.2.2. Vital Signs

Vital signs will be measured as specified in the SoA (Section 1.3) and as clinically indicated. Vital signs include

- Body temperature
- Blood pressure
- Pulse rate
- Respiration rate
- Saturation of peripheral oxygen, and
- Supplemental oxygen flow rate, FiO2 if known, and method of delivery, if applicable.

These tables outline Day 1 vital signs data collection on the CRF in relation to the infusion(s). Infusion times may vary depending on the participant.

[redacted]

*Arms 9-11, 13, 14*

| **Timepoint (minutes)** | **Collect data on CRF** |
|---|---|
| Immediately before infusion | Yes |
| If infusion is <15 minutes, immediately following completion of infusion | Yes |
| During Infusions > 15 minutes, as possible (if applicable) | -- |
| 15 | No |
| 30 | Yes |
| 45 | No |
| 60 | Yes |
| After infusion – every 30 minutes for 1 hour after the end of the infusion | -- |
| end of infusion +30 minutes | Yes |
| end of infusion +60 minutes | No |

Additional vital signs may be measured during the study if warranted, as determined by the investigator.

##### 8.2.3. Clinical Laboratory Assessments

See Section 10.2, Appendix 2 for the list of clinical laboratory tests to be performed and the SoA (Section 1.3) for the timing and frequency. All protocol-required laboratory assessments, as defined in Appendix 2, must be conducted in accordance with the laboratory manual and the SoA.

Clinically significant abnormal laboratory findings are those which are not associated with the underlying disease, unless judged by the investigator to be more severe than expected for the participant's condition.

All laboratory tests with values considered clinically significantly abnormal during participation in the study or within 28 days after the dose of study intervention should be repeated until the values return to normal or baseline or are no longer considered clinically significant by the investigator or sponsor.

If such values do not return to normal/baseline within a period of time judged reasonable by the investigator, the etiology should be identified and the sponsor notified.

The investigator must review the laboratory report, document this review, and record any clinically relevant changes occurring during the study in the AE section of the CRF.

The laboratory reports must be filed with the source documents.

All protocol-required laboratory assessments, as defined in Appendix 2, must be conducted in accordance with the laboratory manual and the SoA (Section 1.3).

If laboratory values from non-protocol specified laboratory assessments performed at the institution’s local laboratory require a change in participant management or are considered clinically significant by the investigator (e.g., SAE or AE or dose modification), report in the AE section of the CRF.

**Pregnancy Testing**

Women of childbearing potential (WOCBP) must undergo pregnancy testing according to the SoA. Participants who are pregnant will be discontinued from the study.

##### 8.2.4. Hospitalization events

If a participant is admitted to the hospital, the participant will remain in the study and follow the procedures outlined in the SoA (Section 1.3). Hospitalization is defined as ≥24 hours of acute care.

The date of hospitalization events will be recorded in the CRF and includes

- hospitalization
- emergency room visit
- ICU admittance
- Extended care facility admittance, and
- Discharge.

##### 8.2.5. Procedures of Special Interest

The participants’ clinical status and concurrent procedures of special interest will be recorded in the CRF and include consciousness status using the alert, consciousness, verbal, pain, unresponsive scale (ACVPU), limitation on activities due to COVID-19 using the patient global assessment for daily activities of physical function, and requirements for

- ongoing hospital medical care
- supplemental oxygen
- non-invasive ventilation or a high flow oxygen device
- mechanical ventilation
- ECMO, or
- additional organ support (e.g., pressors, renal replacement).

##### 8.2.6. Respiratory Support

Once enrolled in the study, participants may be managed with high-flow nasal cannula, noninvasive positive pressure ventilation or any other form respiratory support as needed per investigator discretion.

#### 8.3. Adverse Events and Serious Adverse Events

AEs will be reported by the participant, or, when appropriate, by a caregiver, surrogate, or the participant's legally authorized representative.

The investigator and any qualified designees are responsible for detecting, documenting, and recording events that meet the definition of an AE or SAE and remain responsible for following up AEs that are serious, considered related to the study intervention or study procedures, or that caused the participant to discontinue the study intervention or study.

##### 8.3.1. Time Period and Frequency for Collecting AE and SAE Information

All AEs and SAEs will be collected from the time of signing of the informed consent form (ICF) until participation in study has ended.

Adverse events that begin before the start of study intervention but after signing of the ICF will be recorded on the Adverse Event CRF.

Although all AEs after signing the ICF are recorded by the site in the CRF/electronic data entry, SAE reporting to sponsor begins after the participant has signed the ICF and has received study intervention. However, if an SAE occurs after signing the ICF, but prior to receiving study intervention, it needs to be reported within the SAE reporting timeframe if it is considered reasonably possibly related to study procedures.

All SAEs will be recorded and reported to the sponsor or designee immediately and under no circumstance should this exceed 24 hours, as indicated in Section 10.3, Appendix 3. The investigator will submit any updated SAE data to the sponsor within 24 hours of it being available.

Investigators are not obligated to actively seek AEs or SAEs after conclusion of the study participation. However, if the investigator learns of any SAE, including a death, at any time after a participant has been discharged from the study, and he/she considers the event to be reasonably related to the study intervention or study participation, the investigator must promptly notify the sponsor.

##### 8.3.2. Method of Detecting AEs and SAEs

The method of recording, evaluating, and assessing causality of AE and SAE and the procedures for completing and transmitting SAE reports are provided in Section 10.3.

Care will be taken not to introduce bias when detecting AEs and/or SAEs. Open-ended and non-leading verbal questioning of the participant is the preferred method to inquire about AE occurrences.

##### 8.3.3. Follow-up of AEs and SAEs

After the initial AE/SAE report, the investigator is required to proactively follow each participant at subsequent visits/contacts. All SAEs will be followed until resolution, stabilization, the event is otherwise explained, or the participant is lost to follow-up (as defined in Section 7.3). Further information on follow-up procedures is provided in Section 10.3.

##### 8.3.4. Regulatory Reporting Requirements for SAEs

Prompt notification by the investigator to the sponsor of a SAE is essential so that legal obligations and ethical responsibilities towards the safety of participants and the safety of a study intervention under clinical investigation are met.

The sponsor has a legal responsibility to notify both the local regulatory authority and other regulatory agencies about the safety of a study intervention under clinical investigation. The sponsor will comply with country-specific regulatory requirements relating to safety reporting to the regulatory authority, Institutional Review Boards (IRB)/Independent Ethics Committees (IEC), and investigators.

An investigator who receives an investigator safety report describing a SAE or other specific safety information (e.g., summary or listing of SAEs) from the sponsor will review and then file it with the IB and will notify the IRB/IEC, if appropriate according to local requirements.

##### 8.3.5. Pregnancy

Details of all pregnancies in female participants will be collected.

If a pregnancy is reported, the investigator should inform the sponsor within 24 hours of learning of the pregnancy and should follow the procedures outlined in Section 10.4, Appendix 4.

Abnormal pregnancy outcomes, for example, spontaneous abortion, fetal death, stillbirth, congenital anomalies, and ectopic pregnancy, are considered SAEs.

##### 8.3.6. Hypersensitivity Reactions

Many drugs, but particularly biologic agents, carry the risk of systemic hypersensitivity reactions.

If such a reaction occurs, additional details describing each symptom should be provided to the sponsor in the infusion-related reaction/hypersensitivity CRF.

If symptoms and/or signs occur during or within 6 hours after infusion and are believed to be hypersensitivity or due to cytokine release, then investigators are encouraged to report the event as infusion-related immediate hypersensitivity reaction or cytokine release-associated infusion reaction, respectively.

Sites should have appropriately trained medical staff and appropriate medical equipment available when study participants are receiving intervention. It is recommended that participants who experience a systemic hypersensitivity reaction be treated per the local standard of care.

In the case of generalized urticaria or anaphylaxis, additional blood and urine samples should be collected as described in Section 10.6, Appendix 6, “Recommended Laboratory Testing for Hypersensitivity Events”. Laboratory results are provided to the sponsor via the central laboratory.

In case of skin lesions or rash consistent with vasculitis, efforts should be made to perform the following as soon as possible

- dermatology consultation
- skin biopsy, and
- photographs of lesions of interest, including skin biopsy site.

##### 8.3.7. Infusion-related Reactions

As with other mAbs, infusion-related reactions may occur during or following IV administration. If an infusion-related reaction occurs, additional data describing each symptom and sign should be provided to the sponsor in the CRF.

This table describes the location of infusion-related reaction information in this protocol.

| Topic | Location |
|---|---|
| Special treatment considerations | Section 6.1.1 |
| Premedication for infusions | Section 6.1.1.1 |
| Management of infusion reactions | Section 6.1.1.2 |
| DAIDS table describing severity | Section 6.1.1.2 |
| Treatment guidelines for infusion-related reactions | Section 6.1.1.2 |

Symptoms occurring during or after infusion of study intervention may also be defined according to AE categories such as acute allergic reaction or cytokine release syndrome (refer to DAIDS).

##### 8.3.8. Product Complaints

A product complaint is any written, electronic, or oral communication that alleges deficiencies related to the identity, quality, durability, reliability, safety, effectiveness or performance of a trial intervention.

Sponsor collects product complaints on study intervention and drug delivery systems used in clinical studies in order to ensure the safety of study participants, monitor quality, and to facilitate process and product improvements.

Participants will be instructed to contact the investigator as soon as possible if they have a complaint or problem with the intervention so that the situation can be assessed.

NOTE: AEs/SAEs that are associated with a product complaint will also follow the processes outlined in Section 8.3.3 and Appendix 10.3 of the protocol.

**Time Period for Detecting Product Complaints**

Product complaints that result in an adverse event will be detected, documented, and reported to the sponsor during all periods of the study in which the intervention is used.

If the investigator learns of any product complaint at any time after a participant has been discharged from the study, and such incident is considered reasonably related to an intervention provided for the study, the investigator will promptly notify the sponsor.

**Prompt Reporting of Product Complaints to Sponsor**

Product complaints will be reported to the sponsor within 24 hours after the investigator becomes aware of the complaint.

The Product Complaint Form will be sent to the sponsor by a method designated by the sponsor.

**Follow-up of Product Complaints**

Follow-up applies to all participants, including those who discontinue study intervention.

The investigator is responsible for ensuring that follow-up includes any supplemental investigations as indicated to elucidate the nature and/or causality of the product complaint.

New or updated information will be recorded on the originally completed form with all changes signed and dated by the investigator and submitted to the sponsor.

#### 8.4. Treatment of Overdose

There is no known antidote for an overdose of [redacted], LY3853113 alone, [redacted], LY3853113 in combination with LY3819253 and LY3832479, [redacted]

In the event of an overdose, the investigator should

1. Contact the sponsor immediately.
2. Closely monitor the participant for any AE/SAE and laboratory abnormalities
3. Provide supportive care as necessary, and
4. Document the quantity of the excess dose in the CRF.

Decisions regarding infusion interruptions or modifications will be made by the investigator, in consultation with the sponsor, based on the clinical evaluation of the participant.

#### 8.5. Pharmacokinetics

Venous blood samples will be collected as specified in the SoA (Section 1.3) for determination of concentrations of LY3819253, LY3832479, LY3853113, and [redacted] used to evaluate the PK for LY3819253, LY3832479, LY3853113, and [redacted]

A maximum of 3 samples may be collected at additional time points during the study if warranted and agreed upon between the investigator and the sponsor.

Instructions for the collection and handling of biological samples will be provided by Lilly.

Site personnel will record

- The date and time (24-hour clock time) of administration (start and end of infusion), and
- The date and time (24-hour clock time) of each PK sample.

It is essential that the times are recorded accurately.

##### 8.5.1. Bioanalytical

Samples will be analyzed at a laboratory approved by the sponsor and stored at a facility designated by the sponsor. Concentrations of LY3819253, LY3832479, LY3853113, and CCI [redacted] will be assayed using a validated bioanalytical method. Analyses of samples collected from placebo-treated participants are not planned.

Sample retention is described in Appendix 1, Section 10.1.12. Remaining samples used for PK may be pooled and used for exploratory metabolism or bioanalytical method experiments as deemed appropriate.

#### 8.6. Pharmacodynamics

The SARS-CoV-2 viral RNA level and viral clearance will be evaluated by nasopharyngeal swabs. See Section 10.2 Clinical Laboratory Tests, and Section 1.3, the SoA for sample collection information.

Sample retention is described in Appendix 1, Section 10.1.12. Remaining samples may be used for additional exploratory studies to better understand LY3819253, LY3832479, LY3853113, CCI [redacted] and the disease, which may include sequencing and/or culture of the virus for future studies.

#### 8.7. Genetics

A whole blood sample will be collected for pharmacogenetic analysis where local regulations allow.

See Section 10.2, Clinical Laboratory Tests, and Section 1.3, the SoA for sample collection information.

See Section 10.5 for genetic research, custody, and sample retention information.

#### 8.8. Biomarkers

Blood samples will be collected from all participants for analysis of immune system-related markers. Serum, whole blood for cellular and/or epigenetic analysis, and whole blood RNA samples for exploratory biomarker research will be collected at the time specified in the SoA (Section 1.3) where local regulations allow.

Samples will be stored and analysis may be performed on biomarkers thought to play a role in immune system related responses to viral infection including, but not limited to, immune pathways, cellular composition, serum analytes, or epigenetic biomarkers, to evaluate their association with observed clinical responses to LY3819253, LY3832479, LY3853113, CCI [redacted] and the disease state.

Samples may be used for research to develop methods, assays, prognostics and/or companion diagnostics related to the intervention target (S protein), the COVID-19 disease state, pathways associated with disease, and/or the mechanism of action of the study intervention.

Sample retention is described in Section 10.1.12.

#### 8.9. Immunogenicity Assessments

**Visits and times**

At the visits and times specified in the SoA (Section 1.3), venous blood samples will be collected to determine antibody production against LY3819253, LY3832479, LY3853113 and/or CCI [redacted]. The actual date and time (24-hour clock time) of each sample collection will be recorded.

**Sample collection, handling, and use**

Instructions for the collection and handling of blood samples will be provided by the sponsor.

Immunogenicity may be assessed by a validated assay designed to detect ADAs in the presence of LY3819253, LY3832479, LY3853113, and/or CCI [redacted] at a laboratory approved by the sponsor. Antibodies may be further characterized for their ability to neutralize the activity of LY3819253, LY3832479, LY3853113, and/or CCI [redacted].

**Sample retention**

Sample retention is described in Appendix 1, Section 10.1.12.

#### 8.10. Health Economics

This section is not applicable for this study.

### 9. Statistical Considerations

#### 9.1. Statistical Hypotheses

[redacted]

[redacted]

For treatment arms 9-11, the null hypothesis for the primary endpoint is that there is no difference between LY3853113 alone or in combination with LY3819253 and LY3832479, versus placebo participants in the proportion of participants with SARS-CoV-2 viral load greater than 5.27 on Day 7.

#### 9.2. Sample Size Determination

**Sample Size**

[redacted]

CCI [redacted]

*Treatment arms 9-11*

The planned sample size is approximately 122 participants per treatment arm.

Sample size was determined based on pairwise comparisons of each dose compared to placebo. An assumed sample size of 122 participants per treatment arm provides about 84% power to test the superiority of either LY3853113 alone or the combination with LY3819253 and LY3832479 versus placebo at the two-sided 0.05 alpha level on the proportion of participants with SARS-CoV-2 viral load greater than 5.27 at Day 7. This assumes the true underlying proportion of participants meeting this endpoint is 12% in both the LY3853113 alone treatment arm and the LY3853113 + LY3819253 + LY3832479 combination treatment arm, and 28% in the placebo arm.

*Treatment arms 12-14*

Approximately 100 participants will be enrolled into treatment arm 12 and approximately 50 participants will be enrolled into treatment arm 13. Treatment arm 12 includes a greater number of participants to assess the tolerability of undiluted LY3853113 administration. Approximately 140 participants will be enrolled into treatment arm 14 once treatment arms 12 and 13 have completed enrollment.

**Stratification**

*Treatment arms 1-13*

Participants will be stratified by duration since symptom onset category (≤8 days versus >8 days).

*Treatment arms 12-13*

Participants will be stratified by whether a participant received a SARS-CoV-2 vaccine or not prior to screening.

#### 9.3. Populations for Analyses

This table defines the populations for analysis.

| Population | Description |
|---|---|
| Entered | All participants who sign the informed consent form |
| Efficacy | All randomized participants who received study intervention and provided at least one post-baseline measure for the relevant endpoint. Participants will be analyzed according to the intervention to which they were randomized. (Intention to treat). |
| Safety | All participants randomly assigned and who received study intervention. Participants will be analyzed according to the intervention they actually received. |
| Pharmacokinetic | All randomized participants who received study intervention and have evaluable PK sample. Participants will be analyzed according to the intervention they received. |

#### 9.4. Statistical Analyses

Statistical analysis of this study will be the responsibility of the Sponsor or its designee.

Unless otherwise specified, treatment effects will be conducted using 2-sided tests at an alpha level of 0.05. No adjustment for multiplicity will be performed in this study.

Analyses will be performed separately for treatment arms

CCI

- 9-11,
- 12 and 13, and
- 14.

For treatment arms 12 and 13, participants who have received a SARS-CoV-2 vaccine prior to screening may be summarized separately from those who have not received a SARS-CoV-2 vaccine prior to screening, for all efficacy outcomes.

Details of the handling of dropouts or missing data will be fully described in the statistical analysis plan (SAP).

Any change to the data analysis methods described in the protocol will require an amendment only if it changes a principal feature of the protocol. Any other change to the data analysis methods described in the protocol, and the justification for making the change, will be described in the SAP. Additional exploratory analyses of the data will be conducted as deemed appropriate.

The SAP will be finalized prior to the first final database lock (i.e., first unblinding of the sponsor), and it will include a more technical and detailed description of the statistical analyses described in this section. This section is a summary of the planned statistical analyses of the most important endpoints including primary and secondary endpoints.

##### 9.4.1. General Considerations

This table describes the general statistical methods that may be used in this study.

| Method | Analysis |
|---|---|
| Descriptive Statistics | number of participants, mean, standard deviation, median, minimum, and maximum for continuous measures, and frequency counts and percentages for categorical measures |
| Kaplan-Meier curves and summary statistics | Treatment comparisons of time-to-event based endpoints |
| Logistic regression analysis | Treatment comparisons of binary variables with treatment and randomization stratification variables in the model. |
| Nonparametric<br>(for example, Mann-Whitney or van Elteren tests) | Treatment comparison of ordinal, nominal and non-normally distributed continuous variables. |

Additional statistical methodology, sensitivity analyses accounting for missing data, and adjustments for covariates, if any, will be described in the SAP.

##### 9.4.2. Primary Endpoints

*Treatment Arms 1-11*

The primary endpoint is the proportion of participants with SARS-CoV-2 viral load greater than 5.27 on Day 7, corresponding to Ct value of 27.5 based on nasopharyngeal swab sampling for reverse transcription polymerase chain reaction (RT-PCR) testing for SARS-CoV-2.

Statistical hypotheses testing for the primary endpoint will be conducted using a logistic regression model at the two-sided 0.05 level.

Full details will be provided in the SAP.

*Treatment Arms 12-14*

All investigational product and protocol procedure AEs will be listed, and if the frequency of events allows, safety data will be summarized using descriptive methodology.

The incidence of AEs and SAEs for each treatment will be presented by severity and by association with intervention as perceived by the investigator. Adverse events reported prior to randomization will be distinguished from those reported as new or increased in severity during the study post-randomization.

Safety parameters that will be assessed include, but are not limited to, safety laboratory parameters, and vital signs. The parameters will be listed and summarized using standard descriptive statistics. Additional analysis will be performed if warranted upon review of the data.

##### 9.4.3. Secondary Endpoints

###### 9.4.3.1. Safety (Arms 1-11)

Safety analyses will be conducted using the safety population described above.

All investigational product and protocol procedure AEs will be listed, and if the frequency of events allows, safety data will be summarized using descriptive methodology.

The incidence of AEs and SAEs for each treatment will be presented by severity and by association with intervention as perceived by the investigator. Adverse events reported prior to randomization will be distinguished from those reported as new or increased in severity during the study post-randomization.

Safety parameters that will be assessed include, but are not limited to, safety laboratory parameters, and vital signs. The parameters will be listed and summarized using standard descriptive statistics. Additional analysis will be performed if warranted upon review of the data.

###### 9.4.3.2. Additional Secondary Endpoints

Additional secondary endpoints include

- Proportion (percentage) of participants who experience these events by Day 29
    - COVID-19 related hospitalization (defined as ≥ 24 hours of acute care), or
    - death.
- SARS-CoV-2 viral load change from baseline to
    - Day 3
    - Day 5
    - Day 7
    - Day 11
- Proportion of participants with viral load greater than 5.27 on Day 7 among participants enrolled with ≤ 8 days of symptoms prior to randomization
- Proportion of participants that achieve SARS-CoV-2 clearance (Days 3, 5, 7, 11, and 29)
- Time to SARS-CoV-2 clearance
- SARS-CoV-2 viral load AUC assessed through Day 11.
- 75$^{th}$ percentile of SARS-CoV-2 viral load at Day 7
- Time to symptom resolution
    - symptoms are scored as absent
- Proportion of participants demonstrating symptom resolution via the symptom questionnaire on Days 3, 5, 7, 11, 22, and 29
- AUC from baseline to Day 11
- Change in symptom score (total of ratings) from baseline up to Days 7, 11, 22, and 29
- Time to symptom improvement
    - symptoms scored as moderate or severe at baseline are scored as mild or absent, AND
    - symptoms scored as mild or absent at baseline are scored as absent.
- Proportion of participants demonstrating symptom improvement via the symptom questionnaire on Days 3, 5, 7, 11, 22, and 29
- Proportion (percentage) of participants who experience these events by Day 22
    - COVID-19 related hospitalization (defined as ≥ 24 hours of acute care)
    - death.
- Proportion (percentage) of participants who experience these events through Day 29:
    - COVID-19 related hospitalization (defined as ≥24 hours of acute care),

    - COVID-19 related emergency room visit, or
    - death.
- Characterize the PK of LY3819253, LY3832479, LY3853113, and [redacted]
    - [redacted]
    - Mean concentration of LY3853113 in combination with LY3819253 and LY3832479 on Day 29

Full details of the analyses will be in the SAP.

###### 9.4.3.3. Pharmacokinetic Analyses

Pharmacokinetic analyses will be conducted on data from all participants who receive intervention and have evaluable PK.

Concentration-time data for LY3819253, LY3832479, LY3853113, and [redacted] will be summarized descriptively. A population approach using a nonlinear mixed-effects modeling (NONMEM) program may also be performed.

Study data may be pooled with the results of other studies for population PK and PK/PD analysis purposes.

##### 9.4.4. Exploratory Analyses

Full details of the planned exploratory analyses will be described in the SAP.

Additional exploratory analyses of the data will be conducted as deemed appropriate. Study results may be pooled with the results of other studies for safety, PD, or population PK and PK/PD analysis purposes.

##### 9.4.5. Immunogenicity Analyses

If data from validated immunogenicity assays are available, treatment-emergent anti-drug antibodies (TE-ADAs) may be assessed.

Treatment-emergent ADAs are defined as participants

- with a 2-fold (1 dilution) increase in titer than the minimum required dilution if no ADAs were detected at baseline (treatment-induced ADA) or
- with a 4-fold (2 dilutions) increase in titer compared with baseline if ADAs were detected at baseline (treatment-boosted ADA).

The frequency and percentage of participants with preexisting ADA and who are TE-ADA positive (TE-ADA+) to LY3819253, LY3832479, LY3853113, and/or [redacted] will be tabulated.

The distribution of titers and frequency of neutralizing antibodies (if assessed) for the TE-ADA+ participants may also be tabulated.

The relationship between the presence of antibodies and PK parameters, efficacy response or safety to LY3819253, LY3832479, LY3853113 and/or [redacted] may also be assessed. Additional details may be provided in the SAP.

##### 9.4.6. Subgroup Analyses

This study is not powered for subgroup analyses; therefore, all subgroup analyses will be treated as exploratory.

Subgroup analyses will be conducted for the primary endpoint. Subgroups may include

- time of symptom onset to study randomization
- baseline severity of COVID-19
- age
- sex
- race
- ethnicity
- baseline weight
- baseline body mass index, or
- concomitant medication

Treatment group differences will be evaluated within each category of the subgroup regardless of whether the interaction is statistically significant. If any group within the subgroup is <10% of the total population, only summaries of the efficacy data will be provided, that is, no inferential testing.

Definitions for the levels of the subgroup variables, the analysis methodology, and any additional subgroup analyses will be defined in the SAP.

#### 9.5. Interim Analyses

The ongoing study may be modified based on planned interim analyses. Based on the observed data at the time of the interim analyses, the study may

- suspend enrollment to any treatment arm demonstrating lack of efficacy, and/or
- initiate/expand enrollment to an additional/existing treatment arm (or arms).

The modifications proposed are done so to ensure participants are being exposed to treatment with an acceptable risk-benefit profile during the ongoing trial. Additionally, the potential modifications will provide information to more fully characterize the dose response profile.

Monitoring of unblinded safety data (including AEs, SAEs, and selected laboratory measurements) will occur throughout the study and will be conducted by Assessment Committee (AC) members. Details of the unblinded safety reviews, including the frequency and approximate timing, are specified in the AC charter.

CCI [redacted]

The approximate timing, decision criteria, and statistical methods associated with each possible modification to the ongoing trial will be fully described in the SAP and AC Charter and finalized prior to the first study unblinding.

Periodic adjustments to the allocation ratio may be made to achieve the planned allocation across treatment arms at the conclusion of enrollment. If additional placebo participants are enrolled, then the allocation ratio may change accordingly.

Prior to the primary endpoint, only the AC (and individuals as defined in the unblinding plan) is authorized to evaluate unblinded interim analyses and safety analyses. Study sites will receive information about interim results only if they need to know for the safety of their participants.

Unblinding details are specified in the unblinding plan section of the SAP or in a separate unblinding plan document.

#### 9.6. Data Monitoring Committee (DMC)

The sponsor will form an AC to analyze the interim study data. To minimize any bias introduced into the analysis of the study results, analysis plans will be finalized and approved prior to the interim analyses.

The primary goal of the AC is to review the interim results regarding the continuing safety of study participants and the continuing validity and scientific merit of the study. Information that may unblind the study during the analyses will not be reported to study sites or the blinded study team until the study has been unblinded.

Overall committee structure information is in Section 10.1.5. Details of the AC will be provided in the AC charter.

### 10. Supporting Documentation and Operational Considerations

#### 10.1. Appendix 1: Regulatory, Ethical, and Study Oversight Considerations

##### 10.1.1. Regulatory and Ethical Considerations

This study will be conducted in accordance with the protocol and with the following:

- Consensus ethical principles derived from international guidelines including the Declaration of Helsinki and Council for International Organizations of Medical Sciences (CIOMS) International Ethical Guidelines
- Applicable International Council for Harmonization (ICH) Good Clinical Practice (GCP) Guidelines, and
- Applicable laws and regulations.

The protocol, protocol amendments, ICF, Investigator Brochure, and other relevant documents (e.g., advertisements) must be submitted to an IRB/IEC by the investigator and reviewed and approved by the IRB/IEC before the study is initiated.

Any amendments to the protocol will require IRB/IEC approval before implementation of changes made to the study design, except for changes necessary to eliminate an immediate hazard to study participants.

The investigator will be responsible for the following:

- Providing written summaries of the status of the study to the IRB/IEC annually or more frequently in accordance with the requirements, policies, and procedures established by the IRB/IEC
- Notifying the IRB/IEC of SAEs or other significant safety findings as required by IRB/IEC procedures, and
- Providing oversight of the conduct of the study at the site and adherence to requirements of 21 CFR, ICH guidelines, the IRB/IEC, European regulation 536/2014 for clinical studies (if applicable), and all other applicable local regulations.

Investigator sites are compensated for participation in the study as detailed in the clinical trial agreement.

##### 10.1.2. Financial Disclosure

Investigators and sub-investigators will provide the sponsor with sufficient, accurate financial information as requested to allow the sponsor to submit complete and accurate financial certification or disclosure statements to the appropriate regulatory authorities. Investigators are responsible for providing information on financial interests during the study and for 1 year after completion of the study.

##### 10.1.3. Informed Consent Process

The investigator or their representative will explain the nature of the study, including the risks and benefits, to the participant or their legally authorized representative and answer all questions regarding the study.

Participants must be informed that their participation is voluntary. Participants or their legally authorized representative will be required to sign a statement of informed consent and child/adolescent assent, as appropriate, that meets the requirements of 21 CFR 50, local regulations, ICH guidelines, Health Insurance Portability and Accountability Act (HIPAA) requirements, where applicable, and the IRB/IEC or study center.

Due to strict respiratory isolation policies, limited access to COVID-19 patient rooms, and SARS-CoV-2 transmissibility via droplet-contaminated paper, verbal consent and alternative methods of obtaining consent (for example, by phone) will be allowed if approved by the IRB.

If a signed paper copy of the ICF or child/adolescent assent is allowed by site/institution policy, then the process of how it will be obtained and stored will need to be determined.

Any variation from the standard consent process due to isolation and infection control should be sent to the IRB for approval prior to enrollment. The site will document the process in their regulatory files and demonstrate that the process has IRB concurrence or approval.

The medical record must include a statement that written informed consent or child/adolescent assent was obtained before the participant was entered in the study and the date the written consent or assent was obtained. The authorized person obtaining the informed consent or child/adolescent assent, and, if applicable, the individual designated to witness a verbal consent, must also sign the ICF.

Participants must be re-consented to the most current version of the ICF(s) during their participation in the study.

A copy of the ICF(s) must be provided to the participant or the participant's legally authorized representative and is kept on file.

##### 10.1.4. Data Protection

Participants will be assigned a unique identifier by the sponsor. Any participant records, datasets or tissue samples that are transferred to the sponsor will contain the identifier only; participant names or any information which would make the participant identifiable will not be transferred.

The participant must be informed that their personal study-related data will be used by the sponsor in accordance with local data protection law. The level of disclosure must also be explained to the participant who will be required to give consent for their data to be used as described in the informed consent.

The participant must be informed that their medical records may be examined by Clinical Quality Assurance auditors or other authorized personnel appointed by the sponsor, by appropriate IRB/IEC members, and by inspectors from regulatory authorities.

The sponsor has processes in place to ensure data protection, information security and data integrity. These processes include appropriate contingency plan(s) for appropriate and timely response in the event of a data security breach.

##### 10.1.5. Committees Structure

The AC will consist of members internal and external to the sponsor. The membership will include, at a minimum, a chair external to Lilly, a statistician and two physicians. The AC members will not have data entry/validation responsibilities or direct contact with the site(s) or testing facilities.

##### 10.1.6. Dissemination of Clinical Study Data

A clinical study report will be provided for this study and a summary of study information provided on publicly available websites.

##### 10.1.7. Data Quality Assurance

**Investigator responsibilities**

All participant data relating to the study will be recorded on printed or electronic CRF unless transmitted to the sponsor or designee electronically (e.g., laboratory data). The investigator is responsible for verifying that data entries are accurate and correct by physically or electronically signing the CRF.

The investigator must maintain accurate documentation (source data) that supports the information entered in the CRF.

The investigator must permit study-related monitoring, audits, IRB/IEC review, and regulatory agency inspections and provide direct access to source data documents.

**Data monitoring and management**

The Monitoring Plan includes

- monitoring details describing strategy, for example, risk-based initiatives in operations and quality such as Risk Management and Mitigation Strategies and Analytical Risk-Based Monitoring
- methods
- responsibilities and requirements
- handling of noncompliance issues, and
- monitoring techniques.

The sponsor or designee is responsible for the data management of this study including quality checking of the data. The sponsor assumes accountability for actions delegated to other individuals (e.g., Contract Research Organizations).

Study monitors will perform ongoing source data verification to confirm that data entered into the CRF by authorized site personnel are accurate, complete, and verifiable from source documents. They will also ensure that the safety and rights of participants are being protected, and that the study is being conducted in accordance with the currently approved protocol and any other study agreements, ICH GCP, and all applicable regulatory requirements.

If on-site monitoring activities cannot occur, alternative measures will be used. Examples of alternative measures are use of technology for off-site monitoring or providing pseudonymized copies of source documents to the monitor electronically. The remote source data verification will be focused on critical efficacy data and important safety data.

**Records retention and audits**

Records and documents, including signed ICFs, pertaining to the conduct of this study must be retained by the investigator for the time period outlined in the Clinical Trial Agreement (CTA) unless local regulations or institutional policies require a longer retention period.

No records may be destroyed during the retention period without the written approval of the sponsor. No records may be transferred to another location or party without written notification to the sponsor.

The sponsor or its representatives will periodically check a sample of the participant data recorded against source documents at the study site. The study may be audited by the sponsor or its representatives, and by regulatory agencies at any time. Investigators will be given notice before an audit occurs.

**Data Capture System**

The investigator is responsible for ensuring the accuracy, completeness, legibility, and timeliness of the data reported to the sponsor.

An electronic data capture system (EDC) will be used in this study for the collection of CRF data. The investigator maintains a separate source for the data entered by the investigator or designee into the sponsor-provided EDC system.

Only symptom assessments might be directly recorded by the investigator site personnel or a delegate into the EDC. The directly entered data will serve as source documentation. The investigator will not maintain an original, separate, written or electronic record of these data. A certified copy of the respective data entry will be downloaded by the investigator for retention.

The investigator is responsible for the identification of any data to be considered source and for the confirmation that data reported are accurate and complete by signing the CRF.

Data collected via the sponsor-provided data capture systems will be stored at third parties. The investigator will have continuous access to the data during the study and until decommissioning of the data capture systems. Prior to decommissioning, the investigator will receive an archival copy of pertinent data for retention.

Data managed by a central vendor, such as laboratory test data, will be stored electronically in the central vendor's database system and reports will be provided to the investigator for review and retention. Data will subsequently be transferred from the central vendor to the Sponsor data warehouse.

Data from complaint forms submitted to Sponsor will be encoded and stored in the global product complaint management system.

##### 10.1.8. Source Documents

Source documents provide evidence for the existence of the participant and substantiate the integrity of the data collected. Source documents are filed at the investigator's site.

Data reported on the CRF that are transcribed from source documents must be consistent with the source documents or the discrepancies must be explained. The investigator may need to request previous medical records or transfer records, depending on the study. Current medical records must be available.

The definition of what constitutes source data can be found in Section 10.1.7.

##### 10.1.9. Study and Site Start and Closure

The study start date is the date on which the clinical study will be open for recruitment of participants.

**Site Closure**

The sponsor designee reserves the right to close the study site or terminate the study at any time for any reason at the sole discretion of the sponsor. Study sites will be closed upon study completion. A study site is considered closed when all required documents and study supplies have been collected and a study-site closure visit has been performed.

The investigator may initiate study-site closure at any time, provided there is reasonable cause and enough notice is given in advance of the intended termination.

Reasons for the early closure of a study site by the sponsor or investigator may include but are not limited to:

- Failure of the investigator to comply with the protocol, the requirements of the IRB/IEC or local health authorities, the sponsor's procedures, or GCP guidelines
- Inadequate recruitment of participants by the investigator, or
- Discontinuation of further study intervention development.

If the study is prematurely terminated or suspended, the sponsor shall promptly inform the Investigators, the IECs/IRBs, the regulatory authorities, and any contract research organization(s) used in the study of the reason for termination or suspension, as specified by the applicable regulatory requirements. The Investigator shall promptly inform the participants and assure appropriate participant therapy and/or follow-up.

##### 10.1.10. Publication Policy

In accordance with the sponsor’s publication policy, the results of this study will be submitted for publication by a peer-reviewed journal if the results are deemed to be of significant medical importance.

##### 10.1.11. Investigator Information

Physicians with specialties, including, but not limited to infectious disease, acute or critical care, pulmonary disease, immunology, or other appropriate specialties may participate as investigators.

##### 10.1.12. Long-Term Sample Retention

Sample retention enables use of new technologies, response to regulatory questions, and investigation of variable response that may not be observed until later in the development of the intervention or after the intervention becomes commercially available.

This table describes the retention period for potential sample types.

| Sample Type | Custodian | Retention Period After Last Participant Visit |
|---|---|---|
| Pharmacodynamic Samples | Sponsor or designee | up to 7 years |
| Pharmacogenetics sample | Sponsor or designee | up to 7 years |
| Exploratory Biomarker Samples | Sponsor or designee | up to 7 years |
| Immunogenicity (ADA) sample | Sponsor or designee | up to 7 years |
| Pharmacokinetic (PK) sample | Sponsor or designee | up to 2 years |

#### 10.2. Appendix 2: Clinical Laboratory Tests

Clinical laboratory tests will be performed according to the SoA (Section 1.3).

Additional tests may be performed at any time during the study as determined necessary by the investigator or required by local regulations.

Central and local laboratories will be used. The table below describes when the local or central laboratory will be used.

Laboratory results that could unblind the study will not be reported to investigative sites or other blinded personnel.

Protocol-specific requirements for inclusion or exclusion of participants are detailed in Section 5 of the protocol.

Pregnancy testing will be performed according to the SoA.

Investigators must document their review of each laboratory safety report.

Refer to Section 10.6 for recommended laboratory testing for hypersensitivity events.

| Clinical Laboratory Tests | Comments |
|---|---|
| **Hematology** | Assayed by Lilly-designated laboratory. |
| Hemoglobin | |
| Hematocrit | |
| Erythrocyte count (RBCs - Red Blood Cells) | |
| Mean cell volume | |
| Mean cell hemoglobin | |
| Leukocytes (WBCs - White Blood Cells) | |
| Differential | |
| Neutrophils, segmented | |
| Lymphocytes | |
| Monocytes | |
| Eosinophils | |
| Basophils | |
| Platelets | |
| Cell Morphology (RBC and WBC) | |
| **Clinical Chemistry** | Assayed by Lilly-designated laboratory. |
| Sodium | |
| Potassium | |
| Chloride | |
| Bicarbonate | |
| Total bilirubin | |
| Direct bilirubin | |
| Alkaline phosphatase (ALP) | |
| Alanine aminotransferase (ALT) | |
| Aspartate aminotransferase (AST) | |
| Gamma-glutamyl transferase (GGT) | |
| Blood urea nitrogen (BUN) | |
| Creatinine | |
| Creatine kinase (CK) | |
| Uric acid | |
| Total protein | |
| Albumin | |
| Calcium | |
| Phosphorus | |
| Glucose | |
| Amylase | |
| Lipase | |
| Lactate dehydrogenase (LDH) | |
| **SARS-CoV-2 viral infection determination** | Local laboratory and/or Point-of-Care testing |
| **SARS-CoV-2 Test Panel** | Assayed by Lilly-designated laboratory. |

| Clinical Laboratory Tests | Comments |
|---|---|
| C-reactive protein (CRP); high-sensitivity | For adults only |
| Ferritin | For adults only |
| D-dimer | For adults only |
| Procalcitonin | For adults only |
| **Troponin** | For adults only |
| Troponin I | For adults only |
| Troponin T | For adults only |
| **Hormones (female)** | |
| Urine Pregnancy | Local laboratory |
| Serum Pregnancy | Local laboratory |
| **Pharmacokinetic Samples** | Assayed by Lilly-designated laboratory.<br>Results will not be provided to the investigative sites. |
| LY3819253<br>LY3832479<br>LY3853113<br>CCI [redacted] | |
| **Pharmacodynamic sample** | Assayed by Lilly-designated laboratory.<br>Results will not be provided to the investigative sites. |
| SARS-CoV-2 nasopharyngeal swab | |
| **Pharmacogenetics sample** | For adults only<br>Assayed by Lilly-designated laboratory.<br>Results will not be provided to the investigative sites. |
| **Exploratory Biomarker Samples** | Assayed by Lilly-designated laboratory.<br>Results will not be provided to the investigative sites. |
| Serum | |
| RNA (PAXGene) | |
| Whole Blood (EDTA) | For adults only |
| Whole Blood (EDTA) Epigenetics | For adults only |
| **Exploratory Biomarker Serum** | Assayed by Lilly-designated laboratory.<br>Results will not be provided to the investigative sites. |
| **Immunogenicity Samples** | Assayed by Lilly-designated laboratory.<br>Results will not be provided to the investigative sites. |
| Anti-LY3819253 antibodies<br>Anti-LY3832479 antibodies<br>Anti-LY3853113 antibodies<br>CCI [redacted] | |

#### 10.3. Appendix 3: Adverse Events: Definitions and Procedures for Recording, Evaluating, Follow-up, and Reporting

##### 10.3.1. Definition of AE

| **AE Definition** |
|---|
| • An AE is any untoward medical occurrence in a patient or clinical study participant, temporally associated with the use of study intervention, whether or not considered related to the study intervention.<br>• NOTE: An AE can therefore be any unfavorable and unintended sign (including an abnormal laboratory finding), symptom, or disease (new or exacerbated) temporally associated with the use of study intervention. |

| **Events <u>Meeting</u> the AE Definition** |
|---|
| • Any abnormal laboratory test results (hematology, clinical chemistry, or urinalysis) or other safety assessments (e.g., ECG, radiological scans, vital signs measurements), including those that worsen from baseline, considered clinically significant in the medical and scientific judgment of the investigator (i.e., not related to progression of underlying disease).<br>• Exacerbation of a chronic or intermittent pre-existing condition including either an increase in frequency and/or intensity of the condition.<br>• New conditions detected or diagnosed after study intervention administration even though it may have been present before the start of the study.<br>• Signs, symptoms, or the clinical sequelae of a suspected drug-drug interaction.<br>• Signs, symptoms, or the clinical sequelae of a suspected overdose of either study intervention or a concomitant medication. Overdose per se will not be reported as an AE/SAE unless it is an intentional overdose taken with possible suicidal/self-harming intent. Such overdose should be reported regardless of sequelae.<br>• "Lack of efficacy" or "failure of expected pharmacological action" per se will not be reported as an AE or SAE. Such instances will be captured in the efficacy assessments. However, the signs, symptoms, and/or clinical sequelae resulting from lack of efficacy will be reported as AE or SAE if they fulfill the definition of an AE or SAE. |

| **Events <u>NOT</u> Meeting the AE Definition** |
|---|
| • The following study-specific clinical events related to COVID-19 are exempt from adverse event reporting unless the investigator deems the event to be related to the administration of study drug:<br> o Hypoxemia due to COVID-19 requiring supplemental oxygen; |

| |
|---|
| o Hypoxemia due to COVID-19 requiring non-invasive ventilation or high flow oxygen devices;<br>o Respiratory failure due to COVID-19 requiring invasive mechanical ventilation or ECMO<br>• Any clinically significant abnormal laboratory findings or other abnormal safety assessments which are associated with the underlying disease, unless judged by the investigator to be more severe than expected for the participant's condition.<br>• The disease/disorder being studied or expected progression, signs, or symptoms of the disease/disorder being studied, unless more severe than expected for the participant's condition.<br>• Medical or surgical procedure (e.g., endoscopy, appendectomy): the condition that leads to the procedure is the AE.<br>• Situations in which an untoward medical occurrence did not occur (social and/or convenience admission to a hospital).<br>• Anticipated day-to-day fluctuations of pre-existing disease(s) or condition(s) present or detected at the start of the study that do not worsen. |

##### 10.3.2. Definition of SAE

If an event is not an AE per definition above, then it cannot be an SAE even if serious conditions are met (e.g., hospitalization for signs/symptoms of the disease under study, death due to progression of disease).

| **An SAE is defined as any untoward medical occurrence that, at any dose:** |
|---|
| **a. Results in death** |
| **b. Is life-threatening**<br>The term 'life-threatening' in the definition of 'serious' refers to an event in which the participant was at risk of death at the time of the event. It does not refer to an event, which hypothetically might have caused death, if it were more severe. |
| **c. Requires inpatient hospitalization or prolongation of existing hospitalization**<br>• In general, hospitalization signifies that the participant has been admitted to hospital for observation and/or treatment that would not have been appropriate in the physician's office or outpatient setting. Complications that occur during hospitalization are AEs. If a complication prolongs hospitalization or fulfills any other serious criteria, the event is serious. When in doubt as to whether "hospitalization" occurred or was necessary, the AE should be considered serious.<br>• Hospitalization for elective treatment of a pre-existing condition that did not worsen from baseline is not considered an AE. |

| |
|---|
| **d. Results in persistent disability/incapacity**<br>• The term disability means a substantial disruption of a person's ability to conduct normal life functions.<br>• This definition is not intended to include experiences of relatively minor medical significance such as uncomplicated headache, nausea, vomiting, diarrhea, influenza, and accidental trauma (e.g., sprained ankle) which may interfere with or prevent everyday life functions but do not constitute a substantial disruption. |
| **e. Is a congenital anomaly/birth defect** |
| **f. Other situations:**<br>• Medical or scientific judgment should be exercised in deciding whether SAE reporting is appropriate in other situations such as important medical events that may not be immediately life-threatening or result in death or hospitalization but may jeopardize the participant or may require medical or surgical intervention to prevent one of the other outcomes listed in the above definition. These events should usually be considered serious.<br>• Examples of such events include invasive or malignant cancers, intensive treatment in an emergency room or at home for allergic bronchospasm, blood dyscrasias or convulsions that do not result in hospitalization, or development of drug dependency or drug abuse. |

##### 10.3.3. Recording and Follow-Up of AE and/or SAE

| AE and SAE Recording |
|---|
| • When an AE/SAE occurs, it is the responsibility of the investigator to review all documentation (e.g., hospital progress notes, laboratory reports, and diagnostics reports) related to the event.<br>• The investigator will then record all relevant AE/SAE information in the CRF.<br>• It is **not** acceptable for the investigator to send photocopies of the participant's medical records to Sponsor or designee in lieu of completion of the AE/SAE CRF page.<br>• There may be instances when copies of medical records for certain cases are requested by Sponsor or designee. In this case, all participant identifiers, with the exception of the participant number, will be redacted on the copies of the medical records before submission to Sponsor or designee.<br>• The investigator will attempt to establish a diagnosis of the event based on signs, symptoms, and/or other clinical information. Whenever possible, the diagnosis (not the individual signs/symptoms) will be documented as the AE/SAE. |
| **Assessment of Intensity** |
| The investigator will make an assessment of intensity for each AE and SAE reported during the study and assign it to one of the following categories, which together with serious (i.e., |

SAE) criteria on the AE CRF ("results in death" and "life-threatening"), are aligned with the Division of AIDS (DAIDS) Table for Grading the Severity of Adult and Pediatric Adverse Events, version 2.1 (July 2017).

**Mild:** Mild symptoms causing no or minimal interference with usual social and functional activities, with intervention not indicated.

**Moderate:** Moderate symptoms causing greater than minimal interference with usual social and functional activities, with intervention indicated.

**Severe:** Severe symptoms causing inability to perform usual social and functional activities, with intervention or hospitalization indicated.

An event is defined as 'serious' when it meets at least 1 of the predefined outcomes as described in the definition of an SAE, NOT when it is rated as severe.

**Assessment of Causality**

- The investigator is obligated to assess the relationship between study intervention and each occurrence of each AE/SAE.
- A "reasonable possibility" of a relationship conveys that there are facts, evidence, and/or arguments to suggest a causal relationship, rather than a relationship cannot be ruled out.
- The investigator will use clinical judgment to determine the relationship.
- Alternative causes, such as underlying disease(s), concomitant therapy, and other risk factors, as well as the temporal relationship of the event to study intervention administration will be considered and investigated.
- The investigator will also consult the Investigator's Brochure (IB) and/or Product Information, for marketed products, in his/her assessment.
- For each AE/SAE, the investigator **must** document in the medical notes that he/she has reviewed the AE/SAE and has provided an assessment of causality.
- There may be situations in which an SAE has occurred and the investigator has minimal information to include in the initial report to Sponsor or designee. However, it is very important that the investigator always make an assessment of causality for every event before the initial transmission of the SAE data to Sponsor or designee.
- The investigator may change his/her opinion of causality in light of follow-up information and send a SAE follow-up report with the updated causality assessment.
- The causality assessment is one of the criteria used when determining regulatory reporting requirements.

**Follow-up of AEs and SAEs**

- The investigator is obligated to perform or arrange for the conduct of supplemental measurements and/or evaluations as medically indicated or as requested by Sponsor or

designee to elucidate the nature and/or causality of the AE or SAE as fully as possible. This may include additional laboratory tests or investigations, histopathological examinations, or consultation with other health care professionals.
- If a participant dies during participation in the study or during a recognized follow-up period, the investigator will provide Sponsor or designee with a copy of any post-mortem findings including histopathology.
- New or updated information will be recorded in the originally completed CRF.
- The investigator will submit any updated SAE data to Sponsor or designee within 24 hours of receipt of the information.

##### 10.3.4. Reporting of SAEs

| SAE Reporting via an Electronic Data Collection Tool |
| --- |
| • The primary mechanism for reporting an SAE will be the electronic data collection tool.<br>• If the electronic system is unavailable, then the site will use the paper SAE data collection tool (see next section) in order to report the event within 24 hours.<br>• The site will enter the SAE data into the electronic system as soon as it becomes available.<br>• After the study is completed at a given site, the electronic data collection tool will be taken off-line to prevent the entry of new data or changes to existing data.<br>• If a site receives a report of a new SAE from a study participant or receives updated data on a previously reported SAE after the electronic data collection tool has been taken off-line, then the site can report this information on a paper SAE form (see next section) or to the sponsor by telephone.<br>• Contacts for SAE reporting can be found in site training documents. |

| SAE Reporting via Paper CRF |
| --- |
| • Facsimile transmission of the SAE paper CRF is the preferred method to transmit this information to the sponsor.<br>• Initial notification via telephone does not replace the need for the investigator to complete and sign the SAE CRF pages within the designated reporting time frames.<br>• Contacts for SAE reporting can be found in site training documents. |

#### 10.4. Appendix 4: Contraceptive Guidance and Collection of Pregnancy Information

**Women**

Woman of Childbearing Potential (WOCBP)

A woman is considered fertile following menarche and until becoming post-menopausal unless permanently sterile (see below).

If fertility is unclear (e.g., amenorrhea in adolescents or athletes) and a menstrual cycle cannot be confirmed before first dose of study intervention, additional evaluation should be considered.

Woman not of Childbearing Potential

Women in the following categories are not considered WOCBP

1. Premenarchal
2. Premenopausal female with either
    a. Documented hysterectomy
    b. Documented bilateral salpingectomy, or
    c. Documented bilateral oophorectomy.

For individuals with permanent infertility due to an alternate medical cause other than the above, (e.g., mullerian agenesis, androgen insensitivity), investigator discretion should be applied to determining study entry.

Note: Determination can come from the site personnel's review of the participant's medical records, medical examination, or medical history interview.

3. Postmenopausal female is defined as, women with:
    a. 12 months of amenorrhea for women >55, with no need for FSH
    b. 12 months of amenorrhea for women >40 years old with FSH ≥40 mIU/mL and no other medical condition such as anorexia nervosa and not taking medications during the amenorrhea (e.g., oral contraceptives, hormones, gonadotropin releasing hormone, anti-estrogens, selective estrogen receptor modulators (SERMs), or chemotherapy that induced amenorrhea)

**Participation in the Study**

Women of child-bearing potential may participate in this study.

Women of child-bearing potential must test negative for pregnancy prior to initiation of treatment as indicated by a negative urine pregnancy test at the screening visit.

Women of child-bearing potential who are completely abstinent or in a same sex relationship, as part of their preferred and usual lifestyle, must agree to either remain abstinent or stay in a same sex relationship without sexual relationships with males.

All other women of child-bearing potential must agree to use two forms of effective contraception, where at least one form is highly effective (less than 1% failure rate), for the entirety of the study.

Abstinence or contraception must continue for 90 days after the last dose of intervention.

**Acceptable Methods of Contraception**

Highly effective methods of contraception (less than 1% failure rate) comprise, but are not limited to

- combination oral contraceptives
- implanted contraceptives, or
- intrauterine devices.

Effective methods of contraception comprise but are not limited to diaphragms with spermicide or cervical sponges.

**Not Acceptable Methods of Contraception**

Use of male and female condoms as a double barrier method is not considered acceptable due to the high failure rate when these barrier methods are combined.

Barrier protection methods without concomitant use of a spermicide are not an effective or acceptable method of contraception.

Periodic abstinence (e.g., calendar, ovulation, symptothermal, post-ovulation methods), declaration of abstinence just for the duration of a trial, and withdrawal are not acceptable methods of contraception.

**Men**

Men, regardless of their fertility status, must agree to either remain abstinent (if this is their preferred and usual lifestyle) or use condoms as well as one additional highly effective method of contraception (less than 1% failure rate) or effective method of contraception with non-pregnant women of child bearing potential partners for the duration of the study and until their plasma concentrations are below the level that could result in a relevant potential exposure to a possible fetus, predicted to be 90 days after the last dose.

Men with pregnant partners should use condoms during intercourse for the duration of the study and until the end of estimated relevant potential exposure to the fetus, predicted to be 90 days after the last dose.

**Acceptable Methods of Contraception**

Highly effective methods of contraception (less than 1% failure rate) comprise, but are not limited to

- combination oral contraceptives
- implanted contraceptives, or
- intrauterine devices.

Effective methods of contraception comprise but are not limited to diaphragms with spermicide or cervical sponges.

Men and their partners may choose to use a double–barrier method of contraception that must include use of a spermicide.

**Other Guidance**

Men should refrain from sperm donation for the duration of the study and until their plasma concentrations are below the level that could result in a relevant potential exposure to a possible fetus, predicted to be 90 days after the last dose.

**Collection of Pregnancy Information**

**Male participants with partners who become pregnant**

The investigator will attempt to collect pregnancy information on any male participant's female partner who becomes pregnant while the male participant is in this study. This applies only to male participants who receive study intervention.

After obtaining the necessary signed informed consent and assent (if applicable) from the pregnant female partner directly, the investigator will record pregnancy information on the appropriate form and submit it to the sponsor within 24 hours of learning of the partner's pregnancy.

The female partner will also be followed to determine the outcome of the pregnancy. Information on the status of the mother and child will be forwarded to the sponsor. Generally, the follow-up will be no longer than 6 to 8 weeks following the estimated delivery date. Any termination of the pregnancy will be reported including fetal status (presence or absence of anomalies) and indication for the procedure.

**Female Participants who become pregnant**

The investigator will collect pregnancy information on any female participant who becomes pregnant while participating in this study. The initial information will be recorded on the appropriate form and submitted to the sponsor within 24 hours of learning of a participant's pregnancy.

The participant will be followed to determine the outcome of the pregnancy. The investigator will collect follow-up information on the participant and the neonate and the information will be forwarded to the sponsor. Generally, this information will include a follow-up of at least 5 half-lives after last exposure or birth. Any termination of pregnancy will be reported, including fetal status (presence or absence of anomalies) or indication for the procedure.

While pregnancy itself is not considered to be an AE or SAE, any pregnancy complication or elective termination of a pregnancy for medical reasons will be reported as an AE or SAE.

A spontaneous abortion (occurring at <20 weeks gestational age) or still birth (occurring at >20 weeks gestational age) is always considered to be an SAE and will be reported as such.

Any post-study pregnancy related SAE considered reasonably related to the study intervention by the investigator will be reported to the sponsor as described in Section 8.3.4. While the investigator is not obligated to actively seek this information in former study participants, he or she may learn of an SAE through spontaneous reporting.

Any female participant who becomes pregnant while participating in the study will be withdrawn from the study and will follow the standard discontinuation process.

#### 10.5. Appendix 5: Genetics

Sample collection information is found in Appendix 2, Section 10.2 (Clinical Laboratory Tests).

**Use/Analysis of DNA**

Genetic variation may impact a participant's response to study intervention, susceptibility to, and severity and progression of disease. Variable response to study intervention may be due to genetic determinants that impact drug absorption, distribution, metabolism, and excretion; mechanism of action of the drug; disease etiology; and/or molecular subtype of the disease being treated. Therefore, where local regulations and IRB/IEC allow, a blood sample will be collected for DNA analysis from consenting participants.

DNA samples may be used for research related to SARS-CoV-2 and related diseases. They may also be used to develop tests/assays including diagnostic tests related to LY3819253, LY3832479, LY3853113, [redacted] or SARS-CoV-2. Genetic research may consist of the analysis of one or more candidate genes or the analysis of genetic markers throughout the genome as appropriate.

The samples may be analyzed as part of a multi-study assessment of genetic factors involved in the response to LY3819253, LY3832479, LY3853113, [redacted] or study interventions of this class to understand study disease or related conditions.

The results of genetic analyses may be reported in the clinical study report (CSR) or in a separate study summary.

The sponsor will store the DNA samples in a secure storage space with adequate measures to protect confidentiality.

The samples will be retained at a facility selected by the sponsor or its designee while research on SARS-CoV-2 continues but no longer than 7 years or other period as per local requirements.

#### 10.6. Appendix 6: Recommended Laboratory Testing for Hypersensitivity Events.

Laboratory assessments should be performed if the participant experiences generalized urticaria or if anaphylaxis is suspected.

- Collect sample after the participant has been stabilized, and within 1 to 2 hours of the event; however, samples may be obtained as late as 12 hours after the event as analytes can remain altered for an extended period of time. Record the time at which the sample was collected.
- Obtain a follow-up sample at the next regularly scheduled visit or after approximately 4 weeks, whichever is later.

#### Clinical Lab Tests for Hypersensitivity Events

| Hypersensitivity Tests | Notes<br>Selected test may be obtained in the event of anaphylaxis or systemic allergic/hypersensitivity reactions. |
|---|---|
| LY3819253, LY3832479, LY3853113, and CCI anti-drug antibodies (immunogenicity/ADA) | Assayed by Lilly-designated laboratory.<br>Results will not be provided to the investigative sites. |
| LY3819253, LY3832479, LY3853113, and CCI concentrations (PK) | Assayed by Lilly-designated laboratory.<br>Results will not be provided to the investigative sites. |
| Tryptase | Assayed by Lilly-designated laboratory.<br>Results will not be provided to the investigative sites.<br>Urine N-methylhistamine testing is performed in addition to tryptase testing. Collect the first void urine following the event. Collect a follow-up urine sample after approximately 4 weeks.<br>**Note:** If a tryptase sample is obtained more than 2 hours after the event (that is, within 2 to 12 hours), or is not obtained because more than 12 hours have lapsed since the event, collect a urine sample for N-methylhistamine testing. |
| N-methylhistamine | Assayed by Lilly-designated laboratory.<br>Results will not be provided to the investigative sites. |
| Drug-specific IgE | Will be performed if a validated assay is available.<br>Assayed by Lilly-designated laboratory.<br>Results will not be provided to the investigative sites. |
| Basophil activation test | Will be performed if a validated assay is available.<br>Assayed by Lilly-designated laboratory.<br>Results will not be provided to the investigative sites.<br>**NOTE:** The basophil activation test is an in vitro cell based assay that only requires a serum sample. It is a surrogate assay for drug specific IgE but is not specific for IgE. |
| Complement (C3, C3a and C5a) | Assayed by Lilly-designated laboratory.<br>Results will not be provided to the investigative sites. |
| Cytokine panel | Assayed by Lilly-designated laboratory.<br>Results will not be provided to the investigative sites. |

Abbreviations: ADA = anti-drug antibody; IgE = immunoglobulin E; PK = pharmacokinetic.

#### 10.7. Appendix 7: Liver Safety: Suggested Actions and Follow-up Assessments

**Close Hepatic Monitoring**

This table describes when close hepatic monitoring should occur.

| If a participant with baseline results of ... | develops the following elevations… |
|---|---|
| ALT or AST <1.5× ULN | ALT or AST ≥3× ULN |
| ALP <1.5× ULN | ALP ≥2× ULN |
| TBL <1.5× ULN | TBL ≥2× ULN (except for participants with Gilbert's syndrome) |
| ALT or AST ≥1.5× ULN | ALT or AST ≥2× baseline |
| ALP ≥1.5× ULN | ALP ≥2× baseline |
| TBL ≥1.5× ULN | TBL ≥2× baseline (except for participants with Gilbert's syndrome) |

The laboratory tests listed in Appendix 2, including ALT, AST, ALP, TBL, direct bilirubin, gamma-glutamyl transferase, and creatine kinase, should be repeated within 48 to 72 hours to confirm the abnormality and to determine whether it is increasing or decreasing.

If the abnormality persists or worsens, clinical and laboratory monitoring, and evaluation for the possible causes of the abnormal liver tests should be initiated by the investigator in consultation with the sponsor.

At a minimum, this evaluation should include a physical examination and a thorough medical history, including symptoms, recent illnesses (for example, heart failure, systemic infection, hypotension, seizures) recent travel, history of concomitant medications (including over-the-counter), herbal and dietary supplements, history of alcohol use, and other substance abuse.

Initially, monitoring of symptoms and hepatic biochemical tests should be done at a frequency of 1 to 3 times weekly, based on the participant's clinical condition and hepatic biochemical tests. Subsequently, the frequency of monitoring may be lowered to once every 1 to 2 weeks if the participant's clinical condition and laboratory results stabilize. Monitoring of ALT, AST, ALP, and TBL should continue until levels normalize or return to approximate baseline levels.

**Comprehensive Hepatic Evaluation**

A comprehensive evaluation should be performed to search for possible causes of liver injury if one or more of these conditions occur:

| If a participant with baseline results of... | develops the following elevations… |
|---|---|
| ALT or AST <1.5× ULN | ALT or AST ≥3× ULN with hepatic signs/symptoms*, or<br>ALT or AST ≥5× ULN |
| ALP <1.5× ULN | ALP ≥3× ULN |
| TBL <1.5× ULN | TBL ≥2× ULN (except for participants with Gilbert's syndrome) |
| ALT or AST ≥1.5× ULN | ALT or AST ≥2× baseline with hepatic signs/symptoms*, or<br>ALT or AST ≥3× baseline |
| ALP ≥1.5× ULN | ALP ≥2× baseline |
| TBL ≥1.5× ULN | TBL ≥1.5× baseline (except for participants with Gilbert's syndrome) |

*** Hepatic signs/symptoms are severe fatigue, nausea, vomiting, right upper quadrant abdominal pain, fever, rash, and/or eosinophilia >5%.**

At a minimum, this evaluation should include physical examination and a thorough medical history, as outlined above, as well as tests for PT-INR; tests for viral hepatitis A, B, C, or E; tests for autoimmune hepatitis; and an abdominal imaging study (for example, ultrasound or CT scan).

Based on the participant's history and initial results, further testing should be considered in consultation with the Lilly-designated medical monitor, including tests for hepatitis D virus (HDV), cytomegalovirus (CMV), Epstein-Barr virus (EBV), acetaminophen levels, acetaminophen protein adducts, urine toxicology screen, Wilson's disease, blood alcohol levels, urinary ethyl glucuronide, and serum phosphatidylethanol.

Based on the circumstances and the investigator's assessment of the participant's clinical condition, the investigator should consider referring the participant for a hepatologist or gastroenterologist consultation, magnetic resonance cholangiopancreatography (MRCP), endoscopic retrograde cholangiopancreatography (ERCP), cardiac echocardiogram, or a liver biopsy.

#### Additional Hepatic Data Collection (Hepatic Safety CRF)

Additional hepatic safety data collection in hepatic safety case report forms (CRF) should be performed in study participants who either have a hepatic event considered to be an SAE or meet 1 or more of these conditions:

| If a participant with baseline... | has the following elevations… |
|---|---|
| ALT <1.5 ×ULN | ALT ≥5 × ULN on 2 or more consecutive blood tests |
| ALP <1.5 × ULN | ALP ≥2 × ULN on 2 or more consecutive blood tests |
| TBL <1.5 × ULN | TBL ≥2 × ULN, except for cases of known Gilbert's syndrome |
| ALT ≥1.5 × ULN | ALT ≥3 × baseline on 2 or more consecutive blood tests |
| ALP ≥1.5 × ULN | ALP ≥2 × baseline on 2 or more consecutive blood tests |
| TBL ≥1.5 × ULN | TBL ≥2 × baseline |

Abbreviations: ALP = alkaline phosphatase; ALT = alanine aminotransferase; AST = aspartate aminotransferase; TBL = total bilirubin; ULN = upper limit of normal.

Note: The interval between the 2 consecutive blood tests should be at least 2 days.

##### Hepatic Evaluation Testing

The Lilly-designated central laboratory must complete the analysis of all selected testing except for microbiology testing.

Local testing may be performed in addition to central testing when necessary for immediate participant management.

Results will be reported if a validated test or calculation is available.

| **Hematology** | **Clinical Chemistry** |
|---|---|
| Hemoglobin | Total bilirubin |
| Hematocrit | Direct bilirubin |
| Erythrocytes (RBCs - red blood cells) | Alkaline phosphatase (ALP) |
| Leukocytes (WBCs - white blood cells) | Alanine aminotransferase (ALT) |
| Differential: | Aspartate aminotransferase (AST) |
| Neutrophils, segmented | Gamma-glutamyl transferase (GGT) |
| Lymphocytes | Creatine kinase (CK) |
| Monocytes | **Other Chemistry** |
| Basophils | Acetaminophen |
| Eosinophils | Acetaminophen protein adducts |
| Platelets | Alkaline phosphatase isoenzymes |
| Cell morphology (RBC and WBC) | Ceruloplasmin |
| **Coagulation** | Copper |
| | Ethyl alcohol (EtOH) |
| Prothrombin time, INR (PT-INR) | Haptoglobin |

| | |
|---|---|
| **Serology** | Immunoglobulin IgA (quantitative) |
| Hepatitis A virus (HAV) testing: | Immunoglobulin IgG (quantitative) |
| HAV total antibody | Immunoglobulin IgM (quantitative) |
| HAV IgM antibody | Phosphatidylethanol (PEth) |
| Hepatis B virus (HBV) testing: | **Urine Chemistry** |
| Hepatitis B surface antigen (HBsAg) | Drug screen |
| Hepatitis B surface antibody (anti-HBs) | Ethyl glucuronide (EtG) |
| Hepatitis B core total antibody (anti-HBc) | **Other Serology** |
| Hepatitis B core IgM antibody | Anti-nuclear antibody (ANA) |
| Hepatitis B core IgG antibody | Anti-smooth muscle antibody (ASMA) [a] |
| HBV DNA [b] | Anti-actin antibody [c] |
| Hepatis C virus (HCV) testing: | Epstein-Barr virus (EBV) testing: |
| HCV antibody | EBV antibody |
| HCV RNA [b] | EBV DNA [b] |
| Hepatitis D virus (HDV) testing: | Cytomegalovirus (CMV) testing: |
| HDV antibody | CMV antibody |
| Hepatitis E virus (HEV) testing: | CMV DNA [b] |
| HEV IgG antibody | Herpes simplex virus (HSV) testing: |
| HEV IgM antibody | HSV (Type 1 and 2) antibody |
| HEV RNA [b] | HSV (Type 1 and 2) DNA [b] |
| **Microbiology** [d] | Liver kidney microsomal type 1 (LKM-1) antibody |
| Culture: | |
| Blood | |
| Urine | |

[a] Not required if anti-actin antibody is tested.

[b] Reflex/confirmation dependent on regulatory requirements, testing availability, or both.

[c] Not required if anti-smooth muscle antibody (ASMA) is tested.

[d] Assayed ONLY by investigator-designated local laboratory; no central testing available.

#### 10.8. Appendix 8: Abbreviations

| Term | Definition |
|---|---|
| **AC** | assessment committee |
| **ADA** | anti-drug antibody |
| **ADE** | antibody-dependent enhancement |
| **assent** | Agreement from a child or other individual who is not legally capable of providing consent, but who can understand the circumstances and potential risks involved in participating in a study. |
| **blinding/masking** | A single-blind study is one in which the investigator and/or his staff are aware of the treatment but the participant is not, or vice versa, or when the sponsor is aware of the treatment but the investigator and/his staff and the participant are not.<br>A double-blind study is one in which neither the participant nor any of the investigator or sponsor staff who are involved in the treatment or clinical evaluation of the participants are aware of the treatment received. |
| **CIOMS** | Council for International Organizations of Medical Sciences |
| **Complaint** | A complaint is any written, electronic, or oral communication that alleges deficiencies related to the identity, quality, purity, durability, reliability, safety or effectiveness, or performance of an intervention. |
| **Compliance** | Adherence to all study-related, good clinical practice (GCP), and applicable regulatory requirements. |
| **CRP** | clinical research physician: Individual responsible for the medical conduct of the study. Responsibilities of the CRP may be performed by a physician, clinical research scientist, global safety physician or other medical officer. |
| **DMC** | data monitoring committee |
| **ECG** | Electrocardiogram |
| **FiO2** | fraction of inspired oxygen |
| **Enroll** | The act of assigning a participant to a treatment. Participants who are enrolled in the study are those who have been assigned to a treatment. |
| **Enter** | Participants entered into a study are those who sign the informed consent form directly or through their legally acceptable representatives. |
| **GCP** | good clinical practice |
| **IB** | Investigator's Brochure |
| **ICF** | informed consent form |
| **ICH** | International Council for Harmonisation |

| | |
|---|---|
| **IgG1** | Immunoglobulin G1 |
| **IMP** | Investigational Medicinal Product |
| **Informed consent** | A process by which a participant voluntarily confirms his or her willingness to participate in a particular study, after having been informed of all aspects of the study that are relevant to the participant's decision to participate. Informed consent is documented by means of a written, signed and dated informed consent form. |
| **interim analysis** | An interim analysis is an analysis of clinical study data, separated into treatment groups, that is conducted before the final reporting database is created/locked. |
| **Intervention** | A pharmaceutical form of an active ingredient or placebo being tested or used as a reference in a clinical trial, including products already on the market when used or assembled (formulated or packaged) in a way different from the authorized form, or marketed products used for an unauthorized indication, or marketed products used to gain further information about the authorized form. |
| **IWRS** | interactive web-response system |
| **mAb** | monoclonal antibody |
| **NP** | Nasopharyngeal |
| **Participant** | Equivalent to CDISC term "subject": an individual who participates in a clinical trial, either as recipient of an investigational medicinal product or as a control |
| **PK/PD** | pharmacokinetics/pharmacodynamics |
| **SAE** | serious adverse event |
| **SAP** | statistical analysis plan |
| **Screen** | The act of determining if an individual meets minimum requirements to become part of a pool of potential candidates for participation in a clinical study. |
| **SpO2** | saturation of peripheral oxygen |

#### 10.9. Appendix 9: Protocol Amendment History

**Amendment f 16 March 2021**

**Overall Rationale for the Amendment:**

The purpose of this amendment is to incorporate a new study intervention, LY3853113, into the study design.

| Section # and Name | Description of Change | Brief Rationale |
|---|---|---|
| 1.1 Synopsis; 1.2 Schema; 1.3 Schedule of Activities; 3 Objectives and Endpoints; 4.1 Overall Design; 4.2 Scientific Rationale for Study Design; 6.1 Study Intervention(s) Administered; 8.1 Efficacy Assessments; 8.4 Treatment of Overdose; 8.5 Pharmacokinetics; 8.5.1 Bioanalytical; 8.6 Pharmacodynamics; 8.8 Biomarkers; 8.9 Immunogenicity Assessments; 10.2 Appendix 2: Clinical Laboratory Tests; 10.5 Appendix 5: Genetics; 10.6 Appendix 6: Recommended Laboratory Testing for Hypersensitivity Events | Added Arms 9-13 to study design; updated language throughout protocol to include LY3853113. | Arms added to incorporate new study intervention (LY3853113) into design; language updated throughout to align with the addition of LY3853113 |
| 1.1 Synopsis; 2 Introduction; 2.1 Study Rationale; 2.2 Background; 2.3 Benefit/Risk Assessment | Provided background information for LY3853113 | Updated section to include information about LY3853113 |
| 2.2 Background | Updated LY3819253+ LY3832479 safety information | Updated to provide more recent safety information |
| 4.3 Justification for Dose | Added justification for LY3853113 + LY3819253 + LY3832479 combination | Updated to provide justification for LY3853113 |
| 5.1 Inclusion Criteria; 5.2 Exclusion Criteria; 10.1.3 Informed Consent Process | Added specifications for low or high-risk participants; expanded participant population to include adolescents | Alignment with updated study design |
| 5.1 Inclusion Criteria | Criterion 3: Updated list of COVID-19 symptoms | Updated list to align with current guidance |
| 5.1 Inclusion Criteria; Appendix 4: Contraceptive Guidance and Collection of Pregnancy Information | Criterion 5: Clarified provision of reproductive and contraceptive agreements and guidance in Appendix 4 | Clarification |
| 5.2 Exclusion Criteria | Added Criterion 29 | Alignment with updated study design |
| 6.3 Measures to Minimize Bias: Randomization and Blinding | Added stratification factors for arms 12-13; specified blinding for arms 1-11. | Alignment with updated study design |
| 6.5 Concomitant Therapy | Added statement clarifying that prior SARS-CoV-2 vaccines should be recorded | Clarification |

| Section # and Name | Description of Change | Brief Rationale |
|---|---|---|
| 8.2.2 Vital Signs | Added new tables for Arms 9-11, 13 | Alignment with updated study design |
| 9.1 Statistical Hypotheses; 9.2 Sample Size Determination; 9.4 Statistical Analyses; 9.4.2 Primary Endpoints; 9.4.3.1 Safety (arms 1-11); 9.4.3.2 Additional Secondary Endpoints; 9.4.3.3 Pharmacokinetic Analyses; 9.4.5 Immunogenicity Analyses | Updated statistical considerations to incorporate LY3853113 into study design | Alignment with updated study design |
| Throughout | Minor editorial and formatting changes | Minor, therefore not detailed |

**Amendment e 25 January 2021**

**Overall Rationale for the Amendment:**

CCI [redacted] Additionally, the amendment incorporates new background information for CCI [redacted] and updates the benefit/risk language of LY3819253.

| Section # and Name | Description of Change | Brief Rationale |
|---|---|---|
| CCI [redacted] | [redacted] | [redacted] |
| 2.2. Background | [redacted] | CCI [redacted] |
| 2.3. Benefit/Risk Assessment | Revised language regarding clinical worsening of COVID-19 symptoms after LY3819253 administration | Alignment with bamlanivimab EUA Fact Sheet |
| Throughout | Minor editorial and formatting changes | Minor, therefore not detailed |

**Amendment d 15 January 2021**

**Overall Rationale for the Amendment:**

The purpose of this amendment is to remove pregnant women from the study population.

| Section # and Name | Description of Change | Brief Rationale |
|---|---|---|
| 2.3 Benefit/Risk Assessment; 4.2 Scientific Rationale for Study Design; 5.1 Inclusion Criteria; 5.2 Exclusion Criteria; 7.2 Participant Discontinuation/Withdrawal from the Study; 8.2.3 Clinical Laboratory Assessments; Appendix 4: Contraceptive Guidance and Collection of Pregnancy Information; 11 References | Removal of pregnant women from study population | Response to IRB feedback |

**Amendment c 06 January 2021**

**Overall Rationale for the Amendment:**

The purpose of this amendment is to incorporate a new study intervention, CCI [redacted] into the study design.

| **Section # and Name** | **Description of Change** | **Brief Rationale** |
|---|---|---|
| 1.1 Synopsis; 1.2 Schema; 1.3 Schedule of Activities; 3 Objectives and Endpoints; 4.1 Overall Design; 4.2 Scientific Rationale for Study Design; 6.1 Study Intervention(s) Administered; 8.1 Efficacy Assessments; 8.4 Treatment of Overdose; 8.5 Pharmacokinetics; 8.5.1 Bioanalytical; 8.6 Pharmacodynamics; 8.8 Biomarkers; 8.9 Immunogenicity Assessments; 10.2 Appendix 2: Clinical Laboratory Tests; 10.6 Appendix 6: Recommended Laboratory Testing for Hypersensitivity Events | CCI [redacted] | [redacted] |
| 1.3 Synopsis; 3 Objectives and Endpoints; 9.4.3.2 Additional Secondary Endpoints | Removed Day 22 from proportion of participants that achieve SARS-CoV-2 clearance | No NP swab collection on Day 22 |
| 1.1 Synopsis; 2 Introduction; 2.1 Study Rationale; 2.2 Background; 2.3 Benefit/Risk Assessment | CCI [redacted] | [redacted] |
| 4.3 Justification for Dose | CCI [redacted] | [redacted] |
| 5.1 Inclusion criteria; 5.2 Exclusion criteria; 8.2.3 Clinical Laboratory Assessments; 8.3.5 Pregnancy; 10.4 Appendix 4: Contraceptive Guidance and Collection of Pregnancy Information | Updated study population to include pregnant women | Including due to potential favorable benefit/risk assessment for high-risk group |
| 9.1 Statistical Hypotheses; 9.2 Sample Size Determination; 9.4.3.2 Additional Secondary Endpoints; 9.5 Interim Analyses | CCI [redacted] | [redacted] |
| Throughout | Minor editorial and formatting changes | Minor, therefore not detailed |

**Amendment b 03 December 2020**

**Overall Rationale for the Amendment:**

The purpose of this amendment is to incorporate a new treatment arm [redacted] into the study design. Additionally, minor administrative changes and clarifications have been made to the Schedule of Activities and Secondary Objectives and Endpoints.

| **Section # and Name** | **Description of Change** | **Brief Rationale** |
|---|---|---|
| 1.1 Synopsis; 1.2 Schema; 4.1.1 Design Outline; 6.1 Study Intervention(s) Administered; 9.2 Sample Size Determination | Added treatment [redacted] to study; specified approximate number of participants to be enrolled into [redacted] | Added to explore new dose level in study. |
| 1.1 Synopsis; 3 Objectives and Endpoints; 6.1 Study Intervention(s) Administered; 9.4.3.2 Additional Secondary Endpoints | Modified secondary endpoints for symptom resolution, symptom improvement, SARS-CoV-2 viral load, and SARS-CoV-2 clearance | Modified for consistency with other secondary endpoints |
| 1.3 Schedule of Activities | Modified Day 11 visit window | Administrative |
| 6.1 Study Intervention(s) Administered | Changed ‘pharmacy manual’ to ‘pharmacy preparation instructions’ | Administrative |
| 6.3 Measures to Minimize Bias: Randomization and Blinding | Added language clarifying staggered enrollment of participants | Clarification |
| 9.1 Statistical Hypotheses | Updated hypothesis to include [redacted] | Modifications made to align with updated study design |
| 9.5 Interim Analyses | Provided details of additional interim analysis | Added to align with updated study design |
| Throughout | Minor editorial changes | Minor, therefore not detailed |

#### Amendment a: 16 October 2020

##### Overall Rationale for the Amendment:

This amendment modifies the secondary endpoints that measure clinical events (deaths, hospitalizations, and ER visits) to include events through Day 29, in response to FDA feedback.

| Section # and Name | Description of Change | Brief Rationale |
|---|---|---|
| Section 1.1 Synopsis<br>Section 3 Objectives and Endpoints<br>Section 9.4.3.2 Additional Secondary Endpoints | Added Day 29 to clinical events secondary endpoints | FDA Feedback |
| Throughout the protocol | Minor editorial changes | Minor, therefore not described |

Leo Document ID = 75f8c245-a194-48ae-8def-5381f1ee605b

Approver: PPD

Approval Date & Time: 17-May-2021 19:49:29 GMT

Signature meaning: Approved

Approver: PPD

Approval Date & Time: 18-May-2021 00:09:34 GMT

Signature meaning: Approved
