## Supplementary material for "Bebtelovimab, alone or together with bamlanivimab and etesevimab, as a broadly neutralizing monoclonal antibody treatment for mild to moderate, ambulatory COVID-19": Redacted PYAH Protocol Addenda (4)

### Title Page

**Confidential Information**

The information contained in this document is confidential and is intended for the use of clinical investigators. It is the property of Eli Lilly and Company or its subsidiaries and should not be copied by or distributed to persons not involved in the clinical investigation of LY3819253, LY3832479, and LY3853113, unless such persons are bound by a confidentiality agreement with Eli Lilly and Company or its subsidiaries.

**Note to Regulatory Authorities:** This document may contain protected personal data and/or commercially confidential information exempt from public disclosure. Eli Lilly and Company requests consultation regarding release/redaction prior to any public release. In the United States, this document is subject to Freedom of Information Act (FOIA) Exemption 4 and may not be reproduced or otherwise disseminated without the written approval of Eli Lilly and Company or its subsidiaries.

**Protocol Title:**

A Randomized, Double-blind, Placebo-Controlled, Phase 2 Study to Evaluate the Efficacy and Safety of Mono and Combination Therapy with Monoclonal Antibodies in Participants with Mild to Moderate COVID-19 Illness (BLAZE-4)

**Protocol Number:** J2X-MC-PYAH

**Addendum Number:** 4

**Addendum Statement:** This addendum is to be performed in addition to all procedures required by protocol J2X-MC-PYAH or any subsequent amendments to that protocol.

**Compound:** LY3819253, LY3832479, LY3853113

**Sponsor Name:** Eli Lilly and Company
**Legal Registered Address:** Indianapolis, Indiana USA 46285

**Regulatory Agency Identifier Number(s)**
IND: 150440

**Approval Date:** Protocol Addendum (4) Electronically Signed and Approved by Lilly on date provided below.

Approval Date: 16-Mar-2021 GMT

### 1. Rationale for Addendum

Eli Lilly and Company has a partnership with AbCellera Biologics Inc. (AbCellera; Vancouver, Canada) to develop neutralizing IgG1 monoclonal antibodies (mAbs) to the Spike (S) protein of SARS-CoV-2 as a potential treatment for COVID-19. Candidate antibody gene sequences have been selected from a recently recovered COVID-19 United States patient's serum using AbCellera's core platform screening technologies.

Recently, emergence of mutations to the spike protein of SARS-CoV-2 threaten to render current treatments and vaccines not as effective.

LY3853113 is a novel, highly potent IgG1 neutralizing mAb targeting the spike protein of SARS-CoV-2 that was created in partnership with AbCellera. It binds an epitope within the receptor binding domain that is distinct from those bound by LY3819253 and LY3832479. LY3853113 can neutralize the Wuhan reference strain as well as individual residues present in recent variants of concern (i.e. L452R, D614G, N501Y, N439K, K417N, and E484K). Critically, pseudovirus assays demonstrate that LY3853113 can neutralize variants with the specific combination of receptor binding domain residues of the B.1.1.351 (South African) and B.1.1.28 (P.1/Brazil origin) strains. These variants were recently reported in the United States (CDC 2021), thus it is imperative that new treatments for these emerging variants are developed and deployed quickly.

The purpose of this Phase 1, double-blind, randomized, placebo-controlled, ascending dose substudy is to characterize the safety and tolerability of LY3853113 alone by intravenous infusion CCI [redacted], and in combination with LY3819253 and LY3832479 by intravenous infusion.

### 2. Protocol Additions

#### 2.1. Addendum Schema

Participants enrolled into this addendum will follow the schema below in place of the schema in the main protocol.

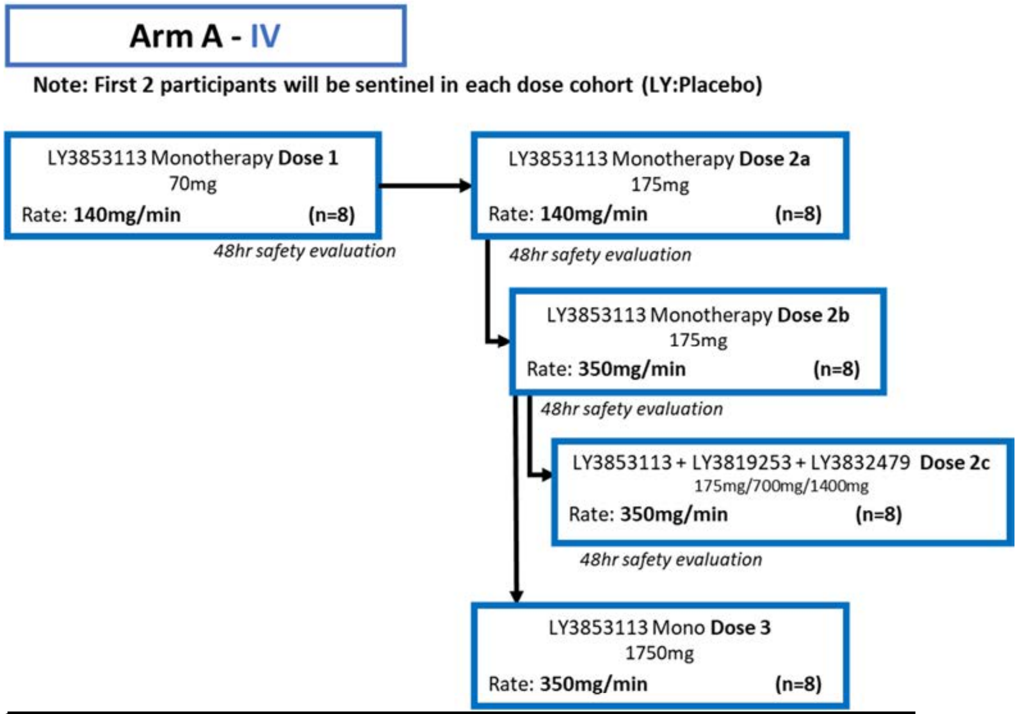

[redacted]

**Note:** Safety data from participants administered LY3819253 and LY3832479, expected prior to the start of the addendum, are informing on a maximum rate not exceeding 350 mg/min. The rate from Doses 2b, 2c, and Dose 3 will be defined in the pharmacy preparation instructions.

The first 2 participants in each cohort will be sentinel (LY3853113:placebo). Subsequent participants will be randomized to the remaining treatment allocations, 5 to LY3853113 or LY3853113 in combination with LY3819253 and LY3832479, and 1 to placebo.

Additional participants may be enrolled in case of discontinuation. In case of discontinuation, a newly enrolled participant will have the same treatment assignment as the corresponding discontinued participant.

Abbreviations: IV = intravenous; n = number of participants; [redacted].

#### 2.2. Schedule of Activities

Participants enrolled into this addendum will follow the Schedule of Activities below in place of the main protocol schedule(s).

| **J2X-MC-PYAH Addendum 4** | **Screen** | **Double-blind treatment and assessments** | | | | | | | | | | ED | Follow-up if hospital inpatient on Day 29 | **Post-treatment follow-up** | | **Comments** |
|---|---|---|---|---|---|---|---|---|---|---|---|---|---|---|---|---|
| **Study Day** | | **1** | **2*** | **3** | **4*** | **5** | **6*** | **7** | **11** | **22*** | **29** | ED | Every 7 days until discharge or Day 60 | **60** | **85** | Screening and Day 1 procedures may occur on the same day.<br>* = telephone visit(s) for all treatment arms |
| **Visit window (number of days)** | | -- | -- | +1 | -- | +1 | -- | +1 | -2/+3 | ±3 | ±3 | ±2 | ±2 | ±4 | ±4 | Visits may not be combined. |
| Urine or serum pregnancy | X | | | | | | | | | | | | | X | X | Only for WOCBP as defined in the main protocol, Section 10.4 Appendix 4.<br>Local laboratory |
| Pharmacokinetic (PK) sample | | X | | X | | X | | X | X | | X | X | | X | X | Day 1:<br>No pre-dose sample needed.<br>IV - one post-dose sample within 30 minutes after the end of IV infusion (may include the flush).<br>CCI [redacted]<br>All other scheduled samples may occur at any time during the day.<br>No samples needed if participant is hospitalized.<br>Lilly-designated central laboratory |
| Immunogenicity (ADA) sample | | X | | | | | | | X | | X | X | | X | X | Day 1: collect before treatment administration.<br>All other scheduled and unscheduled immunogenicity samples should be collected with a time-matched PK sample.<br>No samples needed if participant is hospitalized<br>Lilly-designated central laboratory |
| Pharmacodynamic (PD) NP swab | | X | | X | | X | | X | X | | X | X | | | | Swab is taken from both nostrils.<br>Day 1: swab before treatment administration.<br>No samples needed if participant is hospitalized<br>Lilly-designated central laboratory |

#### 2.3. Benefit/Risk Assessment

The administration of therapeutic mAbs has the potential for injection site reactions and hypersensitivity, including anaphylaxis and infusion related reactions. Increased rates of administration may be associated with an increased risk of such reactions.

The infusions in this study will be administered at a controlled rate, the study participants will be monitored closely, and adjustments in the infusion rate will be made or the infusion stopped, if indicated. Additional information regarding infusion reaction management options is in Section 6.1.1 of the main protocol.

More detailed information about the known and expected benefits and risks and reasonably expected adverse events of LY3819253, LY3832479, and LY3853113 may be found in each respective IB.

#### 2.4. Objectives and Endpoints

| Objectives | Endpoints |
|---|---|
| **Primary** | |
| Characterize the safety and tolerability of LY3853113 alone [redacted] and in combination with LY3819253 and LY3832479 [redacted] | • Safety assessments such as AEs and SAEs |
| **Secondary** | |
| Characterize the pharmacokinetics of LY3853113 [redacted] | • LY3853113 mean concentration on Day 29 |
| Characterize the pharmacodynamics of LY3853113 alone [redacted] and in combination with LY3819253 and LY3832479 [redacted] | • Change from baseline in SARS-CoV-2 viral load (Days 3, 5, 7, and 11)<br>• Proportion of participants with SARS-CoV-2 viral load greater than 5.27 on Day 7<br>• SARS-CoV-2 viral load AUC assessed through Day 11<br>• Time to SARS-CoV-2 clearance |
| **Tertiary/Exploratory** | |
| Characterize the participant’s clinical status | • Duration (days) of hospitalization<br>• Proportion (percentage) of participants admitted to ICU<br>• Proportion (percentage) of participants requiring mechanical ventilation |
| Characterize the pharmacodynamics of LY3853113 alone [redacted] and in combination with | • Proportion of participants that achieve SARS-CoV-2 clearance (Days 3, 5, 7, 11, 29)<br>• 75th percentile of SARS-CoV-2 viral load at Day 7 |

| | |
|---|---|
| LY3819253 and LY3832479 CCI [redacted] | |
| Characterize emergence of viral resistance to LY3853113 | • Comparison from baseline to the last evaluable time point up to Day 29 |

Abbreviations: AE = adverse event; ICU = intensive care unit; IV = intravenous; SAE = serious adverse event; SARS-CoV-2 = severe acute respiratory syndrome coronavirus 2; CCI [redacted].

#### 2.5. Study Design

This is a Phase 1, double-blind, randomized, placebo-controlled, ascending dose substudy that may progress to a Phase 2, double-blind, randomized, placebo-controlled phase.

**Screening**

Interested participants will sign the appropriate informed consent document(s) prior to completion of any procedures. Screening may be performed up to 48 hours prior to dosing. Screening and Day 1 may occur on the same day.

**Treatment and Assessment Period**

This is the general sequence of events during the treatment and assessment period:

- Participants are randomized to intervention group
- Baseline procedures and sample collection are completed
- Participants receive study intervention, and
- All safety monitoring and post-administration sample and data collection are performed.

Remote follow-up visits may be conducted to remove the burden of return visits to the clinic and clinical trial staff reflecting limited medical resources in the COVID-19 pandemic.

This addendum will comprise up to 5 dose cohorts to receive study intervention by IV infusion CCI [redacted]

Cohorts will comprise at least 8 participants each:

- 6 randomized to LY3853113 alone or in combination with LY3819253 and LY3832479, and
- 2 randomized to placebo.

Sentinel dosing will be used in each dose cohort that represents a dose increase or infusion rate change (increase in mg/minute) from the preceding cohort. The first 2 participants in each cohort will be randomized 1:1 to LY3853113 and placebo.

Safety and tolerability will be reviewed for sentinel participants up to 24 hours after dosing. The investigator and the Lilly sponsor team are responsible for determining if safety and tolerability is acceptable to continue with dosing subsequent participants.

Subsequent participants will be randomized to the remaining treatment allocations, 5 to LY3853113 and 1 to placebo.

The decision to dose the next cohort will be made when all participants from the previous cohort have been dosed and safety data is assessed for at least 48 hours after the IV infusion CCI [redacted] by the investigator(s) and Lilly sponsor team.

#### 2.6. Justification of Dose

**Intravenous dosing for LY3853113**

The 175mg dose is the intended target dose for the Phase 2 portion of the study.

The target therapeutic dose of 175 mg was selected using PK/PD modeling in a manner similar to the approaches that were used for LY3819253 and LY3832479. The PK/PD modeling approach includes in vitro potency data (i.e. IC90), predicted human PK, and the expected response in terms of maximal reduction in viral load. The 175 mg dose is expected to result in at least 90% of the population achieving drug concentrations above IC90 through at least 28 days after drug administration, and results in maximum reduction in viral load based on a PK-viral dynamic model.

This Phase 1 addendum will evaluate 3 IV dose levels.

The first dose level is approximately 3 times lower than the target dose level (70 mg). The third dose level is approximately 10 times higher than the target dose (1750 mg) to further evaluate the safety of LY3853113. Based on current knowledge of similar neutralizing mAbs (LY3819253 and LY3832479) that have been studied at doses of up to 7000 mg, the third dose level is anticipated to have an acceptable safety profile. Based on preliminary non-clinical PK results, the PK profile of LY3853113 is similar to that of LY3819253.

Further details about dose selection and coverage of variants of interest may be found in the IB. These doses may be amended if necessary, based on emerging data.

**Intravenous dosing for LY3819253 and LY3832479**

The IV doses for LY3819253 and LY3832479 will be 700 mg and 1400 mg, respectively. These doses are currently the authorized doses in the EUA.

CCI [redacted]

[redacted]

#### 2.7. Study Population

##### 2.7.1. Inclusion Criteria

Participants are eligible to be included in the study only if all of the following criteria apply:

**Age**

1. Are ≥18 and <65 years of age at the time of randomization

**Disease Characteristics**

2. Are currently not hospitalized
3. Have one or more mild or moderate COVID-19 symptoms (CDC 2020; FDA 2021)
   - i. Fever
   - ii. Cough
   - iii. Sore throat
   - iv. Malaise
   - v. Headache
   - vi. Muscle pain
   - vii. Gastrointestinal symptoms
   - viii. Shortness of breath with exertion
   - ix. Nasal congestion or runny nose
   - x. New loss of smell
   - xi. Chills
4. Must have sample collection for first positive SARS-CoV-2 viral infection determination ≤3 days prior to the administration of study drug

**Sex**

5. Men or non-pregnant women
   Reproductive and Contraceptive agreements and requirements are provided in the main protocol, Section 10.4, Appendix 4. Contraceptive use by men or women should be consistent with local regulations for those participating in clinical studies.

#### 2.8. Cohort Escalation Criteria

Data will be evaluated on an ongoing basis until the highest planned dose has been administered.

The decision to dose the next cohort in the addendum will be made when all participants from the previous cohort have been dosed and safety data, including safety laboratory data, AEs and vital signs, are assessed for at least 48 hours after study drug administration by the investigator(s) and Lilly sponsor team.

Available PK and PD data may also be used to guide dose adjustment.

If temporary stopping criteria are met (Section 2.9), dosing will be temporarily stopped and no further participants will be dosed until a full safety review of the study has taken place. The assessment committee (see Section 6.1.2 of main protocol) will be engaged for the full safety review with the sponsor and investigator.

#### 2.9. Temporary Stopping Criteria

Dosing will be temporarily halted, and no further participants will be dosed until a safety review of the study has taken place if:

- Two or more participants at a given dose level develop severe or severe/potentially life-threatening acute AEs related to the administration of study drug (see table in main protocol, Section 6.1.1.2), during or within 2 hours of completion of the administration, that do not resolve with a reduced infusion rate and/or supportive care.
  **OR**
- Two or more participants at a given dose level develop severe AEs within 4 days of dosing which, in the opinion of the investigator, cannot be attributed to the primary disease, concomitant medications or extraneous circumstances with a reasonable possibility.

| Topic | Location in Main Protocol |
|---|---|
| DAIDS table describing severity of reactions | Section 6.1.1.2 |
| Definition of AEs | Section 10.3.1 |
| Assessment of Intensity/Severity | Section 10.3.3 |

Changes to the planned dosing schedule must be appropriately documented and communicated with the study personnel and the IRB/IEC before dosing continues.

#### 2.10. Study Interventions(s) Administered

| **Intervention Name** | Placebo | LY3853113 | LY3853113 | LY3853113 + LY3819253 + LY3832479 |
|---|---|---|---|---|
| **Dose Formulation** | 0.9% sodium chloride solution | Solution | Solution | Solution |
| **Dosage Level(s) (mg)** | Not applicable | 70, 175, 1750 | [redacted] | 175 + 700 + 1400 |
| **Route of Administration** | IV infusion, [redacted] | IV infusion | [redacted] | IV infusion |
| **Use** | Placebo | Experimental | Experimental | Experimental |
| **IMP and NIMP** | IMP | IMP | IMP | IMP |
| **Sourcing** | Commercially available 0.9% sodium chloride solution | From Lilly | From Lilly | From Lilly |

| | | | | |
|---|---|---|---|---|
| **Packaging and Labeling** | Commercially available 0.9% sodium chloride solution | Study Intervention will be provided in glass vials and will be labeled appropriately. | Study Intervention will be provided in glass vials and will be labeled appropriately. | Study Intervention will be provided in glass vials and will be labeled appropriately. |

**Note:** Depending on the results of a given internal data review, doses may be adjusted.

Infusion [redacted] dose preparation information may be found in the pharmacy preparation instructions.

**Arm A IV administration**

*Single Administration*

Participants should be monitored for at least 1 hour after completion of infusion. The infusion rate may be reduced as deemed necessary if an infusion reaction is observed (main protocol, Section 6.1.1.2).

*Multiple Administrations*

In the event of multiple administrations, participants should receive LY3819253 and LY3832479 or placebo first and should be monitored for at least 30 minutes after completion of the first administration and before administration of LY3853113 or placebo. Participants should be monitored for at least 1 hour after second administration. Further details will be included in the pharmacy preparation incstructions. The infusion rate may be reduced as deemed necessary if an infusion reaction is observed (main protocol, Section 6.1.1.2).

[redacted]

[redacted]

[redacted]

CCI [redacted]

#### 2.12. Study Assessments and Procedures

Study procedures and their timing are summarized in the SoA (Section 2.2).

Immediate safety concerns should be discussed with the sponsor immediately upon occurrence or awareness to determine if the participant should continue or discontinue study intervention.

##### 2.12.1 Vital Signs

Vital signs will be measured as specified in the SoA (Section 2.2) and as clinically indicated. Vital signs include

- Body temperature
- Blood pressure
- Pulse rate
- Respiration rate
- Saturation of peripheral oxygen, and
- Supplemental oxygen flow rate, FiO2 if known, and method of delivery, if applicable.

These tables outline Day 1 vital signs data collection on the CRF in relation to the infusion(s). Infusion times may vary depending on the participant.

Single Administration

| **Timepoint (minutes)** | **Collect data on CRF** |
|---|---|
| Immediately before infusion | Yes |
| If infusion is <15 minutes, immediately following completion of infusion | Yes |
| During Infusions > 15 minutes, as possible (if applicable) | -- |
| 15 | No |
| 30 | Yes |
| 45 | No |
| 60 | Yes |
| After infusion – every 30 minutes for 1 hour after the end of the infusion | -- |
| end of infusion +30 minutes | Yes |
| end of infusion +60 minutes | No |

Multiple Administrations

| Timepoint (minutes) | Collect data on CRF |
|---|---|
| Immediately before infusion | Yes |
| If infusion is <15 minutes, immediately following completion of first infusion | Yes |
| During infusion > 15 minutes, as possible (if applicable) | -- |
| 15 | No |
| 30 | Yes |
| 45 | No |
| 60 | Yes |
| After last infusion – every 30 minutes for 1 hour after the end of the last infusion | -- |
| end of infusion +30 minutes | Yes |
| end of infusion +60 minutes | No |

#### 2.13. Statistics

Statistical analyses of this substudy will be the responsibility of the Sponsor or its designee.

**Sample Size**

The sample size for this addendum is customary for first-in-human studies to evaluate safety.

**Populations for Analyses**

This table defines the populations for analysis.

| Population | Description |
|---|---|
| Entered | All participants who sign the informed consent form for the addendum. |
| Efficacy - Addendum | All participants who were allocated and received study intervention in the addendum and provided at least one post-baseline measure for the relevant endpoint. Participants will be analyzed according to the intervention to which they were randomized. (Intention to treat). |
| Safety - Addendum | All participants allocated to treatment in the addendum and who received study intervention. Participants will be analyzed according to the intervention they received. |
| Pharmacokinetic - Addendum | All participants who were allocated and received addendum intervention in the study and have evaluable PK sample. Participants will be analyzed according to the intervention they received. |

**Statistical Analyses**

All analyses in this addendum will be summarized by cohort and administration method, no inferential statistics will be performed. Data from participants in this addendum will be summarized separately from participants in the main protocol. Refer to the PYAH Statistical Analysis Plan for details on handling dropouts or missing data.

**Interim Analyses**

An interim analysis of safety data may be conducted at any time for participants who have reached Day 3.

**Leo Document ID = ef6e577b-7ace-47ec-b1f2-2e400a08c625**

**Approver:** PPD

**Approval Date & Time: 16-Mar-2021 21:05:11 GMT**

**Signature meaning: Approved**

**Approver:** PPD

**Approval Date & Time: 16-Mar-2021 21:14:37 GMT**

**Signature meaning: Approved**
